# Effects of exercise on brain health outcomes in children with overweight/obesity: the ActiveBrains randomized controlled trial

**DOI:** 10.1101/2022.03.17.22272506

**Authors:** Francisco B Ortega, Jose Mora-Gonzalez, Cristina Cadenas-Sanchez, Irene Esteban-Cornejo, Jairo H Migueles, Patricio Solis-Urra, Juan Verdejo-Román, María Rodriguez-Ayllon, Pablo Molina-Garcia, Jonatan R Ruiz, Vicente Martinez-Vizcaino, Charles H Hillman, Kirk I Erickson, Arthur F Kramer, Idoia Labayen, Andrés Catena

**Affiliations:** PROFITH “PROmoting FITness and Health Through Physical Activity” Research Group, Sport and Health University Research Institute (iMUDS), Department of Physical and Sports Education, Faculty of Sport Sciences, University of Granada, 18071 Granada, Spain; Faculty of Sport and Health Sciences, University of Jyväskylä, Jyväskylä, Finland; Department of Biosciences and Nutrition, Karolinska Institutet, Huddinge, Sweden; Department of Health, Medicine and Caring Sciences, Linköping University, Linköping, Sweden; Faculty of Education and Social Sciences, Universidad Andres Bello, Viña del Mar, Chile; Department of Personality, Assessment & Psychological Treatment and Mind, Brain, and Behavior Research Center (CIMCYC), University of Granada, 18071, Granada, Spain; Laboratory of Cognitive and Computational Neuroscience (UCM-UPM), Centre for Biomedical Technology (CTB), Madrid, Spain; Department of Epidemiology, Erasmus MC University Medical Center, Rotterdam, the Netherlands; Biohealth Research Institute, Physical Medicine and Rehabilitation Service, Virgen de las Nieves University Hospital, Granada, Spain; Health and Social Research Center, Universidad de Castilla La Mancha, Cuenca, Spain; Faculty of Health Sciences, Universidad Autónoma de Chile, Talca, Chile; Department of Psychology, Northeastern University, Boston, MA, USA; Department of Physical Therapy, Movement & Rehabilitation Sciences, Northeastern University, Boston, MA, USA; Brain Aging & Cognitive Health Lab, Department of Psychology, University of Pittsburgh, Pittsburgh, PA, USA; Murdoch University, College of Science, Health, Engineering, and Education, Perth, Western Australia; Beckman Institute, University of Illinois at Urbana-Champaign, Champaign, IL, USA; Department of Health Sciences and Institute for Innovation & Sustainable Development in Food Chain (IS-FOOD) at the Public University of Navarra; and IdiSNA, Navarra Institute for Health Research; Pamplona, Spain; School of Psychology, University of Granada, Granada, Spain

## Abstract

**Objectives:** To investigate the effects of exercise on intelligence, executive functions, academic performance and brain outcomes in children with overweight/obesity. In secondary analyses, we explored potential mediators and moderators of the exercise effects.

**Methods:** A total of 109 children (8-11.9y) with overweight/obesity were randomized (intention-to-treat) and 90 (82.6%) completed the post-exercise evaluation and attended ≥70% of the exercise sessions (per-protocol). Participants in the control group continued with their usual routines and received lifestyle recommendations, whereas the exercise group attended 3 sessions/week of aerobic plus resistance training during 20 weeks. Intelligence, executive functions (cognitive flexibility, inhibition, working memory) and academic performance were assessed with standardized tests; and hippocampal volume with magnetic resonance imaging (MRI).

**Results:** In per-protocol analyses, the exercise intervention improved intelligence and cognitive flexibility (medium-large effect sizes observed, 0.4-0.7 SDs). These main effects were consistent in intention-to-treat analyses and after multiple-testing correction. Moreover, we found a positive, small-magnitude (i.e., 0.2-0.3 SDs) effect of exercise on academic performance (total, mathematics and problem solving), which was partially mediated by cognitive flexibility. Inhibition, working memory, hippocampal volume, and other brain MRI outcomes studied were not affected by our exercise program. Our intervention increased cardiorespiratory fitness performance (0.4 SDs) and these changes in fitness mediated some of the effects. Effects were mostly consistent across the studied moderators, except for larger improvements for intelligence in boys compared to girls.

**Conclusion:** Exercise positively impacts intelligence and cognitive flexibility during development in children with overweight/obesity, without changes in the structural and functional brain outcomes studied.

**Trial Registration:** ClinicalTrials.gov Identifier: NCT02295072

**SUMMARY BOX:** *What is already known on this topic:*

- Pediatric obesity is associated not only with poorer physical health but also with poorer cognitive and brain development.
- Previous exercise interventions have mostly focused on executive functions and other dimensions of cognition, yet is largely unknown the extent to which exercise can improve intelligence during childhood, and actually, at any period of life.
- Studies integrating effects of exercise on behavioral and brain magnetic resonance outcomes in a single article are scarce.
- A in-depth study of the exercise characteristics (mode) and intensity, potential compensatory/contamination effects and role of potential mediators and moderators of the exercise effects is warranted.

*What this study adds:*

- A 20-week randomized controlled trial of exercise improved intelligence and cognitive flexibility in preadolescent children with overweight/obesity.
- Moreover, we found a positive, small-magnitude effect of exercise on academic performance, which was partially mediated by cognitive flexibility.
- Cardiorespiratory peak performance mediated some yet not all the exercise effects observed.
- The structural and functional brain outcomes studied were not affected by participation in the exercise program.

**How this study might affect research, practice or policy:** Our investigation suggests that exercise can positively impact intelligence and cognitive flexibility during a sensitive period of brain development in childhood. This stimulus can positively affect academic performance, as shown in our study, indicating that an active lifestyle during preadolescent development may lead to more successful life trajectories. This is particularly important in children with overweight/obesity who are known to be at higher risk of poorer physical and brain health.

## INTRODUCTION

The prevalence of overweight/obesity in youth has quadrupled worldwide from 1975 to 2016 (from 4 to 18%)^1^. Emerging evidence suggests that obesity might negatively impact brain health (i.e., cognitive and brain development)^2–4^. It is therefore necessary to identify effective strategies to attenuate these adverse consequences. In this context, physical exercise is a candidate to produce such positive stimuli, as it exerts multi-systemic benefits on human organs, including the brain^5,6^. Existing exercise-based interventions have mostly targeted executive functions and other dimensions of cognition (e.g., processing speed, language, etc.)^7–9^, yet evidence on the effect of exercise on intelligence and its components, i.e., crystallized and fluid,^10^ is lacking. Against traditional beliefs, the notion that intelligence is “malleable” despite its high heritability is gaining support^11^; yet more research is warranted.

While most previous studies focused on behavioral outcomes (e.g., executive functions and other dimensions of cognition), only a few randomized controlled trials (RCTs) in children have investigated the effects of exercise on brain structure and function ^12–20^. There is a need for high-quality RCTs that combines behavioral and brain imaging outcomes, as well as a better characterization of the exercise dose administered in the interventions^21,22^. Moreover, previous studies in animals^23^ and in older adults^23–25^ have pointed to hippocampal volume as a critical brain outcome affected by exercise. Although the hippocampus is not a brain region directly linked to intelligence, it is a central hub in networks that support executive function and memory. The effects of exercise on this brain region during a period of brain growth remain under investigated. Further, a comprehensive investigation, including a broader set of magnetic resonance imaging (MRI) outcomes, is needed to understand the overall effect of exercise on brain structure and function.

The ActiveBrains RCT^26^ included a broad set of both behavioral and MRI outcomes, and was designed to test the effects of exercise on brain health in pediatric obesity. Our primary aim (*a priori*-planned) was to investigate the effects of a 20-week exercise program on behavioral outcomes, including intelligence, executive function (i.e., cognitive flexibility, inhibition and working memory) and academic performance, as well as on hippocampal volume as a primary region of interest in children with overweight/obesity.

In secondary analyses (*a posteriori*-planned), we explored potential mediators and moderators of the main exercise effects observed in this intervention. First, we investigated cardiorespiratory fitness (CRF) as the main candidate mediator^27,28,37,38,29–36^; and explored other specific brain regions of interest (e.g., prefrontal cortex due to its relationship with intelligence and cognitive flexibility^39–41^), and broader brain structural and functional changes (hypothesis-free analyses) as potential mediators. Second, we tested potential moderators (sex, age, maturation, socioeconomic status and baseline performance) of the intervention effects^42^. Third, we interrogated potential compensatory and contamination effects on the daily activity levels assessed with accelerometers. Lastly, we analyzed the exercise dose, i.e., the actual volume and intensity of the intervention via heart rate (HR) monitoring, as this might have a direct impact on the magnitude of intervention effects.

## METHODS

A brief description of the material and methods is discussed below. Further details are provided in **Extended Methods in Supplement 1**.

### Study design, participants

The ActiveBrains trial^26^ is a parallel-group RCT conducted in children aged 8-11 years with overweight/obesity. A total of 109 participants were randomly allocated into a control or an exercise group. All pre- and post-exercise data were collected from November 2014 through June 2016, with neuroimaging data processing and analyses being conducted during 2017-2021. The parents or legal guardians signed an informed consent to participate in the study. The CONSORT (Consolidated Standards of Reporting Trials) checklist is presented as **Supplement 2**.

### Intervention and control

The participants in the control group continued with their usual routines. Both control and exercise groups were provided with information about healthy nutrition and physical activity recommendations at the beginning of the study. The exercise group was instructed to attend at least 3 (out of 5 offered) supervised sessions/week. Sessions lasted 90 min (60 min aerobic plus 30 min resistance exercises). To increase motivation and adherence, exercise sessions were based on games and playful activities that involved coordinative exercises.

### Outcome measurements

#### Intelligence

Crystallized, fluid and total (i.e., crystallized plus fluid) intelligence was assessed by the Spanish version of the Kaufman Brief Intelligence Test (K-BIT)^43^.

#### Executive function

Cognitive flexibility was assessed using the Design Fluency Test and the Trail Making Test. Inhibition was evaluated with a modified version of the Stroop test (paper-pencil version) ^44–46^. Working memory was measured by a modified version of the Delayed Non-Match-to-Sample (DNMS) computerized task^47^.

#### Academic performance

Academic performance was assessed by the Spanish version of the Woodcock-Johnson III Tests of Achievement^48^.

#### Brain MRI outcomes

The structural and functional MRI outcomes studied are summarized in **Figure 1**. The MRI acquisition and the specific processing steps for each analysis are individually detailed in **Extended Methods in Supplement 1**.

**Figure 1.**
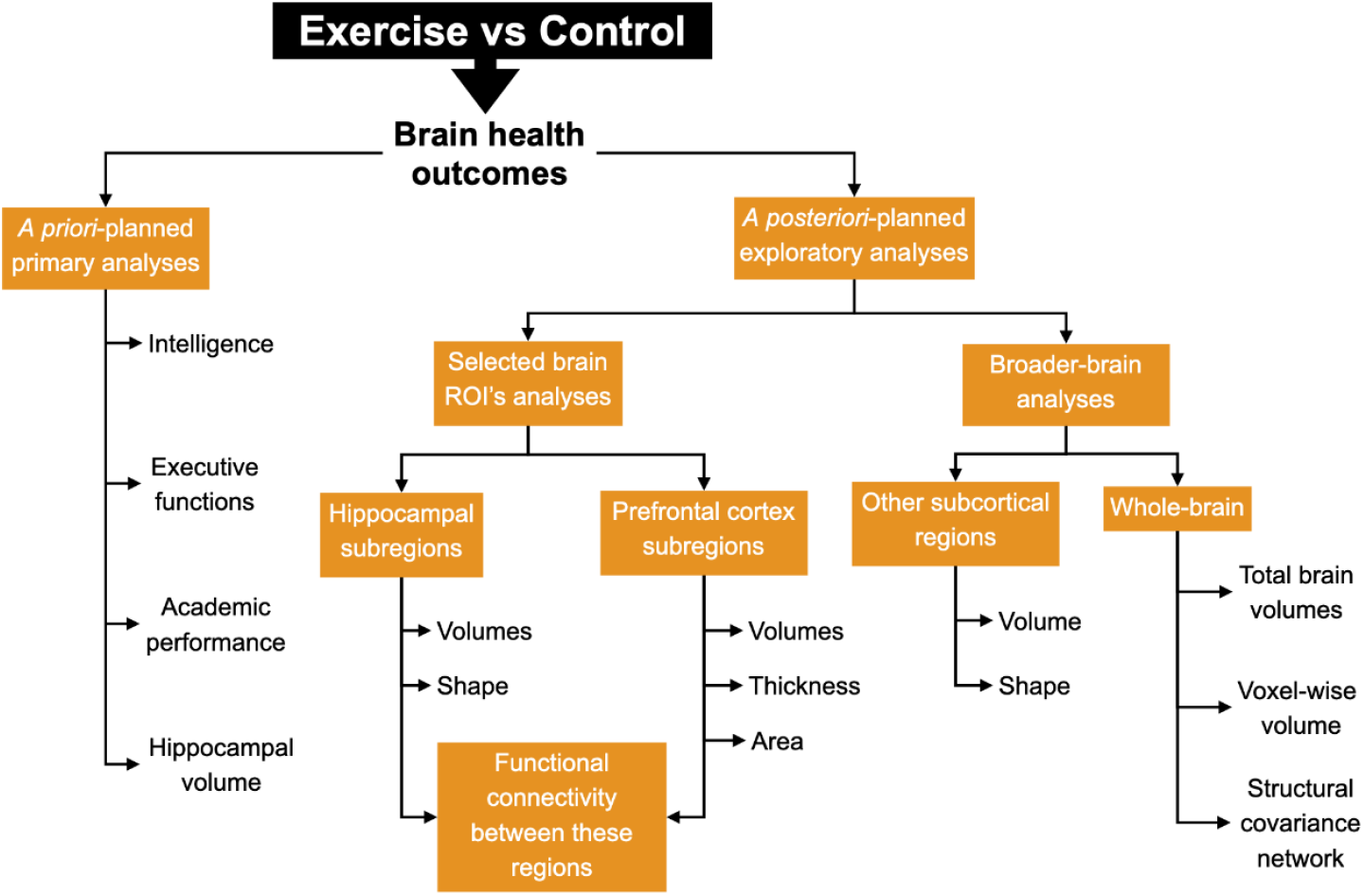
Graphical illustration of the *a priori*-planned main analyses of the study, as well as the *a posteriori*-planned exploratory analyses conducted on different brain health outcomes. ROI indicates region-of-interest

#### Cardiorespiratory fitness

CRF was evaluated under laboratory conditions using a gas analyzer (General Electric Corporation) while performing a maximal incremental treadmill (hp-cosmos ergometer, Munich, Germany) test^49^.

#### Biological maturation

Peak height velocity (PHV) is a common indicator of maturity in children and adolescents^50^. PHV was calculated through Moore’s equations^51^.

#### Socioeconomic status

Parents self-reported their highest educational level attained and current occupation, as described elsewhere^26,52^.

### Overall physical activity assessment before and during the intervention

Activity patterns at baseline and during the intervention (week 10) were assessed with accelerometers worn at hip and wrist (GT3X+, ActiGraph, Pensacola, FL, US) as described elsewhere^53^.

### Statistical analyses

We report the findings from the per-protocol analysis in the main article and the intention-to-treat analyses in **Supplement 1** based on two reasons: 1) We aimed to study the efficacy of the program rather than the effectiveness; and 2) in the field of neuroimaging it is rarely done and technically difficult to apply imputation methods on images. The analyses of the effects of the intervention were tested using analysis of covariance (ANCOVA) with behavioral outcomes as well as on a set of MRI outcomes (with hippocampal volume as a primary region of interest) as dependent variables in separate models, group (exercise versus control) as fixed factors and baseline of the study variable as covariate. The intervention effects are presented as z-scores of change, which indicate how many standard deviations (SDs) the post-exercise program values changed with respect to the baseline mean and SD. Therefore, it can be interpreted as a standardized effect size of the change^54^. Results in raw units of measure are also provided in eTables in Supplement 1. We report significant findings according to the classic P-value threshold of 0.05. Additionally, we applied multiple testing corrections on the primary outcomes^55^. *A posteriori-planned* analyses consisted on exploring potential mediators and moderators. Our mediation analyses are in line with the AGReMA (A Guideline for Reporting Mediation Analyses: https://agrema-statement.org/) statement and the corresponding checklist is provided as **Supplement 3**. The statistical procedures were performed using the SPSS software (version 25.0, IBM Corporation) and R software (v. 3.1.2, https://www.cran.r-project.org/).

## RESULTS

Baseline characteristics of the study sample are presented in **eTable 1 in Supplement 1**. The flowchart of the study is presented in **Figure 2**. Out of the 109 randomized participants, 96 completed the study (12% attrition rate) and 90 met the criteria for per-protocol analyses (83% of the original sample). A graphical illustration of the *a priori-* and *a posteriori*-planned analyses on brain health outcomes conducted in the current study is presented in **Figure 1**. Additional details are provided in **Extended Results in Supplement 1**.

**Figure 2.**
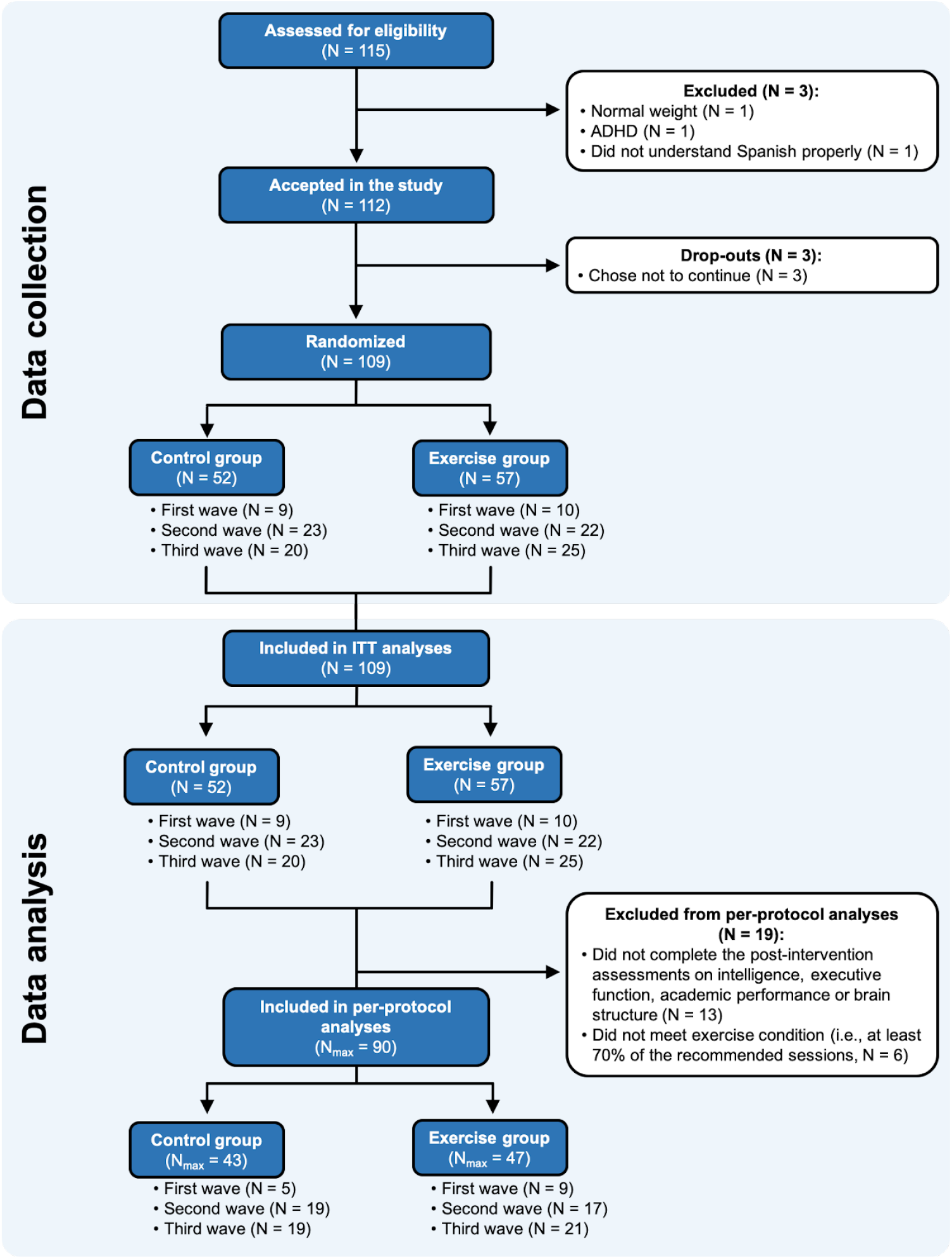
CONSORT flow diagram. ADHD = Attention-deficit hyperactivity disorder. ITT = Intention-to-treat. For final ITT analyses, those participants who left the study during the intervention or did not complete the post-exercise program assessments were imputed (see Statistical analysis section). N_max_ = Maximum sample size included in the analyses. The actual N for each variable can be seen in eTables in Supplement 1.

### 1. A priori-planned analyses. Effects of the exercise intervention on primary outcomes: intelligence, executive functions, academic performance and hippocampal volume

The largest effect size observed in the ActiveBrains exercise program was for crystallized intelligence, with the exercise group improving from pre-to post-exercise compared to the control group 0.72 SDs (P=0.0000003, **Figure 3 and eTable 2 in Supplement 1**). Total intelligence also improved significantly more in the exercise group than in the control group (0.62 SDs, P=0.00008). In addition, exercise positively affected a composite score of cognitive flexibility, derived from two cognitive flexibility tests (0.42 SDs, P=0.005). Within this composite, the largest improvement was observed for performance on the cognitive flexibility test 1 (i.e., Design Fluency Test) (0.5 SDs, P=0.003). The exercise program had a null effect on the other two dimensions of executive function: inhibition and working memory (|0.04| SDs and P=0.8, for both outcomes).

**Figure 3.**
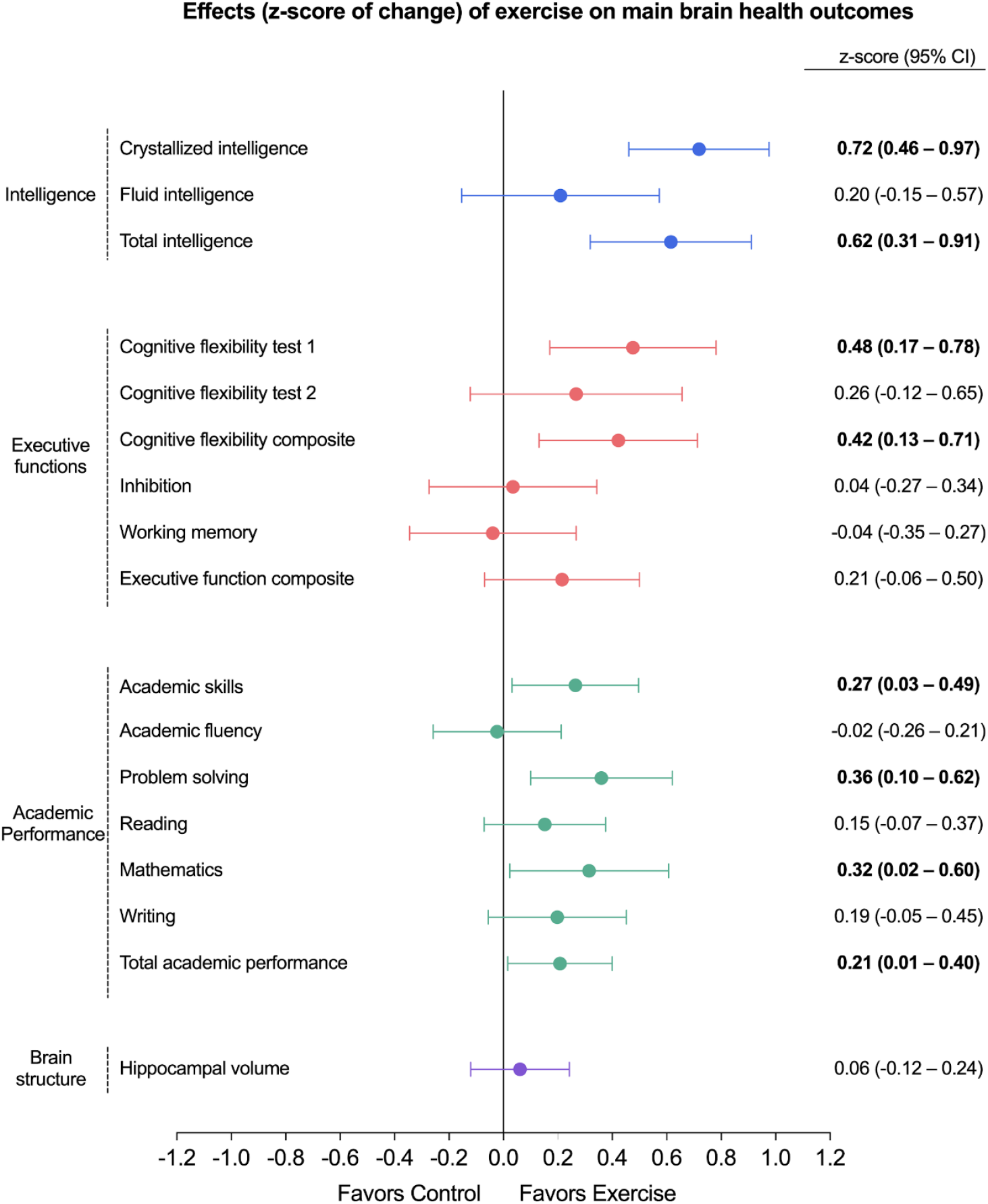
Per-protocol effects of the *ActiveBrains* exercise program on the main brain health outcomes. Dots represent the between-groups difference in z-score values of change, i.e., post-exercise outcomes with respect to the baseline mean and standard deviation. Bars represent 95% confidence intervals (CIs). Each analysis was adjusted for baseline outcomes. Bold font indicates significant effect at P<0.05 (or by the 95% CI not including zero). Each color represents one of the four main brain health dimensions (i.e., intelligence, executive function, academic performance, or brain structure). Intelligence outcomes (i.e., Crystallized intelligence, Fluid intelligence, and Total intelligence) were measured by the Kaufman Brief Intelligence Test. Cognitive flexibility test 1 refers to the Design Fluency Test output. Cognitive flexibility test 2 refers to the Trail Making Test output. Cognitive flexibility composite z-score was calculated as the re-normalized mean of the z-scores for Cognitive flexibility test 1 and Cognitive flexibility test 2. Inhibition was measured by the Stroop Color-Word Test. Working memory was measured by the Delayed Non-Match-to sample computerized task. Executive function composite z-score was calculated as the re-normalized mean of the z-scores for Cognitive flexibility, Inhibition, and Working memory. Academic performance outcomes were obtained from the Spanish version of the Woodcock Johnson III Test of Achievement. Academic skills are the sum of components based on basic skills such as reading decoding, mathematics calculation, and spelling. Academic fluency is the sum of tests based on reading, calculation and writing fluency. Problem solving is the sum of the components based on solving academic problems in reading, mathematics, and writing. Total academic performance is the overall measure of the academic performance based on reading, mathematics, and writing. Hippocampal volume was obtained using the FMRIB’s Integrated Registration and Segmentation Tool (FIRST). Two of the cognitive tests, i.e., the cognitive flexibility test 2 (Trail Making Test) and the inhibition test (Stroop Color-Word Test), are originally expressed inversely, which means that lower scores indicate better performance. To simplify the visual interpretation of the main findings in this Figure, we inverted these two scores so that they can be interpreted in the same fashion as the rest of outcomes (i.e., higher score better performance). On the other hand, we express these cognitive tests in their original units and not inverted in **eTables 2** and **19 in Supplement 1**.

For academic performance (**Figure 3 and eTable 3 in Supplement 1**), exercise significantly improved total academic performance and, particularly, mathematics, problem solving and academic skills. Overall, the effect sizes observed for academic performance were all of small magnitude, ranging from 0.21-0.36 SDs (P-values ranged from 0.035-0.007). The exercise program had a small, non-significant effect on reading and writing skills and a null effect on academic fluency. In exploratory analyses, we observed that the positive impact of exercise on total academic performance, mathematics and academic skills was mediated (30-39% of mediation) by exercise-induced improvements in cognitive flexibility (**eFigure 1A-C in Supplement 1**). Moreover, we observed that the improvements in academic problem solving were mediated (15% of mediation) by improvements in exercise-induced changes in fluid intelligence (**eFigure 1D in Supplement 1**). Alternatively, our exercise program did not have an effect on overall hippocampal volume (**Figure 3 and eTable 4 in Supplement 1)**.

After correction for multiple comparisons on the primary outcomes (the 17 outcomes shown in **Figure 3**), the larger effects on intelligence (crystallized and total) and cognitive flexibility persisted with corrected P≤0.001 and P=0.02 respectively. Likewise, the effects observed on problem solving persisted significant (corrected P=0.02), whereas became borderline non-significant for mathematics, academic skills and total academic performance (all corrected P=0.07).

### 2. A posteriori-planned analyses on brain MRI outcomes

As shown in Figure 1 we explored the effects of the intervention on a set of brain MRI outcomes, including volumetric analyses of hippocampus subregions and prefrontal cortex (**eTables 4 and 5 in Supplement 1**), cortical thickness and surface area of prefrontal cortex and subregions (**eTables 6 and 7 in Supplement 1**), functional connectivity between hippocampus and prefrontal cortex (**eTables 8 to 13 in Supplement 1**). Moreover, we studied the effects of the intervention using a broader brain approach, including: gray matter volumes of subcortical brain structures (**eTable 14 in Supplement 1**), morphologic (shape) analysis of subcortical brain structures (**eFigure 2 in Supplement 1**), total brain volumes (**eTable 15 in Supplement 1**), whole-brain voxel-wise volumetric analysis and whole-brain structural covariance network analysis (**eFigure 3 in Supplement 1**). In short, our intervention did not have significant effect in any of these MRI outcomes studied.

### 3. Effects of the intervention on CRF and its role as mediator

The exercise program improved CRF as indicated by treadmill time-to-exhaustion, with 0.42 SDs larger improvement in the exercise vs. control group (P=0.04) (**eTable 17 in Supplement 1**). A consistent improvement, yet smaller and non-significant, was observed in peak oxygen consumption (VO2peak) expressed in mL/kg/min (0.29 SDs, P=0.13). The effects of the exercise program on crystallized intelligence, problem solving and total academic performance were significantly mediated by improvements in CRF (i.e., time-to-exhaustion), with a mediation effect of 10-20% (**Figure 4**).

**Figure 4.**
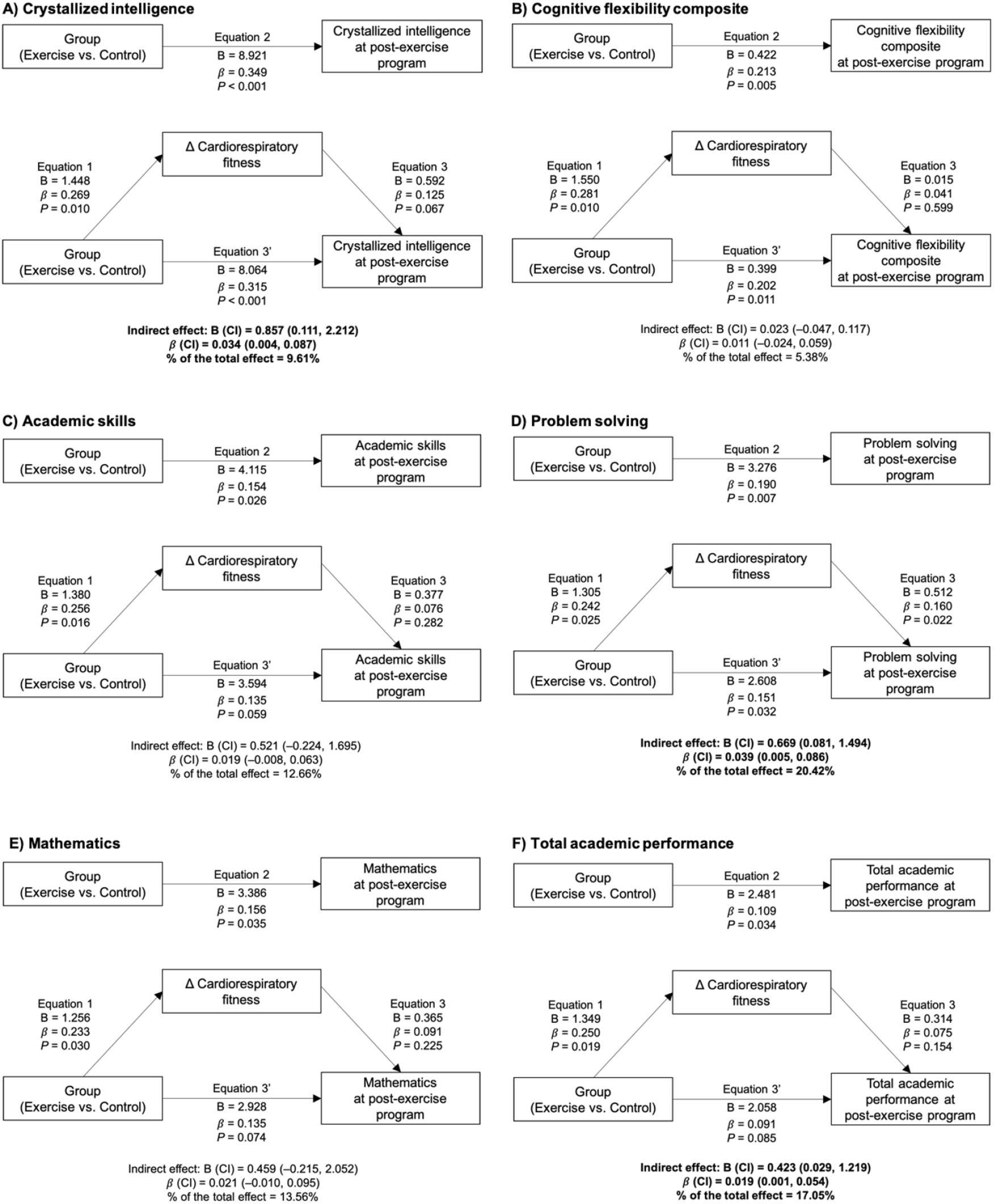
Cardiorespiratory fitness’ change mediation models of the intervention effects (i.e., exercise vs. control) on intelligence, cognitive and academic performance outcomes in children with overweight/obesity. Each analysis was adjusted by the respective intelligence, cognitive or academic performance outcomes at baseline. Bold font indicates significant indirect effect at P<0.05. B indicates unstandardized regression coefficient and *β* indicates standardized regression coefficient. CI indicates Confidence Interval. Delta (Δ) cardiorespiratory fitness expresses the change in total completion time (min) of the treadmill test at post-exercise program with respect to the total completion time (min) at baseline, since it was the main cardiorespiratory fitness outcome influenced by the exercise program. Crystallized intelligence was measured by the Kaufman Brief Intelligence Test. Cognitive flexibility composite z-score was calculated as the re-normalized mean of the z-scores for Design Fluency Test and Trail Making Test. Academic skills are the sum of components based on basic skills such as reading decoding, mathematics calculation, and spelling. Problem solving is the sum of the components based on solving academic problems in reading, mathematics, and writing. Total academic performance is the overall measure of the academic performance based on reading, mathematics, and writing.

### 4. Moderators of the intervention effects

**Figure 5** shows that the effect sizes of the exercise program were consistent across sex, age and maturation for most of the primary outcomes studied, except for crystallized intelligence, wherein the exercise program was more effective in boys as well as in younger and less mature participants. The sex differences observed could be partially explained by the finding that boys spent more time at high intensity zones (i.e., over their individualized anaerobic threshold monitored with heart rate) (**eTable 18 in Supplement 1**). Finally, we also observed that children with lower socioeconomic status (i.e., low parental educational and occupational levels) showed larger improvements in fluid and total intelligence, as did children with a lower performance at baseline in the intelligence test (**eFigure 4 in Supplement 1)**.

### 5. Other exploratory analyses related to the interpretation of the intervention effects (See further details in Extended Results in Supplement 1)

#### Intention-to-treat and dropout analyses

The main effects of this intervention observed in intelligence and cognitive flexibility remained significant in intention-to-treat analyses (**eTables 19-21 in Supplement 1**), indicating the robustness of the main findings. Moreover, participants dropping out during the study did not differ from those completing the study in any of the behavioral outcomes studied (**eTable 22 in Supplement 1**).

**Figure 5.**
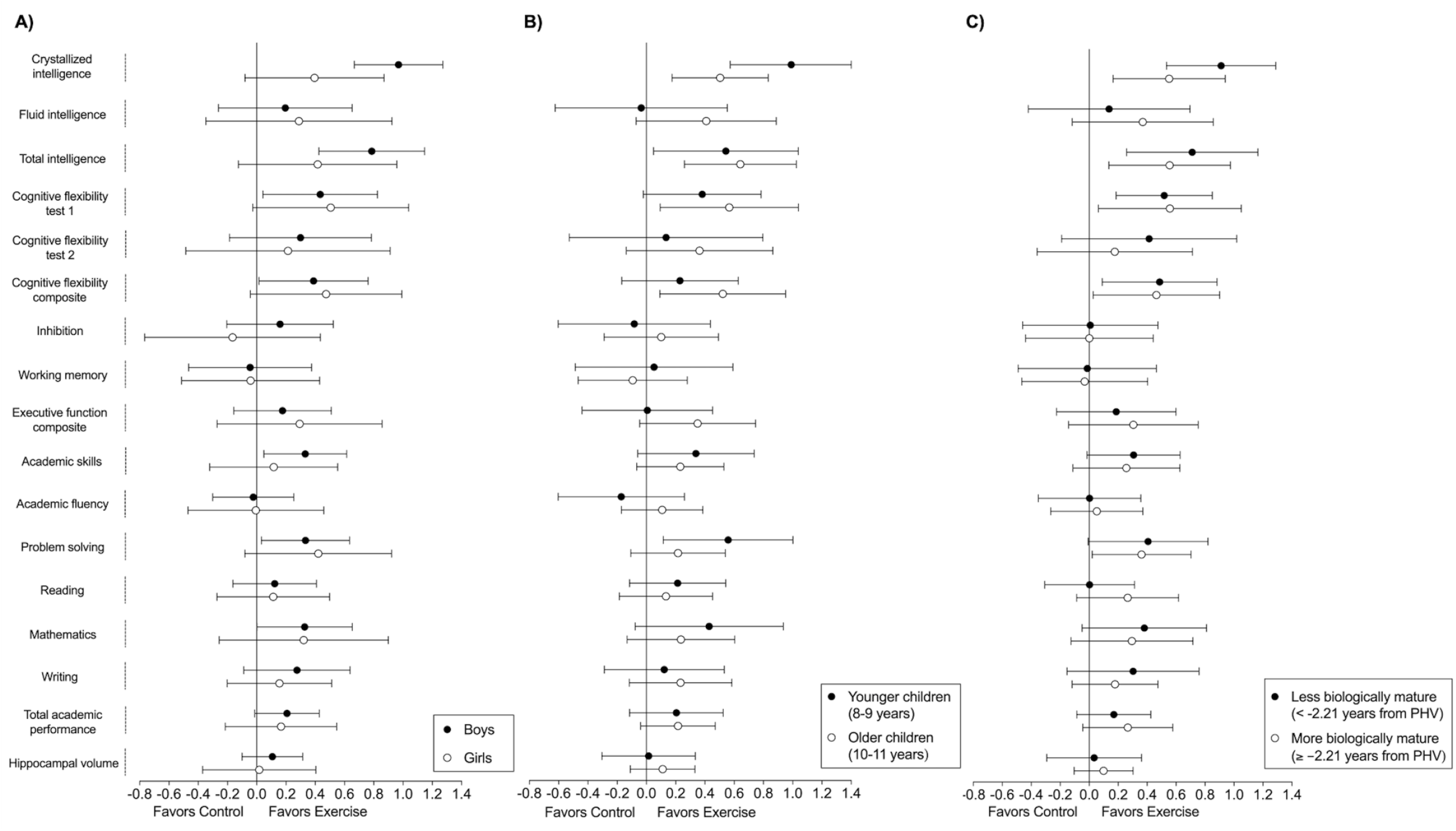
Per-protocol effects of the *ActiveBrains* exercise program on the main brain health outcomes by sex (A), age (B) and biological maturation (C). Each analysis was adjusted by baseline outcomes. Dots represent the between-groups difference in z-score of change, i.e., post-exercise outcomes with respect to the baseline mean and standard deviation. Bars represent 95% confidence intervals. To express biological maturation, years from peak height velocity (PHV) were calculated by subtracting the age of PHV from the chronological age. The difference in years was utilized as a measure of maturity. PHV was dichotomized using the median. Intelligence outcomes (i.e., Crystallized intelligence, Fluid intelligence, and Total intelligence) were measured by the Kaufman Brief Intelligence Test. Cognitive flexibility test 1 refers to the Design Fluency Test output. Cognitive flexibility test 2 refers to the Trail Making Test output. Cognitive flexibility composite z-score was calculated as the re-normalized mean of the z-scores for Cognitive flexibility test 1 and Cognitive flexibility test 2. Inhibition was measured by the Stroop Color-Word Test. Working memory was measured by the Delayed Non-Match-to sample computerized task. Executive function composite z-score was calculated as the re-normalized mean of the z-scores for Cognitive flexibility, Inhibition, and Working memory. Academic performance outcomes were obtained from the Spanish version of the Woodcock Johnson III Test of Achievement. Academic skills are the sum of components based on basic skills such as reading decoding, mathematics calculation, and spelling. Academic fluency is the sum of tests based on reading, calculation and writing fluency. Problem solving is the sum of the components based on solving academic problems in reading, mathematics, and writing. Total academic performance is the overall measure of the academic performance based on reading, mathematics, and writing. Hippocampal volume was obtained using the FMRIB’s Integrated Registration and Segmentation Tool (FIRST). Two of the cognitive tests, i.e., the cognitive flexibility test 2 (Trail Making Test) and the inhibition test (Stroop Color-Word Test), are originally expressed in an inverse way, which means that lower scores indicate better performance. In order to make the visual interpretation of the main findings in these Figure easier, we inverted these two scores so that they could be interpreted in the same fashion as the rest of outcomes (i.e., higher score better performance). On the other hand, we express these cognitive tests in their original units and not inverted in **eTables 2** and **19 in Supplement 1**.

#### Testing potential compensatory and contamination effects

We observed that the exercise group significantly increased their activity levels (P<0.001) during the time frame of the day in which children were participating in the exercise program without reductions (i.e., no compensation) in other moments of the day (**Figure 6 and eFigure 5 in Supplement 1**, results from the hip- and wrist-attached accelerometer, respectively). The control group kept the same levels of daily activity (i.e., no contamination).

**Figure 6.**
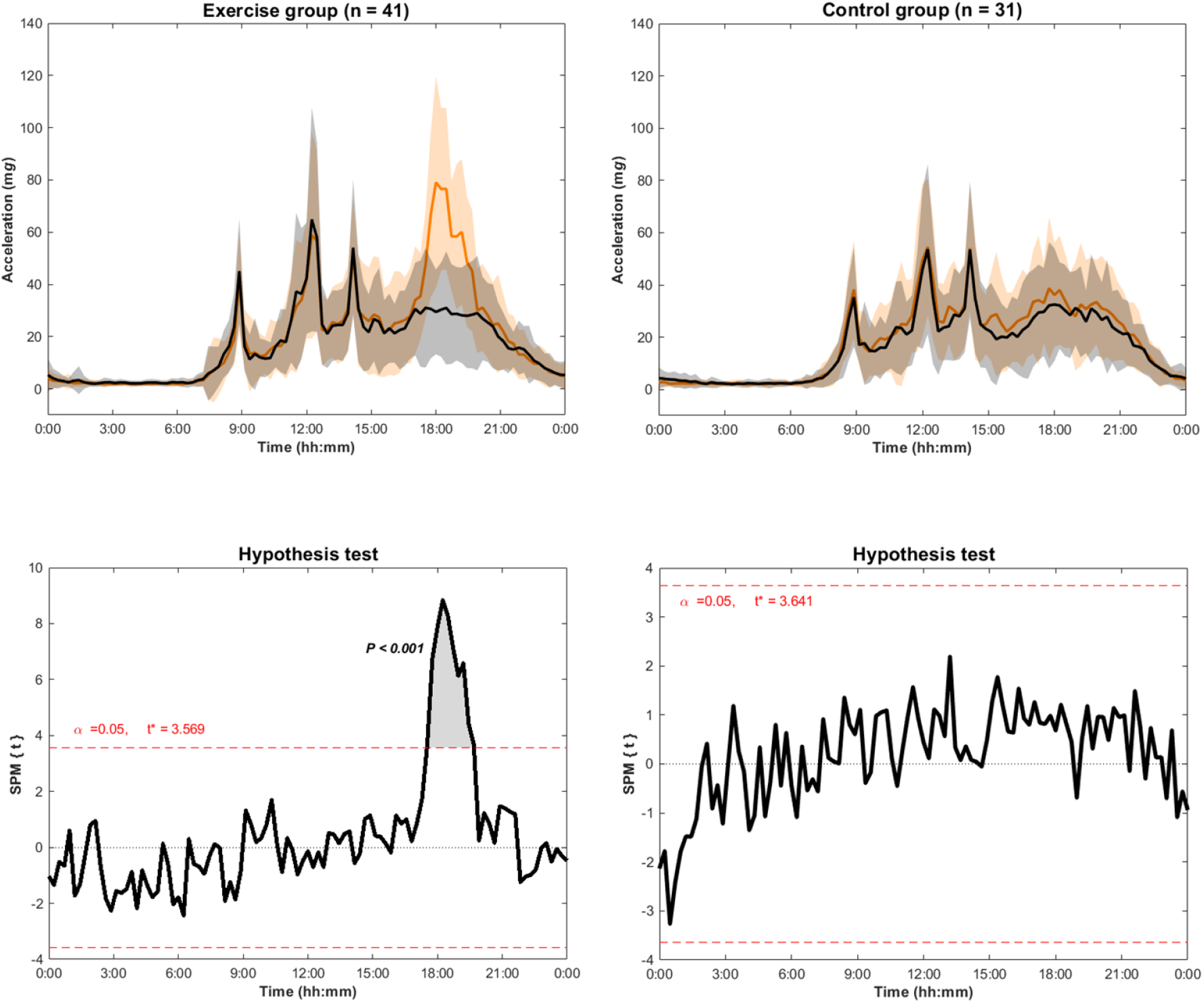
Comparison of the 24 h physical activity patterns derived from aggregated raw accelerations (i.e., Euclidean Norm Minus One accelerations) measured with an accelerometer attached at the right hip at baseline (i.e., black line) and in the middle of the exercise program (i.e., orange line) in exercise and control groups. The hypothesis test shows the threshold (t* = 3.593) at which there are significant physical activity patterns’ differences between baseline and during-exercise periods.

#### Characterization of the actual volume and intensity of the exercise program

We observed an average intensity of 138 bpm (SD=8bpm) per session, indicating that the children trained for more than 1 h at 70% of their maximum HR. In addition, the children accumulated on average 38% of the session time (i.e., 25min) at high intensities over the 80% of their maximum HR (**eFigure 6 in Supplement 1**). As an additional feature of exercise volume received by our participants, we present in **eFigure 7 in Supplement 1** the distribution of the attendance to the exercise sessions.

## DISCUSSION

The ActiveBrains trial contributes to the existing literature with a number of novel findings: 1) a 20-week aerobic+resistance exercise program with a coordinative component, at relatively high intensity for more than 1h, 3 times per week, improved total and crystallized intelligence, cognitive flexibility and academic performance in children with overweight/obesity. We rely mainly on the effects observed on intelligence, particularly on crystallized intelligence, as well as on cognitive flexibility, given the effect sizes and significance observed^56^. In fact, the effects on intelligence and cognitive flexibility outcomes were consistent and robust, persisting after applying multiple test correction as well as in both per-protocol and intention-to-treat analyses. Alternatively, our exercise program had a null effect on other executive functions such as inhibition and working memory, and on hippocampal volume. 2) We did not observe significant effects of exercise on any of the brain MRI outcomes studied (*a posteriori*-planned analyses) and we could not therefore investigate whether changes in brain structure or function mediated the effects observed on behavioral outcomes. 3) The effects of the exercise program on crystallized intelligence, total academic performance and problem solving were partially mediated by exercise-induced improvements in CRF (10-20%, small mediation effect). Interestingly, improvements in most academic performance indicators were largely mediated (∼30-40% of mediation) by exercise-induced changes in cognitive flexibility. 4) The exercise effects were rather consistent across sex, age, socioeconomic status and baseline level subgroups for most of the study outcomes, except for a few cases and particularly for intelligence outcomes that improved more in boys than in girls. An **Extended Discussion** in the context of previous evidence is provided in **Supplement 1**.

The interpretation of the results described above should be made in conjunction with the characteristics of the exercise intervention. We did not observe either compensatory or contamination effects on the daily physical activity levels (a phenomena known to occur sometimes in exercise trials^57,58^), based on our accelerometer analyses. In addition, it is important to highlight that our participants in the exercise group received a relatively high dose of exercise, based on the detailed HR data presented above. It is of utmost importance to investigate the potential compensatory/contamination effects, as well as the difference between planned and actual intensity achieved in the exercise program, since these factors can have a direct impact on the trial’s effects. Further, although our exercise program consisted mostly of aerobic exercise, roughly one third of the actual training time was devoted to resistance training. In this context, recent meta-analyses showed similar benefits on cognition from both aerobic and resistance training^59,60^, as well as for coordinative exercise (yet fewer studies are available for this type of exercise)^42^. Of note, during both the aerobic and resistance training components, the characteristics of the program embedded a definitive coordinative/motor component. This included playful balance, bilateral coordination, hand–eye coordination, and leg–arm coordination exercises as well as spatial orientation and reaction to moving objects/people, and the use of balls, skipping ropes, speed ladders and balloons as the equipment, which together have been defined in previous trials as coordinative/motor exercise^61^. The results from the ActiveBrains trial should therefore be interpreted as the combined effect of aerobic, resistance, and coordinative/motor exercise. Lastly, the exercise program was centered on games that required cooperation with other participants as well as on behaviors modification consistent with rules and instructions. Thus, our exercise program had clear cognitive and social components beyond simply aerobic exercise (e.g. treadmill walking/running or stationary cycling). It has been suggested that adding game elements to an exercise program to make it more cognitively challenging could enhance the effects on executive functions^62–65^. Nevertheless, whether exercise interventions based on active games with higher cognitive demands, like ours, can have a larger impact on brain health outcomes than others with lower cognitive and social demands (e.g., stationary running/biking) needs to be further investigated using RCTs specifically designed to address these research questions.

We acknowledge the limitations of this study. It is unknown whether longer interventions than ours are needed to elicit structural or functional changes in the brain (in **Supplement 1** further discussion on this). Further, although several protocols were adopted to reduce the risk of bias in the evaluations (e.g., randomization after baseline assessment, physical trainers not involved in the evaluations), some of the project staff involved in the post-exercise evaluations were not blinded to the group allocation due to practical reasons, which could add potential bias to the post-intervention measurements. Even assuming an attenuation of the effect sizes after correcting for potential bias, we believe that the main exercise effects on intelligence and cognitive flexibility observed would remain significant given their magnitude, yielding it unlikely to change the study conclusions. Finally, the extent to which the findings from our study conducted in children with overweight/obesity applies to other populations of different characteristics is unknown.

## CONCLUSION

The findings of this study support that intelligence and cognitive flexibility are improved by 20 weeks of exercise of relatively high intensity for more than 1 h, 3 times per week, and during a sensitive period of life, childhood, when the brain is growing and developing. However, we failed to detect which structural or functional changes in the brain may underlie these exercise effects on behavioral outcomes. Alternatively, we observed that exercise-induced changes in CRF explain some of the exercise benefits, yet not most of them. Moreover, our exercise program had small, yet consistent with the literature, effects on academic performance indicators (i.e., mathematics, problem solving and total academic performance), which were mediated by exercise-induced improvements in cognitive flexibility and fluid intelligence. Finally, the intervention effects were generally consistent across the moderators studied, with some exceptions, particularly for intelligence outcomes in which larger effects were observed in boys than in girls. The present study provides a comprehensive investigation on the effects of exercise on cognitive outcomes and academic achievement during growth in the presence of overweight and obesity. However, the brain mechanisms that might explain those effects remains unknown.

## Supporting information

Extended Methods, Extended Results, Extended Discussion, Extended References, eFigures and eTables

CONSORT checklist for conducting and reporting trials

AGReMA checklist for conducting and reporting mediation analyses

Trial protocol

Statistical analysis plan

## Data Availability

We did not obtain children parents consent to widely share the data nor was it included in the IRB protocol.

## Supplemental material

Supplement 1: Extended Methods, Extended Results, Extended Discussion, Extended References, eFigures and eTables

Supplement 2. CONSORT checklist for conducting and reporting trials.

Supplement 3. AGReMA checklist for conducting and reporting mediation analyses.

Supplement 4. Trial protocol

Supplement 5. Statistical analysis plan.

## Twitter

Francisco B. Ortega @ortegaporcel

## Acknowledgments

The authors want to thank other members that have contributed to the ActiveBrains project: Abel Plaza-Florido, Alejandra Mena-Molina, Esther Ubago-Guisado, Ignacio Merino-De Haro, Jose J. Gil-Cosano, Juan Pablo Zavala-Crichton, Lucia V. Torres-Lopez, Luis Gracia-Marco, and Miguel Martín-Matillas from the University of Granada for their participation in the evaluations or intervention in this project; Gala María Enriquez, José Gómez-Vida, José Maldonado, María José Heras, and María Victoria Escolano-Margarit from the “San Cecilio” and “Virgen de las Nieves” Hospitals for assistance with recruitment and screening of participants; Carlos de Teresa, Rosa María Lozano, and Socorro Navarrete from the Centro Andaluz de Medicina del Deporte (CAMD) for medical support and realization of physical health evaluations; María Elisa Merchan, Victoria Muñoz-Hernández, and Wendy Daniela Martínez-Ávila from the University of Granada for their support with the dietary and nutritional evaluations of the project; Ángel Gil, Belén Pastor-Villaescusa, Concepción M. Aguilera, and Maria Cruz Ruiz from the Centre for Biomedical Research of University of Granada for their support with the blood sampling processing and storing; Antonio Verdejo-García from the Monash University, Catherine Davis from the Medical College of Georgia and Jose C. Perales from the University of Granada for their inputs to the project design/conception particularly in the initial phases. All of these persons provided uncompensated assistance. We also thank all of the children and their families for participating in this clinical trial.

## Contributors

FBO and JMG had full access to all the data in the study and take responsibility for the integrity of the data and the accuracy of the data analysis. FBO, IL and AC designed research; FBO, JMG, CCS, IEC, JHM, PSU, JVR, MRA, PMG, JRR, IL, AC performed research; FBO, JMG, CCS, IEC, JHM, PSU, JVR, MRA, PMG, VMV and AC analyzed data (JMG being the person in charge of conducting most of the statistical analyses); and FBO was in charge of drafting the manuscript. All authors critically revised the manuscript, contributed to the conception and interpretation of the analyses and approved the final version of the manuscript to submitted.

## Funding

This study has been mainly supported by grants from the Spanish Ministry of Economy and Competitiveness (DEP2013-47540, DEP2016-79512-R, and DEP2017-91544-EXP), European Regional Development Fund (ERDF)”, by the European Commission (No 667302) and by the Alicia Koplowitz Foundation. Additional funding was obtained from the Andalusian Operational Programme supported with ERDF (FEDER in Spanish, B-CTS-355-UGR18). This study additionally supported by the University of Granada, Plan Propio de Investigación, Visiting Scholar grants and Excellence actions: Units of Excellence; Unit of Excellence on Exercise and Health (UCEES) and by the Junta de Andalucía, Consejería de Conocimiento, Investigación y Universidades and ERDF (SOMM17/6107/UGR). In addition, this study was further supported by the SAMID III network, RETICS, funded by the PN I+D+I 2017-2021 (Spain), and the EXERNET Research Network on Exercise and Health (DEP2005-00046/ACTI; and by the High Council of Sports, 09/UPB/19). J.M-G. has been supported by grants from the Spanish Ministry of Science and Innovation (FPU 14/06837) and the Junta de Andalucía. IEC is supported by the Spanish Ministry of Science and Innovation (FJCI-2014-19563; IJCI-2017-33642; RYC2019-027287-I). C.C-S. has been supported by grants from the Spanish Ministry of Science and Innovation (FPI-BES-2014-068829; FJC2018-037925-I). J.H.M has been supported by the Spanish Ministry of Science and Innovation (FPU15/02645). M.R-A has been supported by the Ramón Areces Foundation. JVR is supported by the Spanish Ministry of Science and Innovation (FJCI-2017-33396, IJC2019-041916-I). P.S-U was supported by a grant from the National Agency for Research and Development (ANID)/BECAS Chile/72180543. This work is part of a Ph.D. Thesis conducted in the Doctoral Programme in Biomedicine of the University of Granada, Spain.

## Competing Interests

None to be declared.

## Patient consent for publication

Not applicable.

## Ethical approval

The ActiveBrains project was approved by the Ethics Committee of the University of Granada, and it was registered in ClinicalTrials.gov (identifier: NCT02295072).

## Data availability statement

We did not obtain children’s parents consent to widely share the data nor was it included in the IRB protocol.

## Notes

### Competing Interest Statement

The authors have declared no competing interest.

### Clinical Trial

NCT02295072

### Clinical Protocols

https://clinicaltrials.gov/ct2/show/NCT02295072?term=NCT02295072&draw=2&rank=1

### Author Declarations

The ActiveBrains project was approved by the Ethics Committee of the University of Granada.

