## Extended Methods, Extended Results, Extended Discussion, Extended References, eFigures and eTables for "Effects of exercise on brain health outcomes in children with overweight/obesity: the ActiveBrains randomized controlled trial"

**This file includes:**

Extended Methods
Extended Results
Extended Discussion
Extended References
eFigures 1 to 7
eTables 1 to 22

#### EXTENDED METHODS

##### *Design and Participants*

The ActiveBrains project is a parallel-group RCT designed to examine the effects of a 20-week physical exercise program on brain, intelligence, executive function and academic performance outcomes, as well as on selected physical and mental health outcomes in children with overweight/obesity (http://profith.ugr.es/activebrains, available in Spanish and English). Details of the ActiveBrains project have been described elsewhere[1]. The present study focused on the primary outcomes, i.e., cognitive and brain outcomes. Briefly, the study was conducted in three waves. The whole data collection period took place between November 2014 and June 2016, with neuroimaging, data processing and analysis being conducted from 2017 to 2021. The eligibility criteria to participate in this study were: 1) children aged 8 to 11.9 years; 2) in the case of girls, pre-menstrual at the moment of baseline assessments; 3) to be classified as overweight or obese at baseline based on sex and age specific World Obesity Federation cut-off points[2,3]; 4) not to have any physical disability or neurological disorder that impeded exercise; 5) not to use medications that influenced central nervous system function; 6) right-handed as assessed by the Edinburgh inventory[4]; and 7) not to have previous diagnosis of attention-deficit hyperactivity disorder (ADHD) or a score above the 85$^{th}$ percentile as measured by the ADHD rating scale[5].

The recruitment process started by contacting families of children with overweight/obesity from databases at the Unit of Pediatrics of the University Hospitals San Cecilio and Virgen de las Nieves (Granada, Spain). Additional strategies included contacting the head teacher of both, public and private schools of Granada to spread informative pamphlets. Furthermore, advertising related to the project was broadcasted in the local media through newspaper, radio, and television outlets. Finally, a total of 109 children with overweight/obesity meeting the general inclusion/exclusion criteria[1], were randomized to exercise and control groups and were included in the intention-to-treat analysis; whereas 90 children were included in the per-protocol analyses (see Statistical analysis section) after meeting the following protocol criteria: (1) completed the pre- and post-intervention assessments, and (2) attended at least 70% of the recommended 3 sessions/week (i.e., exercise group). The flowchart of the study is presented in **Figure 1**.

The participants were randomly allocated to an exercise group and participated in the physical exercise program, while the others were allocated to a wait-list control group. The wait-list control group strategy has been previously used[6,7], and was predicated on the fact that individuals randomized to this group also received the exercise program at the completion of their participation in the ActiveBrains trial. A wait-list control group is especially appropriate for this age group, because it represents typical development across the 20-week investigation period. Simple random allocation of participants into exercise or control groups was performed with a ratio of 1:1 using a random number generator in SPSS software for Windows (version 25.0; Armonk, NY, USA) by a 'blinded' individual not involved in the exercise sessions nor outcome evaluations (done by FBO). This method allowed for the equal probability of allocation to one group or another. To reduce the risk of bias, several protocols were followed: 1) the computer random generation was conducted by a person not involved in the outcome evaluations; 2) randomization was performed immediately after the baseline evaluation; and 3) the physical trainers running the exercise program were not involved in the outcome evaluations or randomization. However, due to practical reasons (limited number of project staff due to budget restrictions), some of the staff involved in the post-exercise evaluations were not blinded to the participants' group allocation (see the Limitation's section in the Discussion of the main text).

##### ***Physical exercise program***

*Characteristics of the exercise program*

The physical exercise program had a duration of 20 weeks, and its design was based on meeting the international physical activity guidelines at the time of the study design[8], consistent with recent updates[9]. Both guidelines highlight that physical activity in youth should be mostly aerobic, yet muscle- and bone-strengthening as well as activity of high intensity should be done at least 3 times per week to maximize the health benefits in youth[9]. Participants had the possibility of attending the program daily from Monday to Friday (i.e., we offered 5 sessions/week, 90 min/session). They were instructed to attend to at least 3 times/week, yet we advised the families that "the more, the better" up to the 5 sessions/week offered. Accordingly, the attendance criterion was set to a minimum of 3 times/week, and participants meeting 70% of the recommended sessions were included in per-protocol analyses, as previously stated. In the **eFigure 7 in Supplement 1**, we present the distribution of the attendance to the exercise program.

The physical exercise program was conducted on a group basis (i.e., 3 waves of intervention) and based on active games, with a noticeable emphasis on the playful component in order to increase adherence to the program. Each session was structured into four parts: 1) a 5-10 min warm-up consisting of 1-2 physical games of 5 min each; 2) a 60-min aerobic part consisting of around four to five physical multi-games requiring moderate-to-vigorous intensities, with special emphasis on high-intensity activities; 3) a 20-min resistance training part consisting of muscle- and bone-strengthening game-based activities. The resistance part included exercises involving large-muscle-groups for which therabands, fitballs as well as participant's own bodyweight were used; and 4) a 5-10 min cool-down part consisting of stretching and relaxation exercises. A typical session included five playground games/sports in the aerobic training part, including motor skills components (i.e., playful balance, coordination, hand-eye coordination, leg-arm coordination, spatial orientation, and reaction to moving objects or persons). In this regard, the space, number of collaborators/opponents, number of objects, size of the objects and signal-assigned movements were modified during each game. The main objective of the resistance training part was to strengthen the core, arms, and legs, which was done in a more analytic way than in the aerobic part. Five to ten exercises focused on the pushing, pulling, and throwing patterns were used.

*Monitoring the intensity of the exercise program*

The intensity of the exercise program was monitored in all children across the whole exercise program. Participants' progress relative to exercise intensity was checked weekly by trained personnel to: (i) adapt the intensity of the program progressively according to the improvements of the participants; and (ii) to identify whether any child was training at lower intensities than expected, thus requiring higher motivation during the exercise sessions. The heart rate (HR) data were recorded during both the aerobic and the resistance training components. Whereas HR is the most common indicator of the intensity of aerobic training, it is not commonly used as indicator of intensity in resistance training, since it does not directly reflect the stimulus at a muscular level. However, HR is informative of the exercise stimulus at the heart level in any type of exercise, including resistance training, and therefore, when interpreting the intensity of the whole program at a cardiorespiratory level, we used the HR data of the whole session, i.e., aerobic plus resistance training parts. Every child always wore the same HR monitor (POLAR

RS300X, Polar Electro Oy Inc., Kempele, Finland) that was individually programmed based on their maximum HR previously achieved in the maximal incremental test (see CRF measurement description below). Moreover, we also programed the HR monitors individually at 80% of the maximal HR and at the level of the anaerobic threshold (determined during the CRF test, see description below), so that we could later obtain the accumulated time over the 80% of the maximum HR and over the anaerobic threshold. These variables provide additional important information, since two different exercise programs could lead to the same average HR of the session, but have totally different amount of time accumulated at high intensities.

*Adverse events linked to the exercise program*

Only a single adverse event occurred during the intervention. One participant had an ankle bone fracture while using a static bike during one of the exercise sessions.

***Control group condition***

Children randomized into the control group were advised to continue with their usual daily routines, yet we provided them with a pamphlet including nutritional and physical activity recommendations. At the time when our study was conducted, Spanish schools typically included 2 lessons of physical education per week of roughly 50-55min, from which it is estimated that only 40-45% of the time in the physical education lessons was actually devoted to moderate-to-vigorous physical activity[10,11]. All of the children in the control group received the exercise program after the trial was completed.

***Outcome measurements***

Every measurement was carried out by the same trained evaluators. Demographic information was provided by self-report questionnaires.

*Intelligence*

Intelligence was assessed by the Spanish version of the Kaufman Brief Intelligence Test (K-BIT)[12], that was individually administered to each child[12]. This test consists of vocabulary and matrices subtests. The vocabulary subtest provides an estimated crystallized intelligence score and the matrices subtest provides an estimated fluid intelligence score. We used the age-specific percentiles for both crystallized and fluid scores, and total intelligence was calculated from their sum[12].

*Executive function*

The three core-dimensions of executive function were evaluated in this study: cognitive flexibility, inhibition, and working memory[13]. A full description of cognitive flexibility, inhibition, and working memory tests can be found elsewhere[14,15].

Cognitive flexibility and inhibition were assessed through three different sub-scales from the Delis–Kaplan Executive Function System (D–KEFS) whose reliability has been demonstrated elsewhere[16–18]. Cognitive flexibility was assessed using the Design Fluency Test and the Trail Making Test. The Design Fluency Test comprised three conditions: filled dots, empty dots, and switching. Participants were instructed to connect the dots using only four straight lines to design as many novel shapes as possible during 60 seconds for each condition. The total number of correctly drawn designs from all three conditions was recorded. A higher number of correctly drawn designs refers to better cognitive flexibility performance. The Trail Making Test comprised five different conditions. We used condition 2 and condition 4, known as Part A and Part B, respectively. In Part A (i.e., Number Sequencing), participants had to draw lines to connect numbers 1–25 in ascending order as fast as possible. In Part B (i.e., Number-

Letter Switching), participants had to draw a line to connect the numbers numerically and the letters alphabetically as fast as possible, switching each time from a number to a letter in consecutive order (e.g., 1–A–2–B, and so on). Part A had a maximum completion time of 2.5 min and Part B of 4 min. In cases where a child exceeded the maximum time, the test was stopped. The total completion time of Part B was subtracted from the total completion time of Part A as an indicator of cognitive flexibility[19]. A smaller B – A difference (sec) indicated better cognitive flexibility.

Inhibition was measured via a modified version of the Stroop test (paper-pencil version), which included four different conditions. We used condition 1 and condition 3. Condition 1 consisted of naming the color of filled rectangles. In condition 3, color-words were printed in a color that differs from their meaning (e.g., the word "red" printed in green ink) and the task consisted of naming the color of the word (i.e., green is the correct answer in the above example) and to avoid reading the word. An inhibition score was calculated as: condition 3 completion time minus condition 1 completion time (sec)[20]. The smaller the difference between test times (sec), the better the performance.

Working memory was measured by a modified version of the Delayed Non-Match-to-Sample (DNMS) computerized task[21]. A total of 16 practice trials followed by 140 experimental trials in 5 different blocks were presented focally on a computer screen using E-Prime software (Psychology Software Tools, Pittsburgh, PA). Each trial consisted of two phases (i.e., sample and choice) and two memory loads (i.e., high and low). For the present study, the high working memory load was used (i.e., 100 trials). The pre-target phase included a memory set of four different sequential stimuli (i.e., Pokemon cartoons) and participants were asked to memorize them. After the last stimulus, a target consisting of two different Pokemon was shown during the choice phase and participants were asked to select the cartoon that had not been previously shown. Response accuracy (%) in the high load was used as an indicator of working memory. Higher response accuracy refers to better working memory performance.

The paper-pencil based tests were given altogether in a session of 40-50 min. All of them were scored and automatically entered into a database by two separate investigators, and cases of disagreement were pursued until an agreement was reached. The DNMS working memory computerized task was given in a separate assessment that lasted 45-50 min.

*Academic performance*

Academic performance was assessed by the Spanish version of the Woodcock-Johnson III Tests of Achievement, which is a valid measure of academic performance[22]. A total of 12 tests were individually administered by a trained evaluator in a session of 100-120 min. Of these 12 tests, there were 3 tests of reading, 3 tests of mathematics, 2 tests of oral language, 3 tests of written language and 1 test of social sciences and humanities. All tests were double-corrected, processed in the Compuscore and profile software version 3.1 (Riverside Publishing Company, Itasca, IL, USA), and standard scores of reading, mathematics, writing, academic skills (i.e., the sum of tests based on basic skills such as reading decoding, mathematics calculation and spelling), academic fluency (i.e., a sum score indicating how fast the reading, calculation and writing task were performed), problem solving (i.e., the sum of tests based on solving academic problems in reading, mathematics and writing) and total performance (i.e., the overall measure of academic performance based on reading, mathematics and writing) were obtained. More information on the sub-components is available at: https://n9.cl/zsikj

*Brain magnetic resonance imaging (MRI) outcomes*

MRI acquisition for the structural and functional brain outcomes is collectively described, and the specific processing steps for each analysis are individually detailed below.

*MRI data acquisition*

All images were collected on a 3.0 Tesla Siemens Magnetom Tim Trio scanner (Siemens Medical Solutions, Erlangen, Germany) with a 32-channel head coil. High-resolution, T1-weighted images were acquired using a 3D MPRAGE (magnetization-prepared rapid gradient-echo) protocol. The acquisition parameters were the following: repetition time (TR) = 2,300 ms; echo time (TE) = 3.1 ms; inversion time (TI) = 900 ms; flip angle = 9º; field of view (FOV) = 256 x 256; acquisition matrix = 320 x 320, 208 slices; resolution = 0.8 x 0.8 x 0.8 mm; and scan duration = 6 min and 34s[23,24]. The resting-state functional MRI (rsfMRI) data consisted of a series of 160 scans acquired using a Gradient Echo Pulse Sequence while participants rested with eyes closed. The parameters were as follows: TR = 2000 ms, TE = 25 ms, flip angle = 80º, FOV = 240 mm, acquisition matrix= 240 x 240, 35 slices, resolution = 3.5 x 3.5 x 3.5 mm, and scan duration of 5 min and 26s.

*MRI data processing*

Volume and shape of hippocampus and other subcortical structures:

Volume and shape of the subcortical structures were extracted using the FMRIB's Integrated Registration and Segmentation Tool (FIRST), a semi-automated model-based segmentation tool in FMRIB's Software Library (FSL) version 5.0.7. FIRST uses Bayesian framework from shape and appearance models obtained from manually segmented images from the Center for Morphometric Analysis, Massachusetts General Hospital, Boston, MA, USA[25]. Briefly, FIRST runs a two-stage affine registration to a standard space template (i.e., Montreal Neurological Institute space) using 12 degrees of freedom and uses a subcortical mask to exclude voxels outside subcortical regions. Second, the hippocampus and other subcortical regions (i.e., 6 per hemisphere: nucleus accumbens, amygdala, caudate, globus pallidum, putamen, and thalamus; and brain stem) are segmented for both hemispheres separately.

Shape analysis is based on the individual meshes composed by a large number of vertices and triangles. The number of triangles and vertices is the same for each nuclei, allowing within- and between- participants comparison of each vertex. The vertex-wise analysis followed methods described elsewhere[26,27]. To assess local changes in each nuclei, we used the radial distance of each vertex to the medial curve of the nuclei (the centroid curve of the nuclei boundary in each section). Regional expansions and contractions (i.e., radial distances related each vertex spatial location to the core line of the structure) of the nuclei are the indicators of local changes in the structure shape[28]. Manual volumetric region labels are parameterized as surface meshes and modeled as a point distribution model. We extracted the volume in $mm^3$ of the hippocampus, including in the main analysis, and the rest of the subcortical regions.

In addition, the hippocampus segmentation from FIRST was then split based on the center of gravity of the region into anterior and posterior sub-regions for each hemisphere separately. This resulted in separate anterior and posterior hippocampal segmentations for each participant, for each hemisphere[29,30]. The final segmentations were visually inspected for quality. Each segmentation was used in the shape analysis and its volume was obtained from FIRST in $mm^3$.

Prefrontal cortex thickness, area and volume analysis:

Detailed information about pre-processing steps is discussed elsewhere [24]. Briefly, MPRAGE (Magnetization Prepared - RApid Gradient Echo) images were analyzed with FreeSurfer software version 5.3.0 (http://surfer.nmr.mgh.harvard.edu) using the standard processing pipeline known as recon-all that has been previously described and well-validated to assess cortical thickness (mm), surface area ($mm^2$) and volume ($mm^3$)[31–33]. Each time point was processed individually. Preprocessing steps included (i) skull-stripping, (ii) automated Talairach registration, (iii) gray/white matter segmentation, (iv) construction of a model gray-white matter boundary, and (v) cerebral cortex parcellation into region of interest based on gyral and sulcal structures from the Destrieux atlas[31–34]. Our *a priori* hypothesis included the prefrontal cortex sub-regions (i.e., 6 per hemisphere: cingulate gyrus, anterior division; inferior frontal gyrus, pars opercularis; inferior frontal gyrus, pars triangularis; middle frontal gyrus; superior frontal gyrus and frontal orbital cortex), then we used the extracted thickness, area, and volume of each sub-region as outcome in the group-level analysis.

Functional connectivity analysis between hippocampus and prefrontal cortex:

Detailed information about pre-processing steps is discussed elsewhere[35]. In sum, the following steps were carried out in FMRIB's Software Library (FSL) version 5.0.7: (i) skull-stripping, (ii) spatial normalization of structural, (iii) alignment of all rsfMRI frames, (iv) co-registration to each participant's structural image and spatial normalization, (v) the rsfMRI time courses were then band-pass filtered (0.1–0.01 Hz), (vi) six affine transformation parameters from the alignment process, as well as the mean time courses from the brain parenchyma including white matter tissue and ventricles were included as covariates. The residualized parameter estimate maps were converted to z scores (via Fishers r to z transformation) to achieve normality and were entered into higher level analyses. For hippocampal connectivity, we used the anterior and posterior sub-regions for each hemisphere separately (detailed above), as seeds.

Voxel-wise whole-brain volumetric analyses:

Detailed information about pre-processing steps is described elsewhere[23]. SPM software (SPM 12; Wellcome Department of Cognitive Neurology, London, UK) implemented in MATLAB (MathWorks, Inc., Natick, MA) was used for the whole-brain volumetric analyses. Imaging pre-processing steps included quality control and alignment, segmentation into gray matter tissue, white matter tissue and cerebrospinal fluid. Then, gray and white matter images were spatially normalized to MNI space and used to create a template using DARTEL. Subsequently, images were normalized to the DARTEL template via non-linear transformation, and modulated with Jacobian determinants. Finally, the images were smoothed by convolving them with an isotropic Gaussian kernel of 8 mm full-width at half-maximum (FWHM).

Total gray and white matter volumes were derived from segmented images, and total brain volume was calculated by adding the volumes of gray and white matterStructural covariance network analysis via Non-Negative Matrix Factorization:

Non-Negative Matrix Factorization (NNMF) analysis was used to identify structural networks. NNMF is a method for extracting structural networks where volume covaries across all participants[36]. An extended function of NNMF was used corresponding to the orthonormal projective non-negative matrix factorization (OPNMF), which was run using "opnmf_mem" in MATLAB with code available in https://github.com/asotiras/brainparts. This approach provided components that could be considered as a biologically more meaningful parts-based representation of the brain as compared to more standard approaches such as principal component analysis (PCA) or independent component analysis (ICA)[36,37]. Smoothed structural gray matter images (all processing information can be found in detail elsewhere[23]) for each subject were reshaped into a matrix including all available pre- and post- images for a high-quality accuracy of the structural networks (dimensions: 198 participants x 2122945 voxels). The local gray matter volumes with a threshold of 0.2 (i.e., to eliminate the voxels with partial volume effect) were then extracted in a whole-brain gray matter mask and used as input for OPNMF (dimensions: 198 participants x 470556 voxels) and approximates this matrix as a product of two matrices with non-negative elements.

The data were represented denoting the corresponding sparse components (W) and the subject specific loading coefficients (H). The first matrix, W, is of size V × K and contains the estimated non-negative networks and their respective loadings on each of the V voxels; and K is the specified number of networks. The W matrix, or "Network Components," is composed of coefficients that denote the relative contribution of each voxel in the network. The second matrix, H, is of size K × N and contains subject-specific loading coefficients for each network. These subject specific coefficients indicate the contribution of each network in reconstructing the original gray matter map. To obtain a range of possible solutions for comparison[36,38], we ran multiple NMF solutions requesting a K from 6 to 24 networks in steps of two (i.e., K=6:2:24). We then calculated the reconstruction error for each solution as the Frobenius norm and plotted the reconstruction error for all solutions. The solution resulted in 20 networks shown in Fig. 5 and Supplementary Table 16.

*Body composition*

Body weight was measured with an electronic scale (SECA 861, Hamburg, Germany) and height (cm) with a stadiometer (SECA 225, Hamburg, Germany). Both measurements were performed twice while participants stood barefoot, wearing light underclothes, with the averages recorded. Body mass index (BMI) was calculated as weight in kilograms divided by height in meters squared ($kg/m^2$). Children were categorized into weight status (to check the inclusion criteria of overweight/obesity) according to age- and sex-specific BMI cutoff points[2,3].

*Cardiorespiratory fitness*

CRF was evaluated under laboratory conditions using a gas analyzer (General Electric Corporation) while performing a maximal incremental treadmill (hp-cosmos ergometer, Munich, Germany) test modified for unfit children[39]. Both the gas analyzer as well as the flow-meter were calibrated before each assessment day according to the manufacturer's instructions. Briefly, participants walked on a treadmill at a constant speed (4.8 km/h) beginning at 6% slope with grade increments of 1% every minute until volitional exhaustion. Participants received strong encouragement during the test to continue the test as long as possible. As in previous major RCT conducted in children[39], not all children attained the maximal oxygen consumption ($VO_2max$), and we therefore used peak oxygen consumption ($VO_2peak$, mL/kg/min) and final completion time of the test (min), namely time-to-exhaustion, in the analyses.

*Biological maturation*

Peak height velocity (PHV) is a common indicator of maturity in children and adolescents[40]. PHV was calculated from age and anthropometric variables following Moore's equations[41]. Years from PHV were calculated by subtracting the age of PHV from chronological age, so that it is interpreted as how many years from the PHV offset a person is, with a value ranging from negative values (before the PHV; less mature) to positive values (after the PHV; more mature).

*Socioeconomic status*

Parents self-reported their higher education attained and current occupation, as described elsewhere[1,42]. A questionnaire about the highest educational level achieved was completed by both parents. We computed a dichotomized parental combined variable for the educational level as low (neither parent had university education) and high (at least one of the parents had university education), which was used in the moderation analyses. In addition, both parents were asked to answer an open question concerning their occupation at baseline. The answers of each parent were categorized following the International Standard Classification of Occupations (ISCO) and taking into account the Homemakers, and Unemployed, resulting in a variable ranging from 1 to 12[42]. The ISCO categories were re-categorized as high (1 to 3), medium (4 to 8), and low (9 to 12) for each parent. We computed a dichotomized parental combined variable for the occupational level as low (neither parent had a high occupational level) and high (at least one of the parents had a high occupational level), to be used in the moderation analyses.

***Overall activity assessment before and during the intervention***

Since previous literature indicates that a compensatory effect in the exercise group and a contamination effect in the control group might occur in overall physical activity levels in participants enrolled in intervention studies[43,44], we determined the changes in overall activity levels in children from both groups from baseline to during the intervention (i.e., in the middle of the intervention, week 10). For this purpose, overall activity patterns at baseline and during the intervention were assessed with accelerometers (GT3X+, ActiGraph, Pensacola, FL, US). The accelerometer data were processed as described elsewhere[45] in accordance with the systematic review on data processing recently conducted by our group[46]. Participants were asked to wear the accelerometer on the right hip and the non-dominant wrist for 7 consecutive days (24 h). A minimum of 4 valid days (i.e., ≥16 h/day), including at least 1 weekend day, was required to be included in the analyses. For the present study, the ENMO (Euclidean Norm Minus One) with negative values rounded to zero of the raw accelerations from the right hip was used as an indicator of overall activity[47,48]. Similar procedures were performed over the accelerations from the non-dominant wrist and used in sensitivity analyses. Accelerometer data processing was performed with the GGIR package[49].

***Statistical analysis***

*Effects of the ActiveBrains exercise program*

The effects on the outcomes were tested according to the per-protocol principle with analysis of covariance (ANCOVA) using post-exercise outcomes as dependent variables, group (i.e., exercise vs. control) as a fixed factor, and baseline outcomes as a covariate. Raw scores from each outcome were first winsorized (when needed) to limit the influence of extreme values; this method consists of replacing extreme high/low values for the closest (highest/lowest) valid value[50]. The z-scores for each outcome at post-exercise program were also formed by dividing the difference of the post-intervention raw score of each participant from the baseline mean by the baseline standard deviation (i.e., [post-exercise individual raw value – baseline mean] / baseline SD). This way of reporting the effects has been used in recent major RCTs focused on cognitive outcomes[50] and has two main advantages: 1) provides standardized estimates that allow comparisons across outcomes with different original measurement units, which are often abstract and not-intuitive in cognitive testing; and 2) these z-scores of change can be interpreted as effect size indicators, e.g., 0.5 z-score means that the mean value at post-exercise program is 0.5 SDs higher than the mean value at baseline, indicating a positive medium-sized change, with negative values indicating the opposite. As effect size indicators, they can be interpreted according to the standard benchmarks, i.e. a value around 0.2 is considered a small effect size, 0.5 is considered a medium effect size and 0.8 is considered a large effect size[51].

Two of the executive function tests, i.e., the cognitive flexibility test 2 (Trail Making Test) and the inhibition test (Stroop Color-Word Test), are originally expressed in an inverse way which means that lower scores indicate better performance. To simplify the visual interpretation of the main findings presented in **Figure 3**, we inverted these two scores so that they can be interpreted in the same fashion as the rest of the outcomes (i.e., higher score better performance), as indicated in the figure legend. However, we kept the original scores and units of measure in **Supplementary eTable 2** and information on how to interpret each test is provided as footnotes. Since we have assessed cognitive flexibility using 2 different tests, we analyzed them separately, as well as we computed a composite z-score for cognitive flexibility (i.e., the Design Fluency Test and Trail Making Test). Likewise, we analyzed the three core executive functions (i.e. cognitive flexibility, inhibition, and working memory) separately as well as combined into a composite score of executive function[52]. The composite scores were computed by averaging the z-scores for their individual components and renormalizing the average of z-scores to have a mean of 0 and a SD of ±1 at baseline. Prior to the computation of the averages, the Trail Making Test and the Stroop Color-Word Test were inverted by multiplying by -1, so that the interpretation of the score would go in line with the rest of tests (higher score, better performance) and could therefore be averaged with the rest of the variables.

For certain MRI outcomes (i.e., volume of hippocampus and other subcortical structures, volume, thickness and surface area of prefrontal cortex subregions and its asymmetric scores, and total brain volumes), the same ANCOVA models were run in SPSS, i.e., post-exercise values were entered into the models as dependent variables, group (i.e., exercise vs. control) as a fixed factor, and baseline values as covariates. Other analyses conducted directly on images and using specific MRI tools for analysis need specific explanations.

*Specific notes on functional connectivity analyses*

Voxel-wise functional connectivity network maps were constructed for each hippocampal seed, for each participant using the pre-processed rsfMRI data. As our hypothesis was focused on the hippocampal functional connectivity with the prefrontal cortex, we selected the 12 prefrontal cortex sub-regions (i.e., 6 per hemisphere: cingulate gyrus, anterior division; inferior frontal gyrus, pars opercularis; inferior frontal gyrus, pars triangularis; middle frontal gyrus; superior frontal gyrus and frontal orbital cortex) for this analysis. Then, we extracted from the first-level seed maps the percentage signal change for each prefrontal cortex sub-region, using the Harvard-Oxford Cortical Structural Atlas, with each hippocampal seed. Finally, the group-level statistical analysis was performed following the per-protocol method with ANCOVA using post-exercise program percentage signal change as dependent variables, group (i.e., exercise vs. control) as a fixed factor, and baseline percentage signal change as covariates.

*Specific notes on shape of subcortical structures' analyses*

To examine the effect of the exercise intervention, FSLmaths (https://fsl.fmrib.ox.ac.uk/fsl/fslwiki/Fslutils) was used for subtracting the shapes of the post- minus pre- intervention. Registrations and all segmented outputs for location and quality of the segmentation for all structures of each child were double-checked. The resulting (post-pre) image was used for the analysis as a dependent variable, and differences between groups were tested.

*Specific notes on voxel-wise whole-brain volumetric analyses*

To investigate the potential differences on gray and white matter volume without anatomical *a priori* predictions, we performed voxel-wise whole-brain volumetric analyses on smoothed images. We used the General Lineal Model approach implemented in SPM12. We used a full factorial design in two-way ANOVAs (2x2). Design specification proceeds in 2 stages. Firstly, we created two factors (group and time) and two levels per factor (i.e., exercise vs. control for group factor, and pre-intervention vs. post-intervention for time factor). Secondly, we separately assigned the images to each cell in the following order: pre-exercise, pre-control, post-exercise, post-control. We then created the positive and negative contrasts for time (i.e., c1=[1 0 -1 0 ], c2=[-1 0 1 0], c3=[0 1 0 -1], c4=[0 -1 0 1]), group (i.e., c5=[1 -1 0 0], c6=[-1 1 0 0], c7=[0 0 1 -1], c8=[0 0 -1 1]), and time*group (i.e., c9=[-1 1 -1], c10=[1 -1 -1 1]). A FDR (false discovery rate)-corrected $p< 0.05$ was initially set for the analyses, and for exploratory purposes, an uncorrected voxel-level threshold of $p < 0.001$ (and also $p < 0.05$) without any extent threshold was chosen.

*Specific notes on structural covariance network analysis*

The group level statistical analysis was performed following the per-protocol method including those participants with available data at both time-points. Each participant specific loading coefficient (H) for each network was used as outcome and included in the ANCOVA using post-exercise program voxel-data as dependent variables, group (i.e., exercise vs. control) as a fixed factor, and baseline voxel-data as covariates.

*Mediation analyses*

We tested whether the effects of the intervention on the main study outcomes were mediated by changes in CRF following the bootstrapping method[53]. Our mediation analyses are in line with the AGReMA (A Guideline for Reporting Mediation Analyses: https://agrema-statement.org/) statement and the corresponding checklist is provided as **Supplement 3**. Mediation analyses were performed using the PROCESS macro for SPSS (SPSS Inc., Chicago, Illinois) with a resample procedure of 5,000 bootstrap samples. These mediation analyses were performed for the outcomes for which significant differences were observed between exercise and control groups in main effect analyses. The unstandardized (B) and standardized ($\beta$) regression coefficients are presented for four equations: Equation 1 regressed the mediator (e.g., change in CRF) on the independent variable (group). Equation 2 regressed the dependent variables (i.e., executive function or academic performance outcomes) on the independent variable. Equation 3 regressed the dependent variables on both the mediator (equation 3) and the independent variable (equation 3'). We also included the outcome of interest at baseline as a confounder. The indirect effects along with its confidence intervals (CIs) were also presented and the significance was considered if the indirect effect significantly differed from zero (i.e., zero was not contained within the CIs). Finally, the percentage of the total effect was computed to know how much of the total effect was explained by the mediation, as follows: (indirect effect / total effect) × 100. This mediation analysis was performed using the variable time-to-exhaustion during the treadmill test (min) as a mediator variable since larger effects of the exercise program were observed on this CRF outcome. The analyses were also replicated using $VO_2$peak (mL/kg/min) as a mediator variable. The same modeling was applied to test whether the effects observed on academic performance outcomes were mediated by exercise-induced changes in executive function or intelligence outcomes.

*Moderation analyses*

To explore whether the effects of the intervention were modified by potential moderators, we ran the same models as for the main effects' analyses but stratifying the analyses by sub-groups of population according to sex (boys *vs*. girls), age (8-9 *vs*. 10-11 years of age), biological maturation (below and above the median of PHV), parental educational level (low *vs*. high), parental occupational level (low *vs*. high), and baseline outcome levels (below and above the median). In a first step, visual inspection of the effects sizes by subgroups shown in plots was used to identify the main potential moderators. In a second step, we run ANCOVA models to test whether the interaction term (e.g. sex × group, age × group, etc.) in those cases were significant.

*Intention-to-treat and dropout analyses*

It is a standard requirement to report effects derived from both per-protocol and intention-to-treat analyses. For the intention-to-treat analysis, we performed multiple imputation to predict missing values at post-exercise outcomes using the predictive mean matching approach. We performed 10 iterations to create 5 databases, which were then averaged to obtain the imputed values for the intention-to-treat analyses [54]. Once we had a dataset with imputed data when missing for the 109 participants initially allocated into the study groups, we ran the same models to test the effects of the intervention as described above. The effects on the primary outcomes are presented in Supplementary eTables.

In addition, we used a one-way ANOVA to test whether the participants that completed the baseline evaluations and randomization, but left the study during the intervention period or did not complete the post-exercise evaluations (i.e., namely the dropouts), differed in the main study outcomes at baseline from the participants who completed the study and post-exercise evaluations (i.e., namely the non-dropouts).

*Testing potential compensatory and contamination effects of the intervention on overall activity levels, and analyses on the intensity of the training sessions*

We performed a 1-dimension curve analysis using SPM1D ((one-dimensional Statistical Parametric Mapping) package available for MATLAB (http://www.spm1d.org)[55] to study whether acceleration values (i.e., expressed as ENMO [m*g*]) identified a significant increase in physical activity during the exercise program in comparison with the physical activity pattern at baseline for the control and exercise groups. SPM1D is a statistical parameter mapping tool using random field theory and can perform conventional statistics on 1-dimensional data, as is the case of the waveform acceleration data. Weekly average acceleration curves were presented separately for exercise and control groups from midnight (i.e., 00:00 AM) to next midnight, i.e., 24 h curves centered at noon (12:00 PM). Paired t-tests over the curves were used to identify significant differences between baseline and exercise patterns for each group throughout the day. SPM involved four steps to compute the t-test analysis[56]: 1) computing the value of a test statistic at each point in the normalized time series; 2) estimating temporal smoothness on the basis of the average temporal gradient; 3) computing the value of test statistic above which only $\alpha$ = 5% of the data would be expected to reach had the test statistic trajectory from an equally smooth random process; 4) computing the probability that specific supra threshold regions could have resulted from an equivalently smooth random process. Finally, we tested sex differences in the intensity achieved during exercise sessions using one-way ANOVA with different HR outcomes in separate models and sex as fixed factor.

### EXTENDED RESULTS

***1. A priori-planned analyses. Effects of the exercise intervention on primary outcomes: intelligence, executive functions (i.e., cognitive flexibility, inhibition and working memory), academic performance and hippocampal volume.***

The results of this section are all presented in the main text.

***2. Extended results on a posteriori-planned analyses on brain MRI outcomes***

*2.1. Analyses on selected brain regions of interest*

2.1.1. Hippocampal subregions (structural MRI volumes)

In a recent observational study, our group found that the activity of certain subregions of the hippocampus (particularly the anterior hippocampus) were more associated with exercise-related factors such as CRF than other hippocampal subregions[57]. Likewise, an aerobic exercise intervention in young- to middle-aged adults observed an increase in anterior hippocampal volume only, not in the posterior subregion[58]. Another aerobic exercise intervention in older adults consistently observed larger effects of exercise in the anterior compared to posterior hippocampus[30], supporting the notion that the anterior hippocampus is selectively more amenable to exercise intervention. Therefore, we tested the effects of our exercise program on the different hippocampus subregions separately (**eTable 4 in Supplement 1**), and observed no effect on any of the hippocampal subregions studied, i.e., the anterior or posterior regions from the right or left hemispheres (all effects sizes <0.1 SDs, P values >0.5).

2.1.2. Prefrontal cortex subregions (structural MRI analyses on gray matter volume, cortical thickness and cortical surface area)

Since the largest effects observed in this trial were on intelligence and cognitive flexibility, and one of the brain regions known to be related to both intelligence and cognitive flexibility is the prefrontal cortex[59–61], we investigated whether our exercise intervention had an effect on gray matter in prefrontal cortex subregions. For this purpose, we analyzed the effects of the intervention at three structural levels of the prefrontal cortex: gray matter volume (**eTable 5 in Supplement 1**), cortical thickness (**eTable 6 in Supplement 1**) and cortical surface area (**eTable 7 in Supplement 1**). We observed no significant effect of exercise on any of these outcomes (all effects sizes $\leq |0.29|$ SDs, P values >0.05).

2.1.3. Functional connectivity between hippocampal and prefrontal cortex subregions (functional MRI analyses)

As discussed above, the two primary regions of interest in our study were the hippocampus (*a priori*-planned) and prefrontal cortex (*a posteriori*-planned). We therefore analyzed the exercise effects on the brain not only at a structural level (see Results' sections above, and **eTables 4-7 in Supplement 1**), but also at a functional connectivity level between these two regions. Our findings did not support any exercise effect on functional connectivity between the hippocampal subregions and prefrontal cortex subregions (all effects sizes $\leq |0.55|$ SDs, P values >0.05; (**eTables 8-13 in Supplement 1**).

##### *2.2. Broader brain analyses*

2.2.1. Other subcortical regions

###### Gray matter volumes of subcortical brain structures

Subcortical brain structures are known to play an important role in cognition and executive function[62–64]. Therefore, in addition to the hippocampus, we tested whether our exercise program had an impact on the gray matter volume of all subcortical structures and observed no significant effects (**eTable 14 in Supplement 1,** all effects sizes ≤ │0.16│SDs, P values ≥ 0.1).

###### Morphologic (shape) analysis of subcortical brain structures

A novel and highly sensitive means to examine potential brain changes is through shape analysis of the subcortical structures. The whole volume of a structure might remain unchanged, whereas some parts of the structure might expand while others contract, and these morphological changes are indicative of neural development[65] and are related to cognitive performance[26,66–68]. In a cross-sectional study, our group reported expansions in different subcortical structures, including the hippocampus, in relation to higher levels of CRF in children[28]. However, whether an exercise intervention is able to change the shape of subcortical structures at any age is still unknown. We investigated this research question for all main subcortical brain structures (15 nuclei): brain stem, and left and right accumbens, amygdala, caudate, hippocampus, pallidum, putamen, and thalamus. Consistent with the results for the volume of subcortical structures reported above, we observed no effect of the exercise intervention on the shape of the hippocampus (as a whole as well as the anterior or posterior hippocampus) or any other subcortical structure. In **eFigure 2 in Supplement 1** we present a graphical illustration of the shape analysis based on radial distances in each structure, as well as the different subcortical structures studied.

2.2.2. Whole brain analyses

###### Total brain volumes

In cross-sectional studies, we have recently observed that higher fit children have bigger brains, i.e., greater total brain volume, total gray matter volume and total white matter volume[69]. Further, a meta-analysis concluded that existing literature supported a modest, but robust positive association between brain size and intelligence[70]. Since exercise is known to improve fitness[71–73], we tested whether our exercise intervention could have small cumulative effects on different regions of the brain and across the whole brain, which could result in increased total brain volume. However, our findings do not support this hypothesis as a null effect of our exercise program was observed on total brain volume, total gray matter volume and total white matter volume (**eTable 15 in Supplement 1,** all effects sizes ≤ │0.1│SDs and all P values ≥ 0.3).

###### Whole-brain voxel-wise volumetric analysis

Likewise, a voxel-wise whole brain volumetric approach was used as a hypothesis-free analysis, covering any potential effect of the exercise program on meaningful clusters in any brain region, yet no significant effect was observed in either gray matter or white matter volumes (data not shown). We have not identified previous exercise trials analyzing their effect on voxel-wise whole-brain volumes. Therefore, we can compare our findings only with previous cross-sectional studies conducted in children in which we identified a number of brain regions where CRF was associated with gray matter[23] and white matter[74] volumes.

###### Whole-brain structural covariance network analysis

Most of the structural brain analyses conducted in our study and in the literature are dependent on predefined brain regions, not only in the relation to exercise, but also in the neuroimaging field. Alternatively, it is of interest to also investigate the brain using non-predefined approaches, particularly in children since brain atlases were created using MRI images from samples of healthy adults. In this context, we applied the structural covariance network analysis delineated by Non-negative Matrix Factorization analysis, that has been recently linked to intelligence[75]. In **eFigure 3 in Supplement 1**, we present the 20-networks model generated and in **eTable 16 in Supplement 1** the exercise effects on these networks, which were all non-significant (all effect sizes ≤ │0.1│ SDs and all P values ≥ 0.1).

##### *3. Effects of the intervention on CRF and its role as mediator*

No additional results in this section.

##### *4. Extended results and interpretation about moderators of the intervention effects*

The strongest moderation effect was observed with sex, for which a significant interaction term (sex × group) was observed (P<0.05). The moderating effect of sex was independent of differences in maturation, known to exist between sexes at these ages, and *vice versa.* We explored potential reasons for why our exercise program had a different effect in boys and girls. Interestingly, we observed that although the average HR of the sessions did not differ between boys and girls, boys spent more time at high intensity zones as defined by time spent over their individualized anaerobic threshold (**eTable 18 in Supplement 1**). In this context, Castelli at al.[76] observed that only the time spent at high intensities (defined as time over the 80% of the individual maximal HR), and not the average HR of the sessions, predicted improvements in cognitive performance in children. These findings, together with ours, support the notion that more time spent at high intensities by boys in our program could, at least partially, explain the larger effects observed in certain outcomes in comparison with girls.

##### 5. Other exploratory analyses related to the interpretation of the intervention effects

*Intention-to-treat and dropout analyses*

Overall, the effects on primary outcomes derived from the intention-to-treat analyses were consistent with those from the per-protocol analyses described above, yet effect sizes were attenuated in most cases. This attenuation is to be expected since participants not included in per-protocol analyses but included in the intention-to-treat analyses attended fewer exercise sessions, which would support the expected effect that a higher exercise dose (attending to more sessions=higher total volume of training) could lead to larger effects. Nevertheless, the strongest effects observed in this trial, i.e., total intelligence, crystallized intelligence and cognitive flexibility, remained significant (effects sizes = 0.5 for intelligence outcomes and 0.3 SDs for cognitive flexibility, P values ranging from 0.019 to <0.00001) (**eTable 19 in Supplement 1**), supporting the robustness of the findings. The effects on academic performance were also attenuated becoming mostly borderline non-significant in the outcomes benefited under per-protocol analyses, i.e., total academic performance, mathematics, problem solving and academic skills (effect sizes=0.2, P values ranging from 0.049 to 0.1) (**eTable 20 in Supplement 1**). The effects on hippocampal volume remained non-significant in the intention-to-treat analyses (**eTable 21 in Supplement 1**).

Overall, our dropout analysis (**eTable 22 in Supplement 1**) showed that the participants who dropped out of the study did not significantly differ at baseline from those that completed the study (i.e., non-dropouts). This holds true for the main study variables, including CRF and all primary outcomes, except for hippocampal volume which was lower in the dropouts (P=0.02).

*Testing the potential compensatory and contamination effects of the intervention on overall activity levels*

We measured overall physical activity before (i.e., baseline) and during the intervention (i.e., week 10) with raw accelerations collected over a week with an ActiGraph GT3X+ monitor attached on the right hip and we averaged them into the metric ENMO (expressed in mg)[47,49]. The SPM1D analysis of the whole 24 h activity curve (mg) shows how the exercise group significantly increased their activity levels in the afternoon-evening (P<0.001) when the exercise program took place, and did not change their activity levels in the hours when not participating in the exercise program. On the other hand, the control group kept the same levels of activity (**Figure 5**) throughout the 24h period. Noteworthy is that the results were consistent when using the data derived from the wrist-attached instead of the hip-attached accelerometer (**eFigure 5 in Supplement 1)**, increasing the robustness of the findings. The application of the SPM1D analysis to test changes in overall activity as a result of an intervention in the study groups is novel and could be applied in future exercise-based RCT.

*Characterization of the actual volume and intensity of the exercise program*

All participants wore HR monitors throughout the program. Although the sessions lasted approximately 90 min, after subtracting the time spent on arrival and affixing the children with HR monitors, warming-up and cooling-down, the actual main part of the session was on average 66 min per session. The HR monitors were individually programmed according to participants' individual maximum HR reached during an incremental maximal test at baseline. We observed that the aerobic training part of the intervention sessions had an average duration of 46 min (SD=3 min), and an average intensity of 148 bpm (SD=8.8 bpm), which was equivalent to 75% (SD=3%) of their maximal HR. The resistance training part of the sessions lasted on average 20 min (SD=2.9 min), during which the children had an average HR of 127 bpm (SD=8 bpm), which was equivalent to 64% (SD=4%) of their maximum HR. This resulted in an average HR of 138 bpm (SD=8bpm) per session, aerobic plus resistance training, which means that the children trained for more than 1 h at 70% of their maximum HR. In addition, the children accumulated on average 38% of the session time (i.e., 25min) at high intensities over the 80% of their maximum HR (**eFigure 6 in Supplement 1**). Taking into account that the American College of Sports Medicine defines vigorous physical activity as ≥ 77% of the maximal HR[77], we can conclude that the children participating in the ActiveBrains trial trained at relatively high intensity for a long duration. As an additional feature of exercise volume received by our participants, we present in **eFigure 7 in Supplement 1** the distribution of the attendance to the exercise program.

#### EXTENDED DISCUSSION

##### Discussion of the findings in the context of previous studies

To the best of our knowledge, only three previous intervention studies tested the chronic effects of exercise on intelligence in a pediatric population. The first study tested the effects of a yoga program, but did not include a control group[78]. The second study, a cluster school-based RCT, investigated the effects of daily physical education sessions, yet half of the "control" group also received daily physical education for half of the intervention period[79]. The third study was a school-based pilot study conducted by our group in only 17-20 children per study group and investigated the effects of increasing the intensity and the number of physical education sessions per week[80]. The conclusions from these three studies suggested potential benefits of exercise on intelligence. Given the preliminary nature of these findings and limitations associated with the study design and sample size, the ActiveBrains RCT provides the strongest evidence thus far regarding a causal effect of physical exercise on intelligence, and particularly crystallized intelligence, denoted by a large effect (i.e., +9 points in the typical punctuation of the test, equivalent to 0.7SDs, larger improvements in the exercise group). Interestingly, although previous evidence for the chronic effects of exercise on intelligence is limited, more evidence is available for the acute effects of exercise[81]. The 2018 Physical Activity Guidelines Scientific Advisory Report concluded that there is evidence supporting an improvement in crystallized intelligence after a single bout of moderate-to-vigorous physical activity in children[81,82], which supports our findings.

Our exercise program demonstrated a medium-sized effect (i.e., 0.5 SDs) on cognitive flexibility, while null effects for the other executive functions tests, i.e., inhibition and working memory (i.e., 0.04 and 0.05 SDs respectively). Systematic reviews and meta-analyses in children and adolescents have reported a significant effect of exercise on overall executive function[83–87], with mixed conclusions among reviews when referring to the specific dimensions of this complex cognitive construct. The large diversity of cognitive tasks used and the different characteristics of the exercise interventions (i.e., mode, frequency, session's duration, intensity, intervention's length) across studies might explain the discrepancies among individual studies. However, cumulative evidence recently synthesized supports a positive effect of exercise on the three core executive functions: working memory, inhibition and cognitive flexibility[87]. Noteworthy, we observed a larger effect in one of the cognitive flexibility tests, the Design Fluency Test, compared to the other one used, the Trail Making Test. The differential effect observed between these two tests may be due to the different nature of the tests. Although typically classified as a cognitive flexibility test, the Design Fluency Test has a multifactorial nature, which includes not only cognitive flexibility but also other cognitive processes such as creativity[18]. This is interesting, since a recent comprehensive review has highlighted that cognitive flexibility and creativity actually share similar brain networks[88]. Concerning academic performance, our findings are in line with existing literature in which exercise has specifically improved mathematics to a higher extent than other academic subjects including language (reading, writing)[89,90]. A major contribution of our study is that the positive effect of exercise on mathematics is partly explained by exercise-induced improvements in fluid intelligence, and that the positive impact of exercise on total academic performance, as well as problem solving and academic skills, is partly mediated by exercise-induced improvements in cognitive flexibility. These findings support the notion that executive function plays an important role in academic performance[91–93] and contributes to the understanding of the cognitive processes by which exercise improves academic performance.

Our exercise program had no significant effects on any of the brain outcomes studied. The consistency of the non-significant findings despite the large number of tests performed increases the credibility and robustness of the no-effect conclusion, and raises a myriad of research questions concerning the dominant characteristics of an exercise trial needed to effectively impact brain outcomes. A first major question is whether our intervention was long enough to elicit structural changes in the brain. Erickson et al., in a sample of older adults, observed no significant effect of aerobic exercise on hippocampal volume at 6 months of intervention, but did so after 12 months of intervention[30]. These findings suggest that interventions lasting longer than 6 months might be required to have a positive impact on hippocampal volume, at least in a population of older adults. This is in agreement with the conclusion from a recent meta-analysis, based on evidence in older populations[94]. However, it is believed that the degree of structural neuroplasticity (which includes gray matter volume) in the developing brain is higher than in the adult or older adult brain[95]. Therefore, it is possible that environmental factors, such as exercise, during a sensitive period of life, could have a faster and/or a larger impact on brain structure compared to when applied later in life. This hypothesis seems to be partially supported by a study conducted in young to middle-aged adults, in which a relatively short aerobic exercise intervention of 6 weeks increased gray matter volume of the anterior hippocampus compared to 6 weeks of no exercise (N=54)[58]. Similarly, a 12-week aerobic exercise program increased hippocampal volume in pediatric brain tumor survivors (N=28)[96]. Another recent study conducted in young adults observed an improvement in gray matter volume of the left medial superior frontal gyrus as a result of a 9-week aerobic exercise intervention (N=120)[97]. On the other hand, our trial conducted during 20 weeks in children failed to support this hypothesis. Less statistical power is not likely the reason for the null effect in our trial compared to the significant effect observed in the two previous trials discussed, since our study involves the largest sample size (with 109 participants included in the intention-to-treat analyses and 82 in the per-protocol analyses) in a pediatric population and the second largest (after the study by Erickson et al. in older adults, N=120[30]) out of the previous 23 exercise-based RCTs focused on hippocampal volume included in the most recent meta-analysis[94]. In this context, we aimed to contribute to the existing knowledge investigating the effects of exercise on voxel-wise whole brain white or gray matter volume, cortical area or thickness, shape of subcortical structures, structural networks or resting-state hippocampal functional connectivity in a pediatric population. Other studies (4 trials conducted in US and 1 in Canada) conducted in children have examined other brain outcomes and observed positive effects of exercise on white matter integrity[96,98,99], task-based functional MRI[6,100,101], and resting-state synchrony[102]. We believe that some brain outcome must have changed in our participants in the exercise group to explain the observed changes in intelligence and cognitive flexibility. That change could have occurred at molecular or cellular level, or some other features that were detected not with the neuroimaging techniques used herein. The continuous advances in the neuroimaging field will open new avenues for the study of the brain in the future, and thus, of the effects of exercise on human brain.

In secondary analyses (*a posteriori*-planned), we investigated potential mediators and moderators of the effects of exercise on brain health outcomes. Whereas previous cross-sectional studies from our group and others have consistently reported that children with a higher CRF have a healthier brain, as indicated by brain structure and function outcomes[23,24,28,35,63,64,69] as well as behavioral outcomes[103–108], information from previous exercise trials focused on brain health outcomes formally testing the potential mediating role of CRF is scarce. In our study, we first tested the effects of the intervention on CRF indicators, and observed an effect on CRF performance (i.e., time-to-exhaustion) of 0.4 SDs larger improvements in exercise than in the control group. Similar results yet of smaller magnitude and non-significant were also observed in $VO_2$peak, with a 0.3 SDs larger improvement in the exercise than control group, which is equivalent to an improvement of 1.4 mL/kg/min. The effect size observed for $VO_2$peak in our trial is in line with that observed in previous studies, such as the FITKids trial, which reported an effect size of 0.3 SDs and 1.3 mL/kg/min[109]. Second, we ran the mediation models and observed that exercise effects on crystallized intelligence, total academic performance and problem solving were partly mediated by exercise-induced improvements in CRF. Of note, this mediation was of small magnitude (10-20%) and non-significant in outcomes such as cognitive flexibility and mathematics, which suggest that the benefits of physical exercise may occur, at least in part, independently of improvements in CRF (e.g. as a result of learning new perceptual-cognitive-motor skills in the games of the intervention). These findings have important implications from both a scientific and a public health standpoint, providing empirical support for the classical theoretical model proposed by Bouchard and Shephard[110]. In line with that model, our findings suggest that physical exercise can improve health, in this case brain health, both directly and indirectly through improvements in CRF. Our trial provides causal evidence that improvements in CRF explain, to a small extent, improvements in brain health. It could be hypothesized that the exposure to exercise from childhood to adulthood could result in progressively differentiated life trajectories on both CRF and brain health, and that in trials like ours we can only observe the beginning of such differentiation. Unfortunately, this is speculative, and exercise-based RCTs focused on brain health lasting longer than 2 years currently do not exist, and will be challenging (yet not impossible, see a recent 5-year exercise RCT[111]) to be conducted in the future.

Although our study was not powered to test significant effects in separate subgroups, we could investigate whether the effect sizes observed were consistent across potential moderators-based subgroups for exploratory purposes. Overall, the effects of exercise on most of the behavioral outcomes were rather consistent, except for some exceptions particularly for the intelligence outcomes. We observed a larger effect on crystalized intelligence in boys than in girls, which is in line with a recent meta-analysis by Ludyga et al.[112] This meta-analysis concluded that sex could be an important moderator and suggested that, among other factors, the larger effects in males could be due to greater engagement in the exercise intervention, yet evidence demonstrating this is lacking. Consonant with this hypothesis, we observed that the time spent in high intensity exercise, as defined by time over the individualized anaerobic threshold, was significantly larger in boys than in girls. This could partially explain the larger effects on crystalized intelligence observed in boys, since exercise at high intensity zones was related to larger effects of exercise on cognitive performance in a previous study in children[76]. However, it is still not clear whether exercise of high intensity produce larger benefits on cognition than that of moderate intensity, as was evidenced in the recent meta-analysis by Ludyga et al.[112]. In addition to sex, we observed that exercise had a larger effect on crystallized intelligence in the children younger than 10 years and in those less biologically mature, which might indicate a more sensitive period for stimulating intelligence. Moreover, we observed that children from families with a lower socioeconomic status, as assessed by lower parental educational and occupational level, show larger improvements in fluid and total intelligence than their peers with higher socioeconomic status. The large disparities in general health, but also in brain health, across different socio-economic status are well known, and the present findings support the notion that those disparities could be reduced, mitigated or even eliminated by engaging in exercise interventions targeting families with lower socio-economic status. If these findings are replicated and confirmed in future exercise RCTs, they can have important implications for health equity. Consonant with the previous finding, participants with lower performance on the intelligence test at baseline, improved more in fluid and total intelligence in response to exercise than those with higher performance at baseline. The larger response to exercise in children with lower baseline performance in the tests studied has been reported for executive function[113], which is in line with our results for intelligence outcomes, yet we did not observe this moderating effect for executive function outcomes in our study.

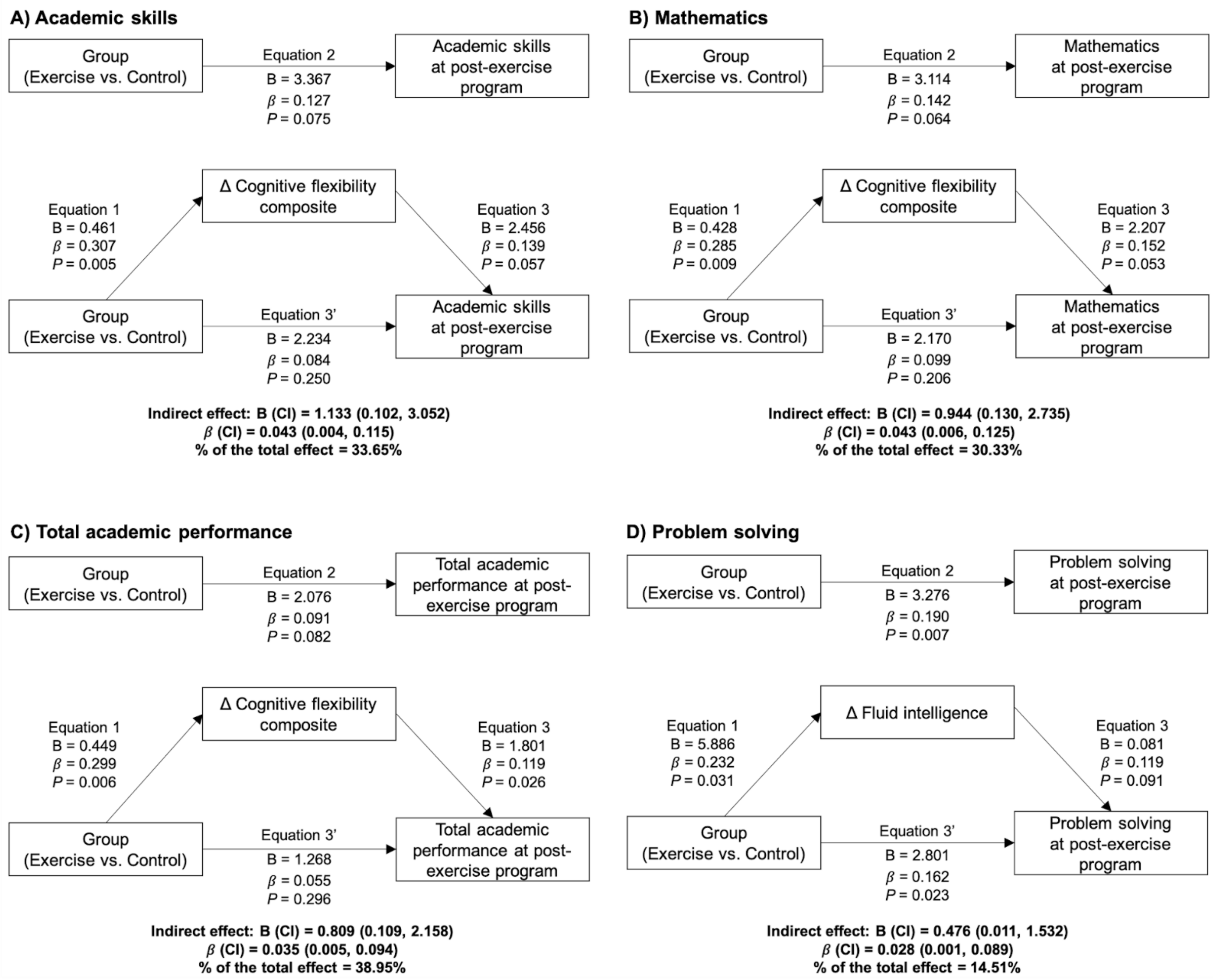


**eFigure 1. Cognitive flexibility and fluid intelligence mediation models of the intervention effects (i.e., exercise vs. control) on academic performance outcomes in children with overweight/obesity.**

Each analysis was adjusted by the respective academic performance outcomes at baseline. Bold font indicates significant indirect effect at $P<0.05$. B indicates unstandardized regression coefficient and $\beta$ indicates standardized regression coefficient.

Delta (Δ) cognitive flexibility and fluid intelligence express the change in total cognitive flexibility and fluid intelligence score at post-exercise program with respect to the score at baseline.

Cognitive flexibility composite z-score was calculated as the re-normalized mean of the z-scores for Design Fluency Test and Trail Making Test. Fluid intelligence was measured by the Kaufman Brief Intelligence Test.

Academic performance outcomes were obtained from the Spanish version of the Woodcock Johnson III Test of Achievement. Academic skills are the sum of components based on basic skills such as reading decoding, mathematics calculation, and spelling. Problem solving is the sum of the components based on solving academic problems in reading, mathematics, and writing. Total academic performance is the overall measure of the academic performance based on reading, mathematics, and writing.

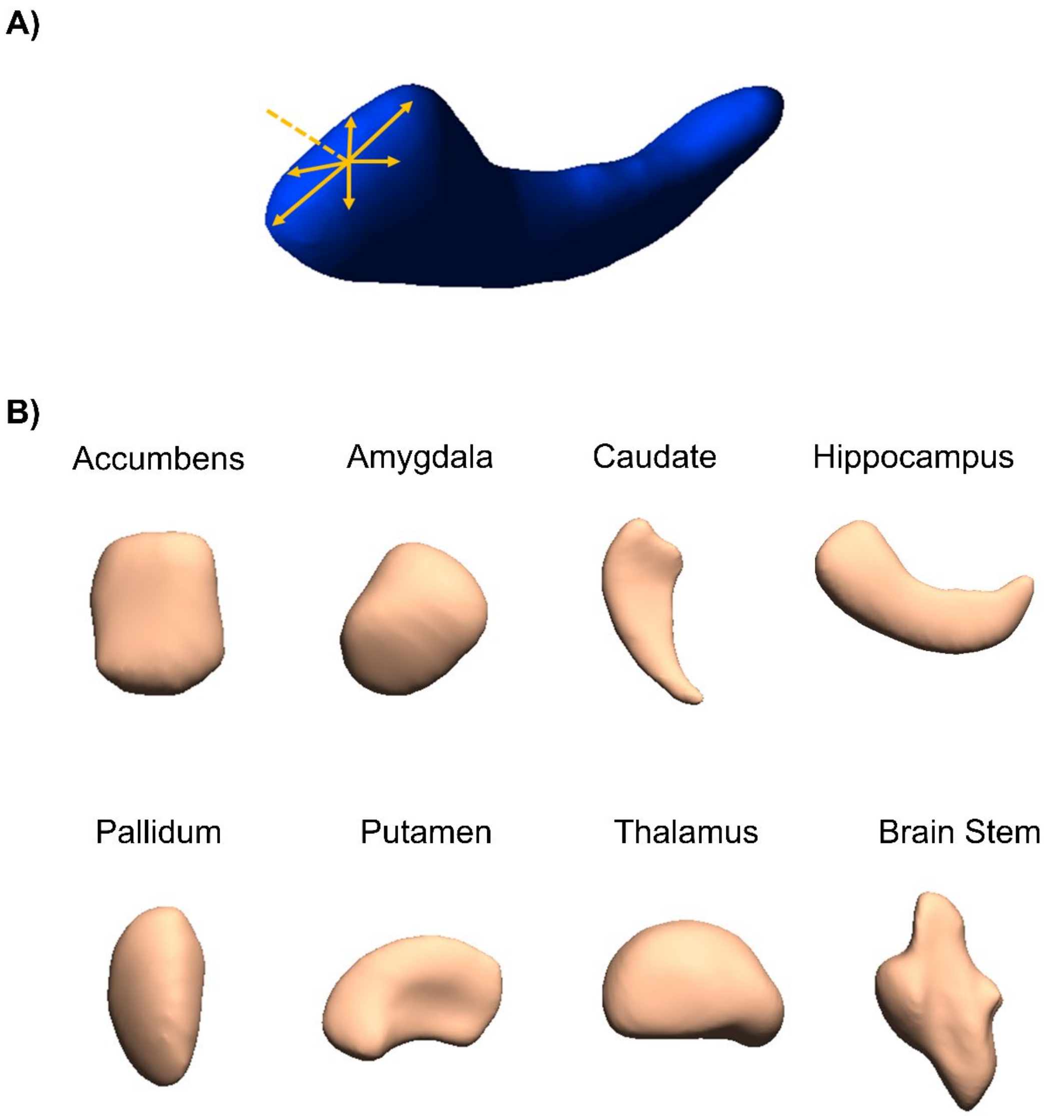


**eFigure 2. An illustration of the shape analysis of subcortical brain structures.**
Panel A, shows an illustration of the radial distances (orange arrows) to the media line (orange discontinuous line) in a section of the left hippocampus. The medial line is independent of the pose of the structures. Panel B, shows the subcortical brain structures analyzed (i.e., Accumbens, Amygdala, Caudate, Hippocampus, Pallidum, Putamen, Thalamus) and the Brain Stem. Structures from the left hemisphere are shown, as an example, yet both hemispheres were analyzed. The analysis of radial distances allows studying expansions (larger radial distance) or contractions (shorter radial distance) in the subcortical brain structures as an effect of the exercise intervention. We observed no significant effect of our exercise program on the shape of subcortical brain structures ($P>0.05$).

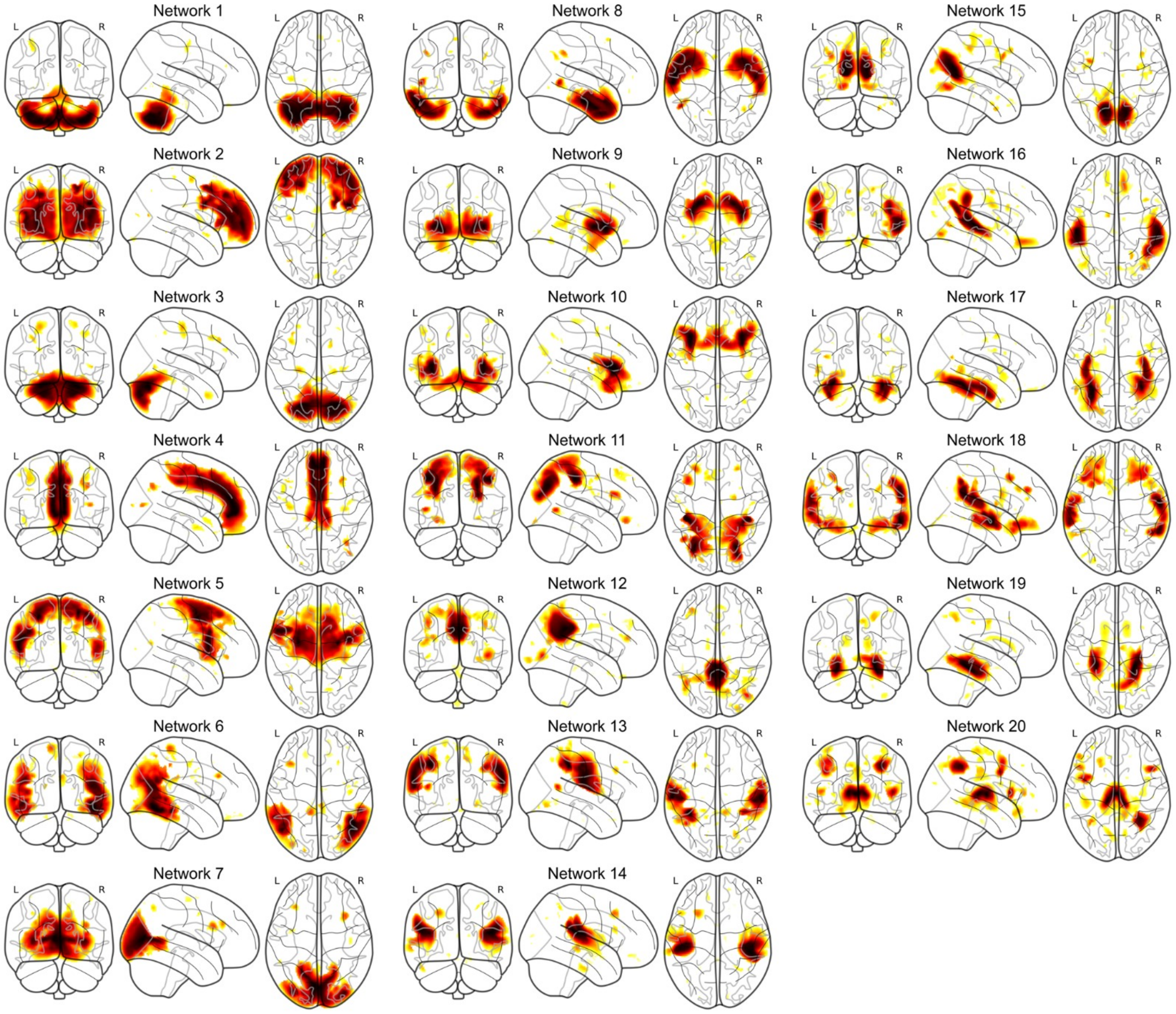


**eFigure 3**. **Structural covariance networks delineated by Non-Negative Matrix Factorization analysis.**
Structural covariance networks are shown for the 20-network solution. The yellow-orange color represents the spatial distribution of each network. For each network, we show the sagittal view (left hemisphere) that best captures the main areas of coverage. The anatomical coverage of each structural covariance network was a follows: (1) cerebellum I-IV, VIIIa, VIIb , crus II, vermis VIIIb to vermis IX; (2) frontal pole; (3) cerebellum V, VI, crus I, vermis VI to vermis VIIIa; (4) frontal medial cortex, paracingulate gyrus to anterior cingulate gyrus; (5) superior frontal gyrus, supplementary motor cortex to precentral gyrus; (6) lateral occipital cortex, angular gyrus to temporooccipital parts of middle and inferior temporal gyri; (7) occipital pole, supracalcarine cortex, intracalcarine cortex to lingual gyrus; (8) temporal pole to temporal fusiform cortex; (9) caudate, putamen ,pallidum, accumbens to amygdala; (10) frontal operculum cortex to insular cortex; (11) superior poscentral gyrus to superior lateral occipital cortex; (12) posterior cingulate gyrus to anterior precuneous cortex; (13) inferior poscentral gyrus to central opercular cortex; (14) anterior supramarginal gyrus to parietal operculum cortex; (15) posterior precuneous cortex to cuneal cortex; (16) posterior supramarginal gyrus to posterior superior temporal gyrus; (17) occipital fusiform gyrus to temporal occipital fusiform cortex; (18) frontal orbital cortex; (19) hippocampus to parahippocampal gyrus; and (20) thalamus. We observed no significant effect of our exercise program on any of these 20 structural networks (P>0.05).

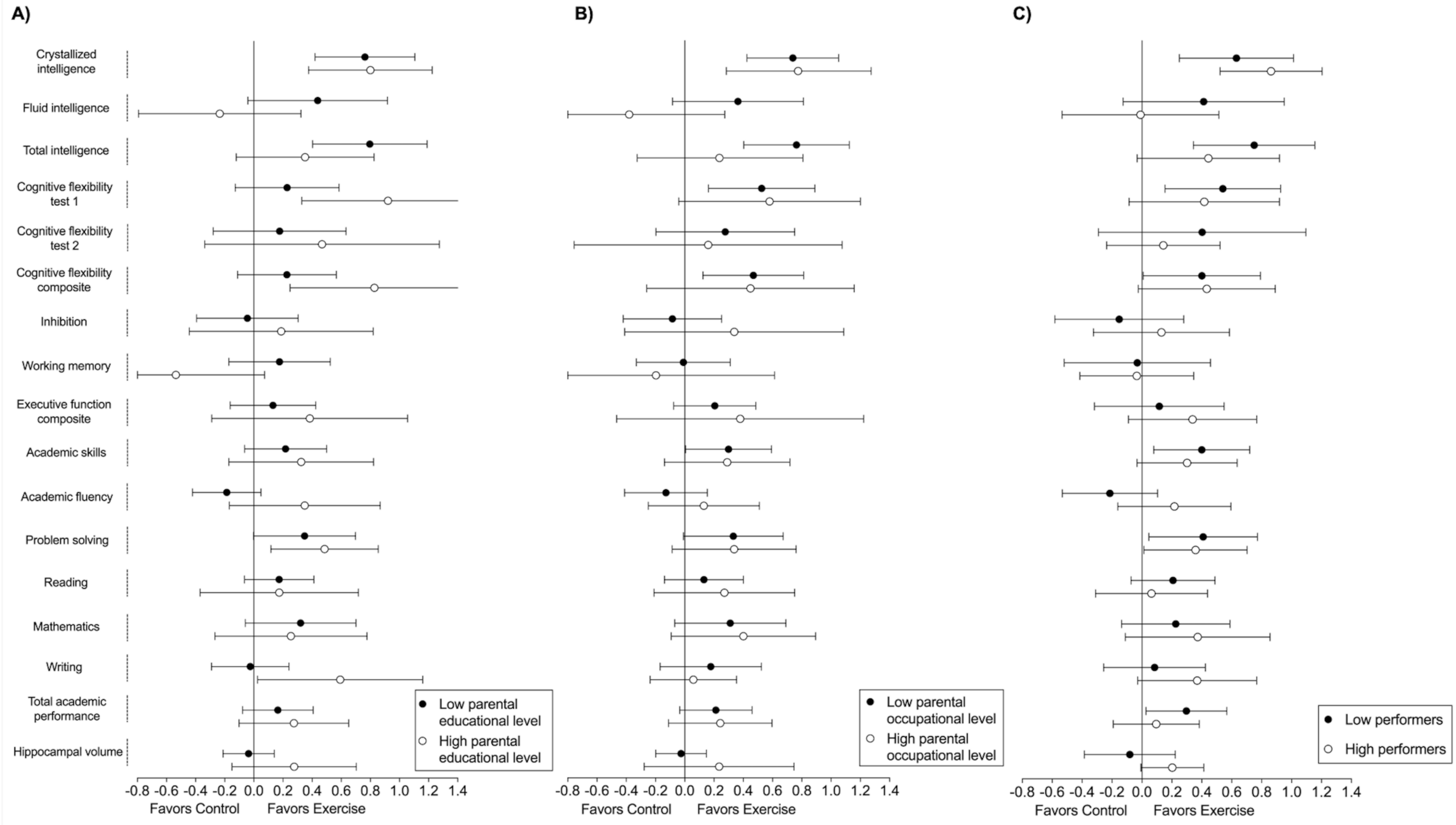
A)
B)
C)
Crystallized intelligence
Fluid intelligence
Total intelligence
Cognitive flexibility test 1
Cognitive flexibility test 2
Cognitive flexibility composite
Inhibition
Working memory
Executive function composite
Academic skills
Academic fluency
Problem solving
Reading
Mathematics
Writing
Total academic performance
Hippocampal volume
Low parental educational level
High parental educational level
Low parental occupational level
High parental occupational level
Low performers
High performers
-0.8 -0.6 -0.4 -0.2 0.0 0.2 0.4 0.6 0.8 1.0 1.2 1.4
Favors Control
Favors Exercise

**eFigure 4. Per-protocol effects of the *ActiveBrains* exercise program on the main brain health outcomes by parental educational levels (A), parental occupational levels (B), and baseline levels (C).**

Each analysis was adjusted by baseline outcomes. Dots represent the between-groups difference in z-score values of change, i.e., post-exercise outcomes with respect to the baseline mean and standard deviation. Bars represent 95% confidence intervals.

The educational level was defined as low when neither parent had university education or high when at least one of the parents had university education. Similarly, occupational level was defined as low when neither parent had a high occupational level and high when at least one of the parents had a high occupational level. Baseline level of performance was dichotomized by the median of the outcome of interest.

Intelligence outcomes (i.e., Crystallized intelligence, Fluid intelligence, and Total intelligence) were measured by the Kaufman Brief Intelligence Test.

Cognitive flexibility test 1 refers to the Design Fluency Test output. Cognitive flexibility test 2 refers to the Trail Making Test output. Cognitive flexibility composite z-score was calculated as the re-normalized mean of the z-scores for Cognitive flexibility test 1 and Cognitive flexibility test 2. Inhibition was measured by the Stroop Color-Word Test. Working memory was measured by the Delayed Non-Match-to sample computerized task. Executive function composite z-score was calculated as the re-normalized mean of the z-scores for Cognitive flexibility, Inhibition, and Working memory.

Academic performance outcomes were obtained from the Spanish version of the Woodcock Johnson III Test of Achievement. Academic skills are the sum of components based on basic skills such as reading decoding, mathematics calculation, and spelling. Academic fluency is the sum of tests based on reading, calculation and writing fluency. Total academic performance is the overall measure of the academic performance based on reading, mathematics, and writing.

Hippocampal volume was obtained using the FMRIB's Integrated Registration and Segmentation Tool (FIRST).

Two of the cognitive tests, i.e., the cognitive flexibility test 2 (Trail Making Test) and the cognitive inhibition test (Stroop Color-Word Test), are originally expressed in an inverse way, which means that lower scores indicate better performance. In order to make the visual interpretation of the main findings in these Figure easier, we inverted these two scores so that they can be interpreted in the same fashion as the rest of outcomes (i.e., higher score better performance). On the other hand, we express these cognitive tests in their original units and not inverted in **Supplementary Table 2** and **Supplementary Table 19**.

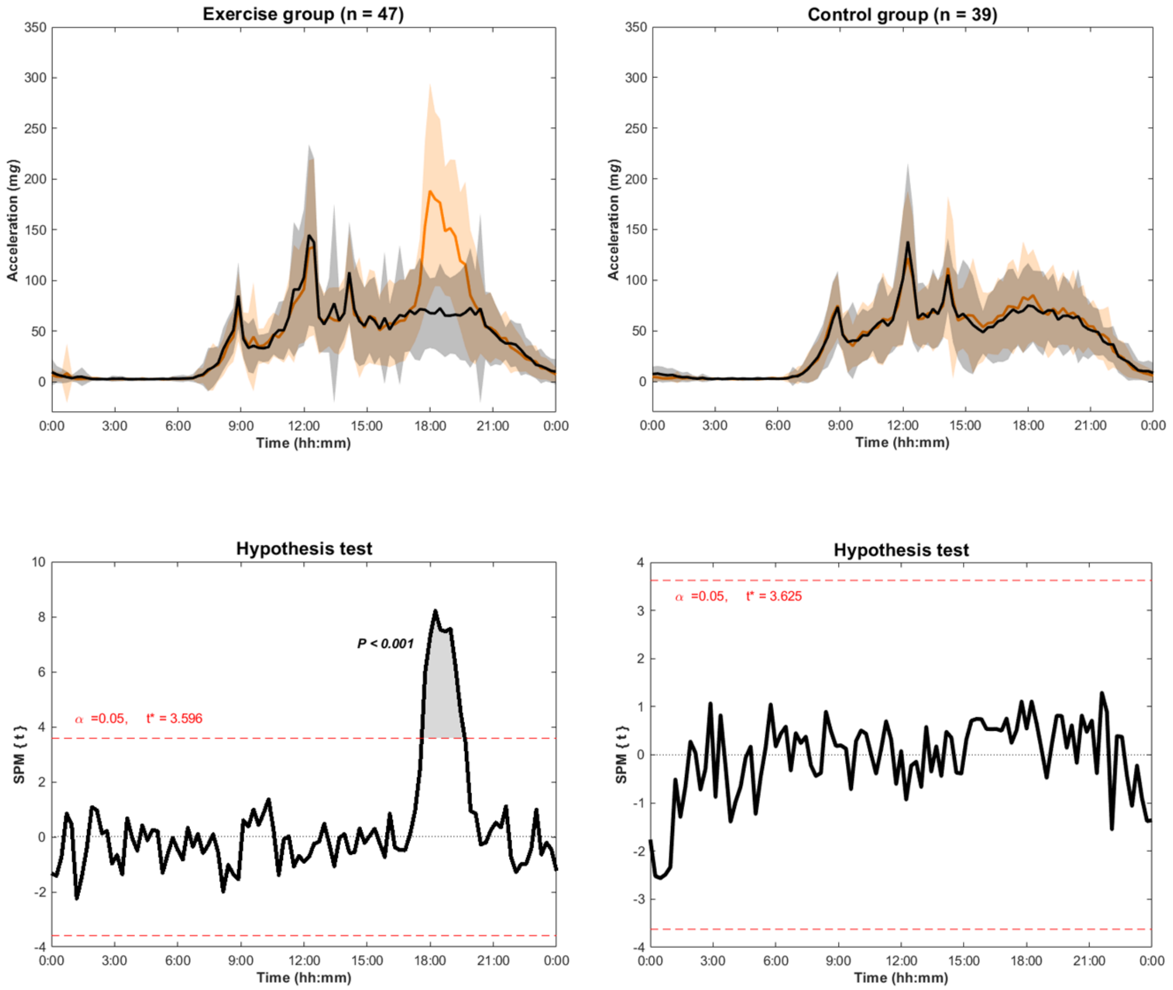


**eFigure 5. Comparison of the 24 h physical activity patterns derived from aggregated raw accelerations (i.e., Euclidean Norm Minus One accelerations) measured with an accelerometer attached at the non-dominant wrist at baseline (i.e., black line) and in the middle of the exercise program (i.e., orange line) in exercise and control groups.**

The hypothesis test shows the threshold ($t^* = 3.630$) at which there are significant physical activity patterns' differences between baseline and during-exercise periods.

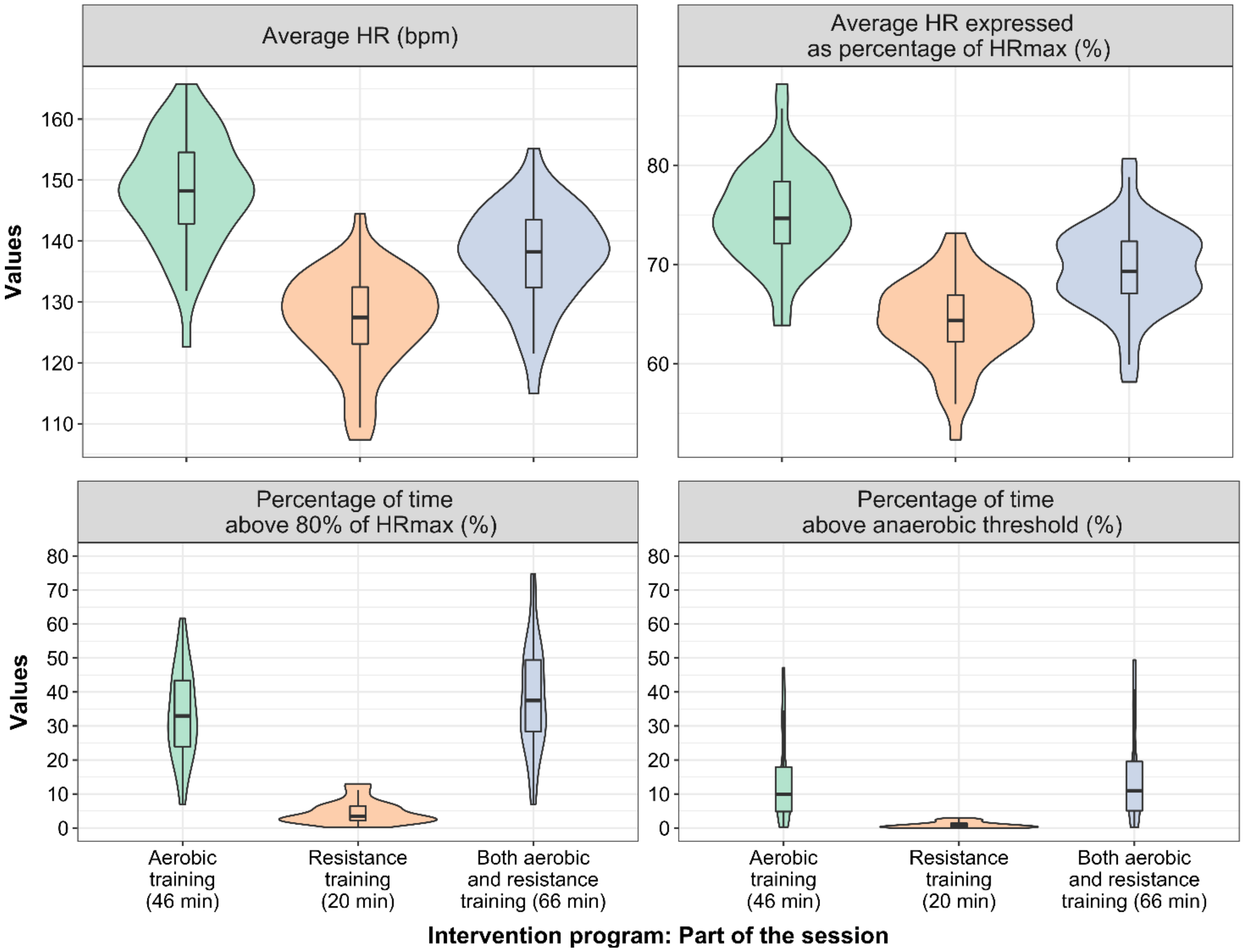


**eFigure 6. Violin plots characterizing the intensity of the exercise program as measured by heart rate (HR) monitors.**

The exercise session was divided in two parts, 46 min of aerobic exercise and 20 min of resistance exercise based on group games, playful exercises with a coordinative component. These times, together with the time spent in arrival, setting HR monitors, warming-up and cooling-down accumulate the planned 90 min per session. The maximum HR and the HR at the anaerobic threshold were determined individually by incremental maximal exercise testing before the intervention started and the HR monitors were individually programmed with those values so that we could later obtain the intensity indicators shown in the figure, i.e., average HR expressed as percentage of the maximum HR, time above 80% of the maximum HR and time above the anaerobic threshold.

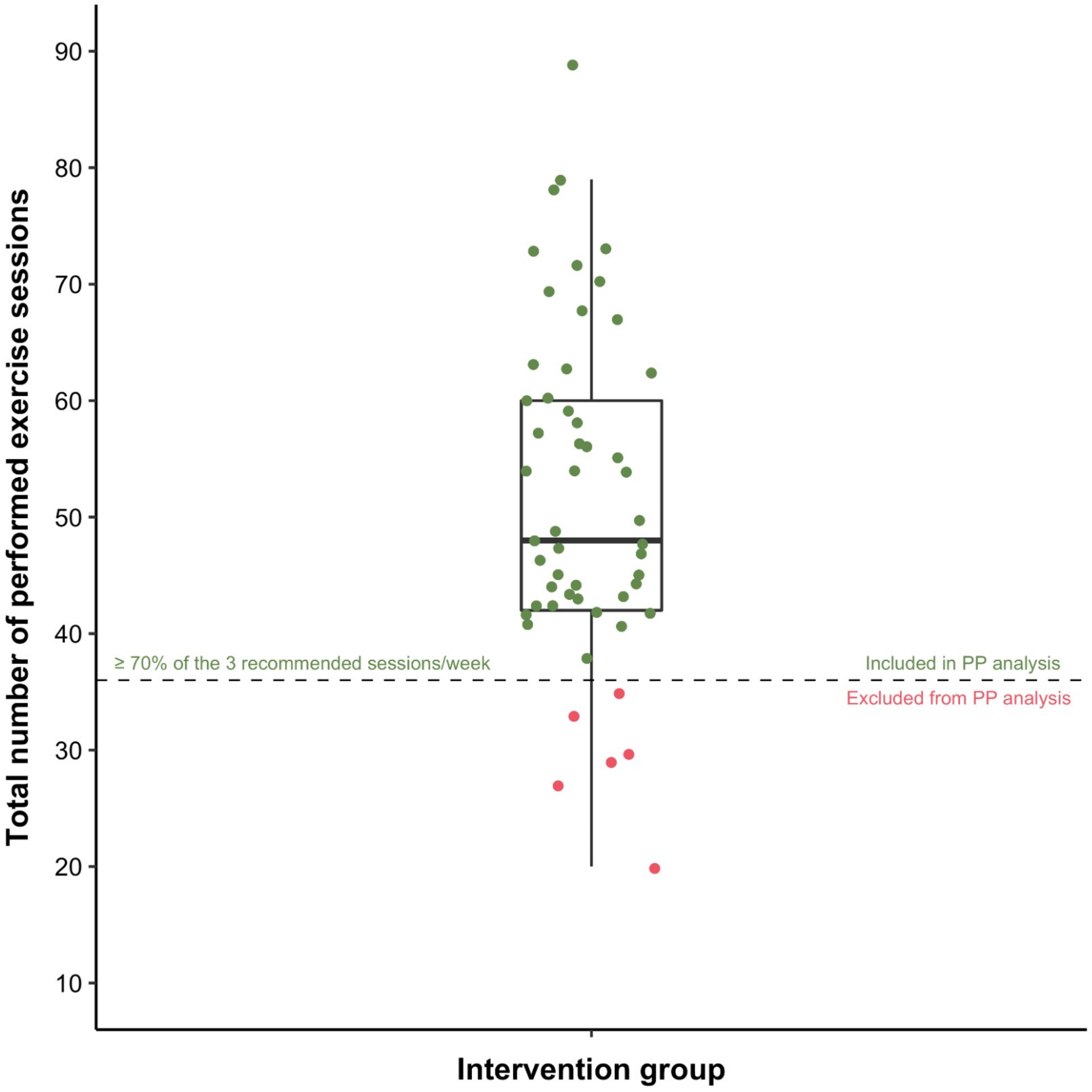


**eFigure 7. Box plot showing the distribution of the attendance to the exercise program.**
PP = Per-Protocol analyses.

**Supplementary eTable 1. Descriptive baseline characteristics of the *ActiveBrains* participants meeting intention-to-treat criteria.**

| | All | | Exercise group | | Control group | |
|---|---|---|---|---|---|---|
| | N | Mean ± SD | N | Mean ± SD | N | Mean ± SD |
| Age (years) | 109 | 10.04 ± 1.13 | 57 | 9.99 ± 1.13 | 52 | 10.10 ± 1.13 |
| Sex | | | | | | |
| Girls (n %) | 45 | 59% | 20 | 35% | 25 | 48% |
| Boys (n %) | 64 | 41% | 37 | 65% | 27 | 52% |
| Weight (kg) | 109 | 56.21 ± 11.23 | 57 | 56.72 ± 12.75 | 52 | 55.66 ± 9.38 |
| Height (cm) | 109 | 144.22 ± 8.41 | 57 | 143.36 ± 8.87 | 52 | 145.16 ± 7.86 |
| Body mass index ($kg/m^2$) | 109 | 26.81 ± 3.62 | 57 | 27.28 ± 4.10 | 52 | 26.29 ± 2.97 |
| Peak height velocity (years) | 109 | –2.26 ± 0.99 | 57 | –2.40 ± 0.91 | 52 | –2.10 ± 1.05 |
| Wave of participation (%) | | | | | | |
| First (n %) | 19 | 18% | 10 | 17% | 9 | 17% |
| Second (n %) | 45 | 41% | 22 | 39% | 23 | 44% |
| Third (n %) | 45 | 41% | 25 | 44% | 20 | 39% |
| Cardiorespiratory fitness | | | | | | |
| Final time in treadmill test (min) | 109 | 8.53 ± 2.76 | 57 | 8.09 ± 2.69 | 52 | 9.02 ± 2.77 |
| Relative $VO_2$peak (mL/kg/min) | 109 | 37.35 ± 4.74 | 57 | 36.72 ± 4.68 | 52 | 38.04 ± 4.76 |
| Intelligence | | | | | | |
| Crystallized intelligence (typical punctuation) | 109 | 103.07 ± 12.84 | 57 | 103.56 ± 13.83 | 52 | 102.54 ± 11.77 |
| Fluid intelligence (typical punctuation) | 109 | 97.68 ± 13.13 | 57 | 96.39 ± 14.02 | 52 | 99.10 ± 12.06 |
| Total intelligence (typical punctuation) | 109 | 98.06 ± 12.43 | 57 | 97.61 ± 13.20 | 52 | 98.54 ± 11.63 |
| Executive function | | | | | | |
| Cognitive flexibility 1 (total correct designs) | 109 | 20.01 ± 6.48 | 57 | 19.77 ± 6.15 | 52 | 20.27 ± 6.88 |
| Cognitive flexibility 2 (sec) | 109 | 91.76 ± 43.03 | 57 | 90.07 ± 41.62 | 52 | 93.60 ± 44.85 |
| Cognitive flexibility composite z-score | 109 | 0.00 ± 1.00 | 57 | 0.00 ± 0.95 | 52 | 0.00 ± 1.06 |
| Inhibition (sec) | 109 | 40.62 ± 17.01 | 57 | 40.54 ± 14.72 | 52 | 40.69 ± 19.36 |
| Working memory (% response accuracy) | 109 | 65.33 ± 16.6 | 57 | 67.18 ± 14.91 | 52 | 63.31 ± 18.21 |
| Executive function composite z-score | 109 | 0.00 ± 1.00 | 57 | 0.04 ± 0.87 | 52 | –0.05 ± 1.13 |
| Academic performance (standard score) | | | | | | |
| Academic skills | 109 | 118.86 ± 15.63 | 57 | 119.86 ± 16.03 | 52 | 117.77 ± 15.26 |
| Academic fluency | 109 | 104.04 ± 12.00 | 57 | 104.77 ± 11.02 | 52 | 103.23 ± 13.06 |
| Problem solving | 109 | 99.69 ± 9.24 | 57 | 101.83 ± 9.14 | 52 | 97.36 ± 8.87 |
| Reading | 109 | 107.71 ± 13.03 | 57 | 109.60 ± 13.08 | 52 | 105.61 ± 12.78 |
| Mathematics | 109 | 101.99 ± 10.91 | 57 | 104.26 ± 11.03 | 52 | 99.49 ± 10.32 |
| Writing | 109 | 103.79 ± 9.04 | 57 | 103.58 ± 8.41 | 52 | 104.02 ± 9.77 |
| Total academic performance | 109 | 109.53 ± 11.83 | 57 | 111.07 ± 11.59 | 52 | 107.84 ± 11.96 |
| Hippocampal volume ($mm^3$) | 109 | 7037.18 ± 671.51 | 57 | 7074.67 ± 672.92 | 52 | 6996.08 ± 674.09 |

Values are expressed as means ± standard deviations (SD), unless otherwise indicated. Baseline differences between groups were determined by one-way analysis of variance (ANOVA) and chi-squared tests for continuous and categorical variables, respectively. Statistically significant values at $P < 0.05$ are shown in bold.

Intelligence outcomes (i.e., Crystallized, Fluid, and Total Intelligence) were measured by the Kaufman Brief Intelligence Test.

Cognitive flexibility 1 was measured by the Design Fluency Test and expressed as number of total correct designs of the three conditions. ‡Cognitive flexibility 2 was measured by the Trail Making Test and expressed as the total completion time (sec) of Part A subtracted from the total completion time (sec) of Part B. A smaller B – A difference score (sec) indicated better cognitive flexibility.

Cognitive flexibility composite z-score was calculated as the re-normalized mean of the z-scores for Cognitive flexibility 1 and Cognitive flexibility 2.
Inhibition was measured by the Stroop Color-Word Test. The inhibition score was obtained by subtracting condition 3 completion time – condition 1 completion time (sec). The lower the difference between tests' times, the better the performance was considered.
Working memory was measured by the Delayed Non-Match-to sample task.
Executive function composite z-score was calculated as the re-normalized mean of the z-scores for Cognitive flexibility, Inhibition, and Working memory.
Academic performance was measured by the Spanish version of the Woodcock Johnson III Test of Achievement.
Academic skills are the sum of components based on basic skills such as reading decoding, mathematics calculation, and spelling.
Academic fluency is the sum of the components based on reading, calculation, and writing fluency.
Problem solving is the sum of the components based on solving academic problems in reading, mathematics, and writing.
Total academic performance is the overall measure of the academic performance based on reading, mathematics, and writing.

**Supplementary eTable 2. Per-protocol effects of the *ActiveBrains* exercise program on raw and z-score post-exercise (i.e. z-score of change from baseline) intelligence and executive function outcomes**

| | | | Mean (95% CI) | | | | |
|---|---|---|---|---|---|---|---|
| | $N_{all}$ | N | Exercise group | N | Control group | Difference between groups | *P* |
| Crystallized intelligence | 90 | 47 | | 43 | | | |
| Raw score (typical punctuation) | | | 111.04 (108.79 to 113.28) | | 102.00 (99.65 to 104.34) | 9.04 (5.79 to 12.28) | **0.0000003** |
| z-score | | | 0.62 (0.44 to 0.80) | | –0.10 (–0.28 to 0.09) | 0.72 (0.46 to 0.97) | |
| Fluid intelligence | 90 | 47 | | 43 | | | |
| Raw score (typical punctuation) | | | 103.25 (99.95 to 106.55) | | 100.49 (97.04 to 103.94) | 2.76 (–2.03 to 7.55) | 0.256 |
| z-score | | | 0.44 (0.18 to 0.68) | | 0.23 (–0.03 to 0.48) | 0.20 (–0.15 to 0.57) | |
| Total intelligence | 90 | 47 | | 43 | | | |
| Raw score (typical punctuation) | | | 106.36 (103.88 to 108.83) | | 98.90 (96.32 to 101.49) | 7.45 (3.86 to 11.03) | **0.00008** |
| z-score | | | 0.69 (0.48 to 0.89) | | 0.07 (–0.14 to 0.28) | 0.62 (0.31 to 0.91) | |
| Cognitive flexibility 1 | 90 | 47 | | 43 | | | |
| Raw score (total correct designs) | | | 24.31 (22.96 to 25.65) | | 21.26 (19.85 to 22.67) | 3.04 (1.08 to 4.99) | **0.003** |
| z-score | | | 0.65 (0.44 to 0.86) | | 0.18 (–0.04 to 0.39) | 0.48 (0.17 to 0.78) | |
| Cognitive flexibility 2 | 84 | 47 | | 37 | | | |
| Raw score (sec) | | | 74.43 (63.23 to 85.64) | | 86.04 (73.41 to 98.66) | –11.60 (–28.48 to 5.27) | 0.175 |
| z-score | | | –0.35 (–0.60 to –0.09) | | –0.08 (–0.40 to 0.21) | –0.26 (–0.65 to 0.12) | |
| Cognitive flexibility composite z-score | 84 | 47 | 0.25 (0.05 to 0.44) | 37 | –0.17 (–0.39 to 0.04) | 0.42 (0.13 to 0.71) | **0.005** |
| Inhibition | 90 | 47 | | 43 | | | |
| Raw score (sec) | | | 31.96 (28.28 to 35.64) | | 32.32 (28.47 to 36.17) | –0.59 (–5.84 to 4.65) | 0.821 |
| z-score | | | –0.51 (–0.72 to –0.30) | | –0.48 (–0.70 to –0.25) | –0.04 (–0.34 to 0.27) | |
| Working memory | 86 | 45 | | 41 | | | |
| Raw score (% accuracy) | | | 65.36 (61.87 to 68.86) | | 66.01 (62.35 to 69.67) | –0.64 (–5.73 to 4.44) | 0.801 |
| z-score | | | 0.01 (–0.20 to 0.22) | | 0.05 (–0.17 to 0.27) | –0.04 (–0.35 to 0.27) | |
| Executive function composite z-score | 82 | 45 | 0.13 (–0.06 to 0.32) | 37 | –0.08 (–0.30 to 0.13) | 0.21 (–0.06 to 0.50) | 0.136 |

z-score values indicate how many standard deviations have the post-exercise program values changed with respect to the baseline mean and standard deviation. E.g., a 0.50 z-score means that the mean value at post-exercise program is 0.50 standard deviations higher than the mean value at baseline, indicating a positive change, with negative values indicating the opposite.
All data presented were adjusted for baseline values of the outcome studied.
Crystallized, Fluid, and Total Intelligence were measured by the Kaufman Brief Intelligence Test.
Cognitive flexibility 1 was measured by the Design Fluency Test and expressed as number of total correct designs of the three conditions.
Cognitive flexibility 2 was measured by the Trail Making Test and expressed as the total completion time (sec) of Part A subtracted from the total completion time (sec) of Part B. A smaller B – A smaller difference in this score (sec) indicated better cognitive flexibility. Note: In the Figures, the z-score is presented inverted for easier visual interpretation in the same direction than the rest of outcomes, but here in the table are presented the real non-inverted values.
Cognitive flexibility composite z-score was calculated as the re-normalized mean of the z-scores for Cognitive flexibility 1 and Cognitive flexibility 2.
Inhibition was measured by the Stroop Color-Word Test. The inhibition score was obtained by subtracting condition 3 completion time – condition 1 completion time (sec). The smaller the difference between tests' times, the better the performance was considered. Note: In the Figures, the z-score is presented inverted for easier visual interpretation in the same direction than the rest of outcomes, but here in the table are presented the real non-inverted values.
Working memory was measured by the Delayed Non-Match-to sample task. The response accuracy (%) in the high load was used as an indicator of working memory. Higher response accuracy refers to better performance.
Executive function composite z-score was calculated as the re-normalized mean of the z-scores for Cognitive flexibility, Inhibition, and Working memory.

**Supplementary eTable 3. Per-protocol effects of the *ActiveBrains* exercise program on raw (standard score) and z-score post-exercise (z-score of change from baseline) academic performance outcomes (Woodcock-Muñoz standardized test).**

| | | | Mean (95% CI) | | | | |
|---|---|---|---|---|---|---|---|
| | $N_{all}$ | N | Exercise group | N | Control group | Difference between groups | *P* |
| Academic skills | 89 | 47 | | 42 | | | |
| Raw score | | | 124.25 (121.78 to 126.72) | | 120.14 (117.52 to 122.75) | 4.11 (0.50 to 7.72) | **0.026** |
| z-score | | | 0.27 (0.11 to 0.43) | | 0.01 (–0.16 to 0.17) | 0.27 (0.03 to 0.49) | |
| Academic fluency | 89 | 47 | | 42 | | | |
| Raw score | | | 106.10 (104.15 to 108.05) | | 106.23 (104.17 to 108.30) | –0.27 (–3.06 to 2.51) | 0.845 |
| z-score | | | 0.16 (0.00 to 0.32) | | 0.18 (0.01 to 0.35) | –0.02 (–0.26 to 0.21) | |
| Problem solving | 89 | 47 | | 42 | | | |
| Raw score | | | 103.11 (101.51 to 104.72) | | 99.84 (98.14 to 101.54) | 3.27 (0.91 to 5.64) | **0.007** |
| z-score | | | 0.41 (0.24 to 0.59) | | 0.05 (–0.13 to 0.24) | 0.36 (0.10 to 0.62) | |
| Reading | 89 | 47 | | 42 | | | |
| Raw score | | | 111.57 (109.64 to 113.50) | | 109.64 (107.59 to 111.69) | 1.93 (–0.90 to 4.76) | 0.180 |
| z-score | | | 0.23 (0.07 to 0.38) | | 0.07 (–0.09 to 0.23) | 0.15 (–0.07 to 0.37) | |
| Mathematics | 89 | 47 | | 42 | | | |
| Raw score | | | 105.72 (103.59 to 107.84) | | 102.33 (100.00 to 104.58) | 3.38 (0.24 to 6.52) | **0.035** |
| z-score | | | 0.35 (0.15 to 0.55) | | 0.04 (–0.17 to 0.25) | 0.32 (0.02 to 0.60) | |
| Writing | 89 | 47 | | 42 | | | |
| Raw score | | | 118.89 (116.71 to 121.07) | | 116.42 (114.12 to 118.73) | 2.47 (–0.70 to 5.64) | 0.125 |
| z-score | | | 0.31 (0.13 to 0.48) | | 0.11 (–0.07 to 0.29) | 0.19 (–0.05 to 0.45) | |
| Total performance | 89 | 47 | | 42 | | | |
| Raw score | | | 113.60 (112.04 to 115.16) | | 111.12 (109.47 to 112.77) | 2.48 (0.19 to 4.77) | **0.034** |
| z-score | | | 0.31 (0.18 to 0.44) | | 0.10 (–0.04 to 0.24) | 0.21 (0.01 to 0.40) | |

z-score values indicate how many standard deviations have the post-exercise program values changed with respect to the baseline mean and standard deviation. E.g., a 0.50 z-score means that the mean value at post-exercise program is 0.50 standard deviations higher than the mean value at baseline, indicating a positive change, with negative values indicating the opposite.
All data presented were adjusted for baseline values of the outcome studied.
Academic skills are the sum of components based on basic skills such as reading decoding, mathematics calculation, and spelling.
Academic fluency is the sum of the components based on reading, calculation, and writing fluency.
Problem solving is the sum of the components based on solving academic problems in reading, mathematics, and writing.
Total performance is the overall measure of the academic performance based on reading, mathematics, and writing.

**Supplementary eTable 4. Per-protocol effects of the *ActiveBrains* exercise program on raw (mm³) and z-scores of post-exercise hippocampal volume**

| | | | Mean (95% CI) | | | | |
|---|---|---|---|---|---|---|---|
| | $N_{all}$ | N | Exercise group | N | Control group | Difference between groups | *P* |
| Hippocampus | 82 | 45 | | 37 | | | |
| Raw score | | | 7178.88 (7094.44 to 7263.32) | | 7136.23 (7043.10 to 7229.36) | 42.64 (–83.11 to 168.40) | 0.502 |
| z-score | | | 0.19 (0.07 to 0.32) | | 0.13 (–0.00 to 0.27) | 0.06 (–0.12 to 0.24) | |
| Right hippocampus | 82 | 45 | | 37 | | | |
| Raw score | | | 3673.63 (3622.00 to 3725.26) | | 3658.15 (3601.20 to 3715.10) | 15.47 (–61.45 to 92.40) | 0.690 |
| z-score | | | 0.22 (0.09 to 0.35) | | 0.18 (0.03 to 0.33) | 0.04 (–0.16 to 0.24) | |
| Right anterior hippocampus | 82 | 45 | | 37 | | | |
| Raw score | | | 2115.25 (2081.69 to 2148.81) | | 2098.60 (2061.57 to 2135.62) | 16.65 (–33.40 to 66.70) | 0.510 |
| z-score | | | 0.22 (0.09 to 0.36) | | 0.16 (0.00 to 0.31) | 0.07 (–0.14 to 0.28) | |
| Right posterior hippocampus | 82 | 45 | | 37 | | | |
| Raw score | | | 1556.19 (1531.98 to 1580.40) | | 1554.16 (1527.46 to 1580.86) | 2.03 (–34.00 to 38.00) | 0.911 |
| z-score | | | 0.19 (0.03 to 0.35) | | 0.18 (0.00 to 0.35) | 0.01 (–0.22 to 0.25) | |
| Left hippocampus | 82 | 45 | | 37 | | | |
| Raw score | | | 3504.09 (3444.55 to 3563.64) | | 3479.47 (3413.80 to 3545.14) | 24.62 (–64.03 to 113.27) | 0.582 |
| z-score | | | 0.13 (–0.03 to 0.28) | | 0.06 (–0.11 to 0.23) | 0.06 (–0.17 to 0.30) | |
| Left anterior hippocampus | 82 | 45 | | 37 | | | |
| Raw score | | | 2015.62 (1981.01 to 2050.23) | | 1998.28 (1960.12 to 2036.45) | 17.34 (–34.19 to 68.86) | 0.505 |
| z-score | | | 0.12 (–0.03 to 0.27) | | 0.04 (–0.13 to 0.21) | 0.08 (–0.15 to 0.31) | |
| Left posterior hippocampus | 82 | 45 | | 37 | | | |
| Raw score | | | 1488.66 (1457.51 to 1519.80) | | 1476.53 (1442.18 to 1510.88) | 12.13 (–34.23 to 58.49) | 0.604 |
| z-score | | | 0.13 (–0.05 to 0.32) | | 0.06 (–0.15 to 0.26) | 0.07 (–0.20 to 0.35) | |

z-score values indicate how many standard deviations the post-exercise program values changed with respect to the baseline mean and standard deviation. e.g., a 0.50 z-score means that the mean value at post-exercise program is 0.50 standard deviations higher than the mean value at baseline, indicating a positive change, with negative values indicating the opposite.

All data presented were adjusted for baseline values of the outcome studied.

**Supplementary eTable 5. Per-protocol effects of the ActiveBrains exercise program on raw ($mm^3$) and z-scores of post-exercise prefrontal cortex gray matter volume outcomes.**

| | | | Mean (95% CI) | | | | |
|---|---|---|---|---|---|---|---|
| | Nall | N | Exercise group | N | Control group | Difference between groups | *P* |
| L cingulate volume, anterior division | | | | | | | |
| Raw score | 82 | 45 | 5383.08 (5165.28 to 5600.88) | 37 | 5418.49 (5178.27 to 5658.72) | -35.41 (-359.89 to 289.06) | 0.829 |
| z-score | 82 | 45 | -0.08 (-0.34 to 0.19) | 37 | -0.03 (-0.33 to 0.26) | -0.04 (-0.44 to 0.36) | |
| L inferior frontal volume, pars opercularis | | | | | | | |
| Raw score | 82 | 45 | 4207.93 (4055.96 to 4359.89) | 37 | 4293.31 (4125.49 to 4461.12) | -85.38 (-313.32 to 142.56) | 0.458 |
| z-score | 82 | 45 | -0.22 (-0.48 to 0.03) | 37 | -0.08 (-0.36 to 0.2) | -0.14 (-0.52 to 0.24) | |
| L inferior frontal volume, pars triangularis | | | | | | | |
| Raw score | 82 | 45 | 3493.18 (3368.65 to 3617.72) | 37 | 3568.7 (3431.34 to 3706.05) | -75.52 (-261.02 to 109.98) | 0.420 |
| z-score | 82 | 45 | -0.14 (-0.35 to 0.08) | 37 | -0.01 (-0.24 to 0.23) | -0.13 (-0.45 to 0.19) | |
| L middle frontal volume | | | | | | | |
| Raw score | 82 | 45 | 13156.85 (12781 to 13532.7) | 37 | 13019.29 (12604.14 to 13434.44) | 137.56 (-426.93 to 702.05) | 0.629 |
| z-score | 82 | 45 | -0.11 (-0.27 to 0.06) | 37 | -0.17 (-0.35 to 0.01) | 0.06 (-0.19 to 0.31) | |
| L superior frontal volume | | | | | | | |
| Raw score | 82 | 45 | 20617.07 (20113.41 to 21120.72) | 37 | 20712.05 (20156.28 to 21267.82) | -94.98 (-847.28 to 657.31) | 0.802 |
| z-score | 82 | 45 | -0.19 (-0.42 to 0.03) | 37 | -0.15 (-0.4 to 0.1) | -0.04 (-0.37 to 0.29) | |
| L orbital volume | | | | | | | |
| Raw score | 82 | 45 | 7276.4 (7041.07 to 7511.73) | 37 | 7306.54 (7046.78 to 7566.3) | -30.14 (-382.26 to 321.98) | 0.865 |
| z-score | 82 | 45 | -0.08 (-0.3 to 0.14) | 37 | -0.05 (-0.3 to 0.19) | -0.03 (-0.36 to 0.3) | |
| R cingulate volume, anterior division | | | | | | | |
| Raw score | 82 | 45 | 6293.6 (6113.67 to 6473.53) | 37 | 6307.11 (6108.23 to 6505.98) | -13.51 (-284.7 to 257.69) | 0.921 |
| z-score | 82 | 45 | 0.05 (-0.16 to 0.26) | 37 | 0.07 (-0.16 to 0.3) | -0.02 (-0.33 to 0.3) | |
| R inferior frontal volume, pars opercularis | | | | | | | |
| Raw score | 82 | 45 | 3871.48 (3768.03 to 3974.93) | 37 | 3766.82 (3652.72 to 3880.91) | 104.66 (-49.41 to 258.74) | 0.180 |
| z-score | 82 | 45 | 0.09 (-0.07 to 0.25) | 37 | -0.07 (-0.25 to 0.1) | 0.16 (-0.08 to 0.4) | |
| R inferior frontal volume, pars triangularis | | | | | | | |
| Raw score | 82 | 45 | 3291.33 (3187.96 to 3394.7) | 37 | 3294.3 (3180.28 to 3408.32) | -2.97 (-157.06 to 151.12) | 0.969 |
| z-score | 82 | 45 | 0.04 (-0.1 to 0.19) | 37 | 0.05 (-0.12 to 0.21) | 0 (-0.22 to 0.22) | |
| R middle frontal volume | | | | | | | |

| | | | | | | | |
|---|---|---|---|---|---|---|---|
| Raw score | 82 | 45 | 11854.95 (11479.12 to 12230.77) | 37 | 11517.31 (11102.17 to 11932.45) | 337.64 (-226.96 to 902.24) | 0.237 |
| z-score | 82 | 45 | 0.01 (-0.18 to 0.2) | 37 | -0.16 (-0.37 to 0.05) | 0.17 (-0.11 to 0.45) | |
| R superior frontal volume | | | | | | | |
| Raw score | 82 | 45 | 19306.95 (18840.06 to 19773.83) | 37 | 19521.5 (19006.23 to 20036.76) | -214.55 (-912.45 to 483.35) | 0.542 |
| z-score | 82 | 45 | -0.24 (-0.45 to -0.03) | 37 | -0.14 (-0.38 to 0.09) | -0.1 (-0.41 to 0.22) | |
| R orbital volume | | | | | | | |
| Raw score | 82 | 45 | 7805.98 (7579.03 to 8032.94) | 37 | 7666.43 (7415.76 to 7917.09) | 139.56 (-201.14 to 480.25) | 0.417 |
| z-score | 82 | 45 | 0.02 (-0.2 to 0.25) | 37 | -0.12 (-0.36 to 0.13) | 0.14 (-0.2 to 0.48) | |

z-score values indicate how many standard deviations have the post-exercise program values changed with respect to the baseline mean and standard deviation. E.g., a 0.50 z-score means that the mean value at post-exercise program is 0.50 standard deviations higher than the mean value at baseline, indicating a positive change, with negative values indicating the opposite. All data presented were adjusted for baseline values.

**Supplementary eTable 6. Per-protocol effects of the ActiveBrains exercise program on raw (mm) and z-scores of post-exercise prefrontal cortex cortical thickness outcomes.**
**Mean (95% CI)**

| | Nall | N | Exercise group | N | Control group | Difference between groups | *P* |
|---|---|---|---|---|---|---|---|
| L cingulate thickness, anterior division | | | | | | | |
| Raw score | 82 | 45 | 3.02 (2.97 to 3.06) | 37 | 2.98 (2.93 to 3.03) | 0.04 (-0.04 to 0.11) | 0.332 |
| z-score | 82 | 45 | 0.08 (-0.16 to 0.31) | 37 | -0.1 (-0.35 to 0.16) | 0.17 (-0.18 to 0.52) | |
| L inferior frontal thickness, pars opercularis | | | | | | | |
| Raw score | 82 | 45 | 3.08 (3.05 to 3.12) | 37 | 3.1 (3.06 to 3.13) | -0.02 (-0.06 to 0.04) | 0.562 |
| z-score | 82 | 45 | -0.26 (-0.49 to -0.03) | 37 | -0.16 (-0.41 to 0.1) | -0.1 (-0.44 to 0.24) | |
| L inferior frontal thickness, pars triangularis | | | | | | | |
| Raw score | 82 | 45 | 2.99 (2.93 to 3.04) | 37 | 3.03 (2.97 to 3.09) | -0.05 (-0.13 to 0.04) | 0.261 |
| z-score | 82 | 45 | -0.12 (-0.46 to 0.22) | 37 | 0.17 (-0.2 to 0.55) | -0.29 (-0.8 to 0.22) | |
| L middle frontal thickness | | | | | | | |
| Raw score | 82 | 45 | 2.93 (2.9 to 2.97) | 37 | 2.94 (2.9 to 2.98) | 0 (-0.06 to 0.05) | 0.929 |
| z-score | 82 | 45 | -0.22 (-0.46 to 0.02) | 37 | -0.2 (-0.47 to 0.06) | -0.02 (-0.38 to 0.35) | |
| L superior frontal thickness | | | | | | | |
| Raw score | 82 | 45 | 3.15 (3.12 to 3.18) | 37 | 3.14 (3.1 to 3.18) | 0.01 (-0.04 to 0.06) | 0.766 |
| z-score | 82 | 45 | -0.12 (-0.33 to 0.08) | 37 | -0.17 (-0.4 to 0.06) | 0.05 (-0.26 to 0.35) | |
| L orbital thickness | | | | | | | |
| Raw score | 82 | 45 | 3.05 (2.99 to 3.1) | 37 | 3.03 (2.98 to 3.09) | 0.01 (-0.06 to 0.09) | 0.763 |
| z-score | 82 | 45 | 0.02 (-0.2 to 0.24) | 37 | -0.03 (-0.27 to 0.21) | 0.05 (-0.28 to 0.38) | |
| R cingulate thickness, anterior division | | | | | | | |
| Raw score | 82 | 45 | 2.82 (2.78 to 2.86) | 37 | 2.8 (2.75 to 2.84) | 0.02 (-0.04 to 0.09) | 0.448 |
| z-score | 82 | 45 | 0.02 (-0.21 to 0.24) | 37 | -0.11 (-0.36 to 0.13) | 0.13 (-0.2 to 0.46) | |
| R inferior frontal thickness, pars opercularis | | | | | | | |
| Raw score | 82 | 45 | 3.05 (3.02 to 3.09) | 37 | 3.06 (3.02 to 3.1) | 0 (-0.06 to 0.05) | 0.922 |
| z-score | 82 | 45 | -0.14 (-0.36 to 0.08) | 37 | -0.13 (-0.37 to 0.12) | -0.02 (-0.34 to 0.31) | |

| | | | | | | | |
|---|---|---|---|---|---|---|---|
| R inferior frontal thickness, pars triangularis | | | | | | | |
| Raw score | 82 | 45 | 2.92 (2.88 to 2.97) | 37 | 2.92 (2.87 to 2.97) | 0 (-0.07 to 0.07) | 0.995 |
| z-score | 82 | 45 | -0.04 (-0.26 to 0.17) | 37 | -0.04 (-0.28 to 0.2) | 0 (-0.32 to 0.32) | |
| R middle frontal thickness | | | | | | | |
| Raw score | 82 | 45 | 2.85 (2.81 to 2.88) | 37 | 2.79 (2.75 to 2.83) | 0.06 (0 to 0.11) | 0.051 |
| z-score | 82 | 45 | 0.01 (-0.23 to 0.26) | 37 | -0.35 (-0.62 to -0.08) | 0.36 (0 to 0.73) | |
| R superior frontal thickness | | | | | | | |
| Raw score | 82 | 45 | 3.03 (3 to 3.07) | 37 | 3.02 (2.98 to 3.06) | 0.01 (-0.04 to 0.07) | 0.638 |
| z-score | 82 | 45 | -0.22 (-0.46 to 0.02) | 37 | -0.3 (-0.57 to -0.04) | 0.08 (-0.27 to 0.44) | |
| R orbital thickness | | | | | | | |
| Raw score | 82 | 45 | 3.06 (3.01 to 3.11) | 37 | 3.01 (2.96 to 3.06) | 0.05 (-0.02 to 0.12) | 0.189 |
| z-score | 82 | 45 | 0.09 (-0.13 to 0.3) | 37 | -0.12 (-0.36 to 0.11) | 0.21 (-0.11 to 0.54) | |

z-score values indicate how many standard deviations have the post-exercise program values changed with respect to the baseline mean and standard deviation. e.g., a 0.50 z-score means that the mean value at post-exercise program is 0.50 standard deviations higher than the mean value at baseline, indicating a positive change, with negative values indicating the opposite. All data presented were adjusted for baseline values.

**Supplementary eTable 7. Per-protocol effects of the ActiveBrains exercise program on raw ($mm^2$) and z-scores of post-exercise prefrontal cortex surface area outcomes.**

| | | | Mean (95% CI) | | | | |
|---|---|---|---|---|---|---|---|
| | Nall | N | Exercise group | N | Control group | Difference between groups | *P* |
| L cingulate area, anterior division | | | | | | | |
| Raw score | 82 | 45 | 1606.24 (1544.18 to 1668.3) | 37 | 1627.87 (1559.4 to 1696.34) | -21.63 (-114.23 to 70.97) | 0.643 |
| z-score | 82 | 45 | -0.07 (-0.29 to 0.15) | 37 | 0.01 (-0.24 to 0.25) | -0.08 (-0.4 to 0.25) | |
| L inferior frontal area, pars opercularis | | | | | | | |
| Raw score | 82 | 45 | 1001.05 (969.41 to 1032.69) | 37 | 1028.27 (993.37 to 1063.16) | -27.22 (-74.34 to 19.9) | 0.254 |
| z-score | 82 | 45 | -0.11 (-0.32 to 0.1) | 37 | 0.07 (-0.16 to 0.3) | -0.18 (-0.49 to 0.13) | |
| L inferior frontal area, pars triangularis | | | | | | | |
| Raw score | 82 | 45 | 851.64 (823.94 to 879.35) | 37 | 838.54 (807.98 to 869.11) | 13.1 (-28.21 to 54.42) | 0.530 |
| z-score | 82 | 45 | -0.03 (-0.21 to 0.15) | 37 | -0.11 (-0.31 to 0.09) | 0.08 (-0.18 to 0.35) | |
| L middle frontal area | | | | | | | |
| Raw score | 82 | 45 | 3258.98 (3175.34 to 3342.61) | 37 | 3207.97 (3115.46 to 3300.49) | 51.01 (-75.61 to 177.62) | 0.425 |
| z-score | 82 | 45 | 0.01 (-0.14 to 0.17) | 37 | -0.08 (-0.26 to 0.09) | 0.1 (-0.14 to 0.33) | |
| L superior frontal area | | | | | | | |
| Raw score | 82 | 45 | 5077 (4969.95 to 5184.06) | 37 | 5079.32 (4961.06 to 5197.58) | -2.31 (-163.21 to 158.58) | 0.977 |
| z-score | 82 | 45 | -0.08 (-0.26 to 0.09) | 37 | -0.08 (-0.27 to 0.11) | 0 (-0.26 to 0.26) | |
| L orbital area | | | | | | | |
| Raw score | 82 | 45 | 1738.01 (1696.56 to 1779.45) | 37 | 1733.89 (1688.12 to 1779.65) | 4.12 (-58 to 66.24) | 0.895 |
| z-score | 82 | 45 | -0.04 (-0.21 to 0.13) | 37 | -0.06 (-0.24 to 0.13) | 0.02 (-0.24 to 0.27) | |
| R cingulate area, anterior division | | | | | | | |
| Raw score | 82 | 45 | 1995.97 (1944.57 to 2047.37) | 37 | 1996.53 (1939.77 to 2053.28) | -0.56 (-77.64 to 76.52) | 0.989 |
| z-score | 82 | 45 | 0.09 (-0.09 to 0.28) | 37 | 0.1 (-0.11 to 0.3) | 0 (-0.28 to 0.28) | |
| R inferior frontal area, pars opercularis | | | | | | | |
| Raw score | 82 | 45 | 939.21 (915.91 to 962.5) | 37 | 905.8 (880.1 to 931.5) | 33.41 (-1.36 to 68.17) | 0.059 |
| z-score | 82 | 45 | 0.15 (0 to 0.29) | 37 | -0.06 (-0.21 to 0.1) | 0.2 (-0.01 to 0.42) | |
| R inferior frontal area, pars triangularis | | | | | | | |
| Raw score | 82 | 45 | 810.23 (785.34 to 835.11) | 37 | 800.97 (773.51 to 828.42) | 9.26 (-27.86 to 46.37) | 0.621 |
| z-score | 82 | 45 | 0.09 (-0.05 to 0.24) | 37 | 0.04 (-0.12 to 0.2) | 0.05 (-0.16 to 0.27) | |
| R middle frontal area | | | | | | | |

| | | | | | | | |
|---|---|---|---|---|---|---|---|
| Raw score | 82 | 45 | 2948.92 (2866.91 to 3030.93) | 37 | 2927.99 (2837.31 to 3018.67) | 20.93 (-102.93 to 144.78) | 0.738 |
| z-score | 82 | 45 | 0.02 (-0.15 to 0.19) | 37 | -0.02 (-0.21 to 0.17) | 0.04 (-0.22 to 0.3) | |
| R superior frontal area | | | | | | | |
| Raw score | 82 | 45 | 4772.08 (4674.39 to 4869.77) | 37 | 4850.5 (4742.64 to 4958.35) | -78.42 (-224.74 to 67.9) | 0.289 |
| z-score | 82 | 45 | -0.14 (-0.3 to 0.03) | 37 | 0 (-0.19 to 0.18) | -0.13 (-0.38 to 0.11) | |
| R orbital area | | | | | | | |
| Raw score | 82 | 45 | 1812.65 (1771.26 to 1854.04) | 37 | 1801.16 (1755.44 to 1846.87) | 11.49 (-50.66 to 73.65) | 0.714 |
| z-score | 82 | 45 | 0.02 (-0.17 to 0.21) | 37 | -0.03 (-0.24 to 0.18) | 0.05 (-0.23 to 0.34) | |

z-score values indicate how many standard deviations have the post-exercise program values changed with respect to the baseline mean and standard deviation. e.g., a 0.50 z-score means that the mean value at post-exercise program is 0.50 standard deviations higher than the mean value at baseline, indicating a positive change, with negative values indicating the opposite. All data presented were adjusted for baseline values.

**Supplementary eTable 8**. **Per-protocol effects of the ActiveBrains exercise program on raw (*β* values) and z-scores of post-exercise left hippocampal functional connectivity with prefrontal cortex subregions.**

| | Mean (95% CI) | | | | | | |
|---|---|---|---|---|---|---|---|
| | Nall | N | Exercise group | N | Control group | Difference between groups | *P* |
| L cingulate gyrus, anterior division | | | | | | | |
| Raw score | 80 | 44 | 0.19 (0.13 to 0.25) | 36 | 0.19 (0.12 to 0.25) | 0 (-0.09 to 0.1) | 0.914 |
| z-score | 80 | 44 | -0.04 (-0.27 to 0.2) | 36 | -0.05 (-0.31 to 0.2) | 0.02 (-0.33 to 0.37) | |
| L inferior frontal gyrus, pars opercularis | | | | | | | |
| Raw score | 80 | 44 | 0.11 (0.02 to 0.19) | 36 | 0.11 (0.02 to 0.21) | -0.01 (-0.14 to 0.12) | 0.919 |
| z-score | 80 | 44 | 0.05 (-0.26 to 0.36) | 36 | 0.07 (-0.27 to 0.42) | -0.02 (-0.49 to 0.44) | |
| L inferior frontal gyrus, pars triangularis | | | | | | | |
| Raw score | 80 | 44 | 0.13 (0.02 to 0.24) | 36 | 0.12 (-0.01 to 0.24) | 0.01 (-0.15 to 0.18) | 0.882 |
| z-score | 80 | 44 | 0.06 (-0.3 to 0.43) | 36 | 0.02 (-0.38 to 0.43) | 0.04 (-0.51 to 0.59) | |
| L middle frontal gyrus | | | | | | | |
| Raw score | 80 | 44 | -0.06 (-0.24 to 0.12) | 36 | -0.04 (-0.24 to 0.16) | -0.02 (-0.28 to 0.25) | 0.896 |
| z-score | 80 | 44 | 0.35 (-0.07 to 0.78) | 36 | 0.4 (-0.08 to 0.87) | -0.04 (-0.68 to 0.6) | |
| L superior frontal gyrus | | | | | | | |
| Raw score | 80 | 44 | 0.02 (-0.12 to 0.16) | 36 | 0.04 (-0.11 to 0.19) | -0.02 (-0.22 to 0.18) | 0.854 |
| z-score | 80 | 44 | 0.42 (0.11 to 0.72) | 36 | 0.46 (0.12 to 0.8) | -0.04 (-0.5 to 0.41) | |
| L frontal orbital cortex | | | | | | | |
| Raw score | 80 | 44 | 0.3 (0.1 to 0.49) | 36 | 0.32 (0.1 to 0.53) | -0.02 (-0.3 to 0.27) | 0.899 |
| z-score | 80 | 44 | 0.1 (-0.28 to 0.48) | 36 | 0.13 (-0.29 to 0.55) | -0.04 (-0.6 to 0.53) | |
| R cingulate gyrus, anterior division | | | | | | | |
| Raw score | 80 | 44 | 0.18 (0.13 to 0.24) | 36 | 0.16 (0.1 to 0.22) | 0.02 (-0.06 to 0.11) | 0.554 |
| z-score | 80 | 44 | -0.01 (-0.23 to 0.21) | 36 | -0.11 (-0.35 to 0.13) | 0.1 (-0.23 to 0.42) | |
| R inferior frontal gyrus, pars opercularis | | | | | | | |
| Raw score | 80 | 44 | 0.11 (0 to 0.22) | 36 | 0.13 (0.01 to 0.25) | -0.02 (-0.18 to 0.15) | 0.824 |
| z-score | 80 | 44 | -0.02 (-0.29 to 0.25) | 36 | 0.02 (-0.27 to 0.32) | -0.04 (-0.44 to 0.36) | |
| R inferior frontal gyrus, pars triangularis | | | | | | | |
| Raw score | 80 | 44 | 0.14 (0.01 to 0.27) | 36 | 0.17 (0.03 to 0.32) | -0.03 (-0.23 to 0.16) | 0.733 |
| z-score | 80 | 44 | -0.05 (-0.32 to 0.22) | 36 | 0.02 (-0.28 to 0.32) | -0.07 (-0.47 to 0.34) | |
| R middle frontal gyrus | | | | | | | |
| Raw score | 80 | 44 | -0.11 (-0.26 to 0.03) | 36 | -0.06 (-0.22 to 0.1) | -0.05 (-0.27 to 0.16) | 0.612 |
| z-score | 80 | 44 | 0.27 (-0.12 to 0.66) | 36 | 0.42 (-0.01 to 0.85) | -0.15 (-0.73 to 0.44) | |

| | | | | | | | |
|---|---|---|---|---|---|---|---|
| R superior frontal gyrus | | | | | | | |
| Raw score | 80 | 44 | -0.04 (-0.17 to 0.09) | 36 | -0.03 (-0.18 to 0.12) | -0.01 (-0.21 to 0.19) | 0.940 |
| z-score | 80 | 44 | 0.47 (0.2 to 0.75) | 36 | 0.49 (0.18 to 0.79) | -0.02 (-0.43 to 0.4) | |
| R frontal orbital cortex | | | | | | | |
| Raw score | 80 | 44 | 0.27 (0.04 to 0.5) | 36 | 0.31 (0.06 to 0.57) | -0.04 (-0.39 to 0.3) | 0.812 |
| z-score | 80 | 44 | 0.1 (-0.3 to 0.51) | 36 | 0.17 (-0.28 to 0.62) | -0.07 (-0.68 to 0.53) | |

z-score values indicate how many standard deviations have the post-exercise program values changed with respect to the baseline mean and standard deviation. e.g., a 0.50 z-score means that the mean value at post-exercise program is 0.50 standard deviations higher than the mean value at baseline, indicating a positive change, with negative values indicating the opposite. All data presented were adjusted for baseline values.

**Supplementary eTable 9**. **Per-protocol effects of the ActiveBrains exercise program on raw ($\beta$ values) and z-scores of post-exercise left anterior hippocampal functional connectivity with prefrontal cortex subregions.**

| | | | Mean (95% CI) | | | | |
|---|---|---|---|---|---|---|---|
| | Nall | N | Exercise group | N | Control group | Difference between groups | *P* |
| L cingulate gyrus, anterior division | | | | | | | |
| Raw score | 80 | 44 | 0.06 (-0.04 to 0.16) | 36 | 0.14 (0.04 to 0.25) | -0.08 (-0.23 to 0.06) | 0.266 |
| z-score | 80 | 44 | -0.35 (-0.79 to 0.08) | 36 | 0.02 (-0.47 to 0.5) | -0.37 (-1.02 to 0.28) | |
| L inferior frontal gyrus, pars opercularis | | | | | | | |
| Raw score | 80 | 44 | 0.06 (-0.02 to 0.14) | 36 | 0.08 (-0.02 to 0.16) | -0.01 (-0.14 to 0.11) | 0.813 |
| z-score | 80 | 44 | -0.04 (-0.42 to 0.34) | 36 | 0.03 (-0.39 to 0.45) | -0.07 (-0.64 to 0.5) | |
| L inferior frontal gyrus, pars triangularis | | | | | | | |
| Raw score | 80 | 44 | 0.09 (-0.02 to 0.2) | 36 | 0.08 (-0.04 to 0.2) | 0.01 (-0.15 to 0.17) | 0.928 |
| z-score | 80 | 44 | 0.01 (-0.42 to 0.44) | 36 | -0.02 (-0.49 to 0.46) | 0.03 (-0.61 to 0.67) | |
| L middle frontal gyrus | | | | | | | |
| Raw score | 80 | 44 | 0.01 (-0.16 to 0.18) | 36 | -0.02 (-0.2 to 0.17) | 0.02 (-0.23 to 0.27) | 0.863 |
| z-score | 80 | 44 | 0.3 (-0.08 to 0.67) | 36 | 0.25 (-0.16 to 0.66) | 0.05 (-0.5 to 0.6) | |
| L superior frontal gyrus | | | | | | | |
| Raw score | 80 | 44 | 0.11 (-0.01 to 0.23) | 36 | 0.08 (-0.05 to 0.22) | 0.03 (-0.16 to 0.21) | 0.768 |
| z-score | 80 | 44 | 0.5 (0.21 to 0.79) | 36 | 0.43 (0.11 to 0.76) | 0.06 (-0.37 to 0.5) | |
| L frontal orbital cortex | | | | | | | |
| Raw score | 80 | 44 | 0.32 (0.13 to 0.51) | 36 | 0.36 (0.15 to 0.56) | -0.04 (-0.32 to 0.24) | 0.790 |
| z-score | 80 | 44 | 0.06 (-0.34 to 0.47) | 36 | 0.15 (-0.31 to 0.6) | -0.08 (-0.69 to 0.53) | |
| R cingulate gyrus, anterior division | | | | | | | |
| Raw score | 80 | 44 | 0.12 (0.06 to 0.17) | 36 | 0.11 (0.05 to 0.17) | 0 (-0.08 to 0.09) | 0.912 |
| z-score | 80 | 44 | -0.02 (-0.28 to 0.23) | 36 | -0.04 (-0.33 to 0.24) | 0.02 (-0.36 to 0.4) | |
| R inferior frontal gyrus, pars opercularis | | | | | | | |
| Raw score | 80 | 44 | 0.03 (-0.06 to 0.13) | 36 | 0.1 (-0.01 to 0.2) | -0.06 (-0.21 to 0.08) | 0.399 |
| z-score | 80 | 44 | -0.07 (-0.32 to 0.18) | 36 | 0.09 (-0.19 to 0.36) | -0.16 (-0.54 to 0.22) | |
| R inferior frontal gyrus, pars triangularis | | | | | | | |
| Raw score | 80 | 44 | 0.05 (-0.06 to 0.16) | 36 | 0.14 (0.01 to 0.26) | -0.09 (-0.26 to 0.08) | 0.309 |
| z-score | 80 | 44 | -0.09 (-0.33 to 0.14) | 36 | 0.09 (-0.17 to 0.35) | -0.18 (-0.54 to 0.17) | |
| R middle frontal gyrus | | | | | | | |
| Raw score | 80 | 44 | -0.04 (-0.19 to 0.1) | 36 | -0.03 (-0.19 to 0.13) | -0.01 (-0.23 to 0.21) | 0.911 |
| z-score | 80 | 44 | 0.43 (0.02 to 0.84) | 36 | 0.46 (0.01 to 0.92) | -0.04 (-0.65 to 0.58) | |

| | | | | | | | |
|---|---|---|---|---|---|---|---|
| R superior frontal gyrus | | | | | | | |
| Raw score | 80 | 44 | 0.06 (-0.05 to 0.18) | 36 | 0.04 (-0.09 to 0.17) | 0.03 (-0.15 to 0.2) | 0.763 |
| z-score | 80 | 44 | 0.59 (0.33 to 0.84) | 36 | 0.53 (0.25 to 0.81) | 0.06 (-0.32 to 0.44) | |
| R frontal orbital cortex | | | | | | | |
| Raw score | 80 | 44 | 0.26 (0.06 to 0.46) | 36 | 0.35 (0.12 to 0.57) | -0.09 (-0.39 to 0.21) | 0.545 |
| z-score | 80 | 44 | 0.02 (-0.4 to 0.43) | 36 | 0.2 (-0.25 to 0.66) | -0.19 (-0.81 to 0.43) | |

z-score values indicate how many standard deviations have the post-exercise program values changed with respect to the baseline mean and standard deviation. e.g., a 0.50 z-score means that the mean value at post-exercise program is 0.50 standard deviations higher than the mean value at baseline, indicating a positive change, with negative values indicating the opposite. All data presented were adjusted for baseline values.

**Supplementary eTable 10**. **Per-protocol effects of the ActiveBrains exercise program on raw (*β* values) and z-scores of post-exercise left posterior hippocampal functional connectivity with prefrontal cortex subregions**

| | | | Mean (95% CI) | | | | |
|---|---|---|---|---|---|---|---|
| | Nall | N | Exercise group | N | Control group | Difference between groups | *P* |
| L cingulate gyrus, anterior division | | | | | | | |
| Raw score | 80 | 44 | 0.22 (0.16 to 0.28) | 36 | 0.22 (0.15 to 0.28) | 0 (-0.09 to 0.1) | 0.944 |
| z-score | 80 | 44 | -0.01 (-0.22 to 0.2) | 36 | -0.02 (-0.25 to 0.21) | 0.01 (-0.3 to 0.32) | |
| L inferior frontal gyrus, pars opercularis | | | | | | | |
| Raw score | 80 | 44 | 0.15 (0.07 to 0.23) | 36 | 0.14 (0.05 to 0.23) | 0.02 (-0.1 to 0.14) | 0.788 |
| z-score | 80 | 44 | 0.18 (-0.08 to 0.45) | 36 | 0.13 (-0.16 to 0.42) | 0.05 (-0.34 to 0.45) | |
| L inferior frontal gyrus, pars triangularis | | | | | | | |
| Raw score | 80 | 44 | 0.14 (0.06 to 0.23) | 36 | 0.12 (0.02 to 0.22) | 0.02 (-0.11 to 0.15) | 0.723 |
| z-score | 80 | 44 | 0.15 (-0.13 to 0.42) | 36 | 0.08 (-0.23 to 0.38) | 0.07 (-0.34 to 0.48) | |
| L middle frontal gyrus | | | | | | | |
| Raw score | 80 | 44 | -0.15 (-0.32 to 0.02) | 36 | -0.1 (-0.29 to 0.09) | -0.05 (-0.3 to 0.21) | 0.700 |
| z-score | 80 | 44 | 0.26 (-0.05 to 0.57) | 36 | 0.35 (0 to 0.69) | -0.09 (-0.55 to 0.37) | |
| L superior frontal gyrus | | | | | | | |
| Raw score | 80 | 44 | -0.13 (-0.27 to 0.01) | 36 | -0.04 (-0.19 to 0.12) | -0.09 (-0.3 to 0.11) | 0.372 |
| z-score | 80 | 44 | 0.2 (-0.05 to 0.45) | 36 | 0.37 (0.09 to 0.65) | -0.17 (-0.54 to 0.21) | |
| L frontal orbital cortex | | | | | | | |
| Raw score | 80 | 44 | 0.06 (-0.14 to 0.26) | 36 | 0.09 (-0.14 to 0.31) | -0.02 (-0.33 to 0.28) | 0.872 |
| z-score | 80 | 44 | 0.06 (-0.26 to 0.39) | 36 | 0.1 (-0.25 to 0.46) | -0.04 (-0.52 to 0.45) | |
| R cingulate gyrus, anterior division | | | | | | | |
| Raw score | 80 | 44 | 0.23 (0.17 to 0.28) | 36 | 0.19 (0.13 to 0.26) | 0.03 (-0.05 to 0.12) | 0.434 |
| z-score | 80 | 44 | 0.02 (-0.17 to 0.22) | 36 | -0.09 (-0.31 to 0.12) | 0.12 (-0.18 to 0.41) | |
| R inferior frontal gyrus, pars opercularis | | | | | | | |
| Raw score | 80 | 44 | 0.23 (0.13 to 0.32) | 36 | 0.12 (0.01 to 0.23) | 0.11 (-0.04 to 0.25) | 0.155 |
| z-score | 80 | 44 | 0.14 (-0.12 to 0.42) | 36 | -0.14 (-0.44 to 0.15) | 0.29 (-0.11 to 0.69) | |
| R inferior frontal gyrus, pars triangularis | | | | | | | |
| Raw score | 80 | 44 | 0.26 (0.13 to 0.38) | 36 | 0.14 (0 to 0.27) | 0.12 (-0.07 to 0.3) | 0.207 |
| z-score | 80 | 44 | 0.08 (-0.21 to 0.37) | 36 | -0.2 (-0.52 to 0.12) | 0.28 (-0.16 to 0.72) | |
| R middle frontal gyrus | | | | | | | |
| Raw score | 80 | 44 | -0.16 (-0.26 to -0.05) | 36 | -0.13 (-0.24 to -0.01) | -0.03 (-0.19 to 0.12) | 0.689 |
| z-score | 80 | 44 | 0.05 (-0.22 to 0.33) | 36 | 0.14 (-0.16 to 0.44) | -0.08 (-0.49 to 0.32) | |

| | | | | | | | |
|---|---|---|---|---|---|---|---|
| R superior frontal gyrus | | | | | | | |
| Raw score | 80 | 44 | -0.2 (-0.34 to -0.05) | 36 | -0.13 (-0.28 to 0.03) | -0.07 (-0.28 to 0.14) | 0.525 |
| z-score | 80 | 44 | 0.18 (-0.1 to 0.47) | 36 | 0.32 (0.01 to 0.63) | -0.14 (-0.56 to 0.29) | |
| R frontal orbital cortex | | | | | | | |
| Raw score | 80 | 44 | 0.1 (-0.17 to 0.37) | 36 | 0.11 (-0.19 to 0.4) | -0.01 (-0.41 to 0.39) | 0.973 |
| z-score | 80 | 44 | 0.15 (-0.19 to 0.5) | 36 | 0.16 (-0.22 to 0.54) | -0.01 (-0.52 to 0.51) | |

z-score values indicate how many standard deviations have the post-exercise program values changed with respect to the baseline mean and standard deviation. e.g., a 0.50 z-score means that the mean value at post-exercise program is 0.50 standard deviations higher than the mean value at baseline, indicating a positive change, with negative values indicating the opposite. All data presented were adjusted for baseline values.

**Supplementary eTable 11**. **Per-protocol effects of the ActiveBrains exercise program on raw (*β* values) and z-scores of post-exercise right hippocampal functional connectivity with prefrontal cortex subregions.**

| | | | Mean (95% CI) | | | | |
|---|---|---|---|---|---|---|---|
| | Nall | N | Exercise group | N | Control group | Difference between groups | *P* |
| L cingulate gyrus, anterior division | | | | | | | |
| Raw score | 80 | 44 | 0.19 (0.11 to 0.26) | 36 | 0.22 (0.13 to 0.3) | -0.03 (-0.14 to 0.08) | 0.590 |
| z-score | 80 | 44 | 0.03 (-0.24 to 0.31) | 36 | 0.14 (-0.16 to 0.45) | -0.11 (-0.52 to 0.3) | |
| L inferior frontal gyrus, pars opercularis | | | | | | | |
| Raw score | 80 | 44 | 0.13 (0.05 to 0.21) | 36 | 0.1 (0.01 to 0.19) | 0.03 (-0.09 to 0.15) | 0.625 |
| z-score | 80 | 44 | 0.25 (-0.04 to 0.55) | 36 | 0.14 (-0.19 to 0.47) | 0.11 (-0.33 to 0.55) | |
| L inferior frontal gyrus, pars triangularis | | | | | | | |
| Raw score | 80 | 44 | 0.16 (0.06 to 0.26) | 36 | 0.11 (0 to 0.22) | 0.04 (-0.1 to 0.19) | 0.545 |
| z-score | 80 | 44 | 0.26 (-0.06 to 0.58) | 36 | 0.11 (-0.24 to 0.47) | 0.15 (-0.34 to 0.63) | |
| L middle frontal gyrus | | | | | | | |
| Raw score | 80 | 44 | -0.11 (-0.27 to 0.05) | 36 | 0.01 (-0.17 to 0.19) | -0.12 (-0.36 to 0.12) | 0.311 |
| z-score | 80 | 44 | 0.22 (-0.16 to 0.59) | 36 | 0.5 (0.09 to 0.91) | -0.28 (-0.84 to 0.27) | |
| L superior frontal gyrus | | | | | | | |
| Raw score | 80 | 44 | -0.05 (-0.16 to 0.06) | 36 | 0.05 (-0.07 to 0.17) | -0.1 (-0.26 to 0.06) | 0.220 |
| z-score | 80 | 44 | 0.3 (0.03 to 0.57) | 36 | 0.55 (0.25 to 0.84) | -0.25 (-0.65 to 0.15) | |
| L frontal orbital cortex | | | | | | | |
| Raw score | 80 | 44 | 0.22 (0.04 to 0.4) | 36 | 0.34 (0.14 to 0.54) | -0.13 (-0.4 to 0.14) | 0.356 |
| z-score | 80 | 44 | -0.03 (-0.41 to 0.35) | 36 | 0.23 (-0.18 to 0.65) | -0.26 (-0.83 to 0.3) | |
| R cingulate gyrus, anterior division | | | | | | | |
| Raw score | 80 | 44 | 0.18 (0.11 to 0.25) | 36 | 0.2 (0.12 to 0.28) | -0.02 (-0.12 to 0.09) | 0.756 |
| z-score | 80 | 44 | 0.04 (-0.23 to 0.3) | 36 | 0.1 (-0.2 to 0.4) | -0.06 (-0.46 to 0.34) | |
| R inferior frontal gyrus, pars opercularis | | | | | | | |
| Raw score | 80 | 44 | 0.12 (0.02 to 0.24) | 36 | 0.15 (0.03 to 0.27) | -0.02 (-0.19 to 0.14) | 0.769 |
| z-score | 80 | 44 | 0.01 (-0.31 to 0.32) | 36 | 0.08 (-0.27 to 0.43) | -0.07 (-0.54 to 0.4) | |
| R inferior frontal gyrus, pars triangularis | | | | | | | |
| Raw score | 80 | 44 | 0.14 (0.01 to 0.27) | 36 | 0.19 (0.05 to 0.33) | -0.05 (-0.24 to 0.15) | 0.636 |
| z-score | 80 | 44 | -0.03 (-0.35 to 0.29) | 36 | 0.08 (-0.27 to 0.44) | -0.11 (-0.59 to 0.36) | |
| R middle frontal gyrus | | | | | | | |
| Raw score | 80 | 44 | -0.16 (-0.28 to -0.04) | 36 | 0 (-0.14 to 0.13) | -0.16 (-0.34 to 0.03) | 0.092 |
| z-score | 80 | 44 | 0.05 (-0.31 to 0.4) | 36 | 0.5 (0.11 to 0.89) | -0.45 (-0.98 to 0.08) | |

| | | | | | | | |
|---|---|---|---|---|---|---|---|
| R superior frontal gyrus | | | | | | | |
| Raw score | 80 | 44 | -0.08 (-0.19 to 0.03) | 36 | 0.01 (-0.11 to 0.13) | -0.09 (-0.25 to 0.07) | 0.268 |
| z-score | 80 | 44 | 0.34 (0.1 to 0.58) | 36 | 0.54 (0.27 to 0.8) | -0.2 (-0.56 to 0.16) | |
| R frontal orbital cortex | | | | | | | |
| Raw score | 80 | 44 | 0.3 (0.1 to 0.51) | 36 | 0.39 (0.16 to 0.61) | -0.08 (-0.38 to 0.22) | 0.596 |
| z-score | 80 | 44 | 0.14 (-0.28 to 0.55) | 36 | 0.3 (-0.16 to 0.76) | -0.16 (-0.78 to 0.45) | |

z-score values indicate how many standard deviations have the post-exercise program values changed with respect to the baseline mean and standard deviation. e.g., a 0.50 z-score means that the mean value at post-exercise program is 0.50 standard deviations higher than the mean value at baseline, indicating a positive change, with negative values indicating the opposite. All data presented were adjusted for baseline values.

**Supplementary eTable 12. Per-protocol effects of the ActiveBrains exercise program on raw (*β* values) and z-scores of post-exercise right anterior hippocampal functional connectivity with prefrontal cortex subregions.**

| | Mean (95% CI) | | | | | | |
|---|---|---|---|---|---|---|---|
| | Nall | N | Exercise group | N | Control group | Difference between groups | *P* |
| L cingulate gyrus, anterior division | | | | | | | |
| Raw score | 80 | 44 | 0.13 (0.05 to 0.2) | 36 | 0.16 (0.08 to 0.24) | -0.04 (-0.14 to 0.08) | 0.528 |
| z-score | 80 | 44 | 0 (-0.31 to 0.3) | 36 | 0.14 (-0.19 to 0.47) | -0.14 (-0.59 to 0.31) | |
| L inferior frontal gyrus, pars opercularis | | | | | | | |
| Raw score | 80 | 44 | 0.08 (0.01 to 0.16) | 36 | 0.06 (-0.02 to 0.15) | 0.02 (-0.09 to 0.14) | 0.679 |
| z-score | 80 | 44 | 0.26 (-0.08 to 0.59) | 36 | 0.15 (-0.21 to 0.52) | 0.1 (-0.39 to 0.6) | |
| L inferior frontal gyrus, pars triangularis | | | | | | | |
| Raw score | 80 | 44 | 0.11 (0.01 to 0.21) | 36 | 0.08 (-0.04 to 0.19) | 0.04 (-0.11 to 0.19) | 0.622 |
| z-score | 80 | 44 | 0.27 (-0.1 to 0.64) | 36 | 0.14 (-0.27 to 0.54) | 0.14 (-0.41 to 0.69) | |
| L middle frontal gyrus | | | | | | | |
| Raw score | 80 | 44 | -0.1 (-0.26 to 0.06) | 36 | 0.05 (-0.13 to 0.22) | -0.15 (-0.38 to 0.09) | 0.223 |
| z-score | 80 | 44 | 0.11 (-0.26 to 0.48) | 36 | 0.45 (0.04 to 0.86) | -0.34 (-0.89 to 0.21) | |
| L superior frontal gyrus | | | | | | | |
| Raw score | 80 | 44 | 0 (-0.11 to 0.1) | 36 | 0.1 (-0.02 to 0.21) | -0.1 (-0.26 to 0.05) | 0.189 |
| z-score | 80 | 44 | 0.29 (0.02 to 0.55) | 36 | 0.55 (0.26 to 0.84) | -0.26 (-0.66 to 0.13) | |
| L frontal orbital cortex | | | | | | | |
| Raw score | 80 | 44 | 0.22 (0.04 to 0.41) | 36 | 0.36 (0.16 to 0.57) | -0.14 (-0.42 to 0.14) | 0.311 |
| z-score | 80 | 44 | -0.14 (-0.55 to 0.28) | 36 | 0.18 (-0.28 to 0.63) | -0.31 (-0.93 to 0.3) | |
| R cingulate gyrus, anterior division | | | | | | | |
| Raw score | 80 | 44 | 0.12 (0.05 to 0.19) | 36 | 0.14 (0.07 to 0.22) | -0.03 (-0.13 to 0.08) | 0.598 |
| z-score | 80 | 44 | -0.04 (-0.34 to 0.26) | 36 | 0.08 (-0.25 to 0.41) | -0.12 (-0.56 to 0.33) | |
| R inferior frontal gyrus, pars opercularis | | | | | | | |
| Raw score | 80 | 44 | 0.04 (-0.06 to 0.15) | 36 | 0.09 (-0.02 to 0.21) | -0.05 (-0.21 to 0.11) | 0.537 |
| z-score | 80 | 44 | -0.05 (-0.37 to 0.26) | 36 | 0.1 (-0.26 to 0.45) | -0.15 (-0.62 to 0.33) | |
| R inferior frontal gyrus, pars triangularis | | | | | | | |
| Raw score | 80 | 44 | 0.08 (-0.05 to 0.2) | 36 | 0.15 (0.01 to 0.29) | -0.07 (-0.26 to 0.12) | 0.449 |
| z-score | 80 | 44 | -0.05 (-0.37 to 0.27) | 36 | 0.13 (-0.22 to 0.48) | -0.18 (-0.66 to 0.29) | |
| R middle frontal gyrus | | | | | | | |
| Raw score | 80 | 44 | -0.16 (-0.3 to -0.03) | 36 | 0.03 (-0.12 to 0.18) | -0.19 (-0.39 to 0) | 0.055 |
| z-score | 80 | 44 | -0.08 (-0.45 to 0.3) | 36 | 0.48 (0.06 to 0.89) | -0.55 (-1.11 to 0.01) | |

| | | | | | | | |
|---|---|---|---|---|---|---|---|
| R superior frontal gyrus | | | | | | | |
| Raw score | 80 | 44 | -0.02 (-0.13 to 0.09) | 36 | 0.08 (-0.04 to 0.2) | -0.1 (-0.26 to 0.06) | 0.231 |
| z-score | 80 | 44 | 0.36 (0.09 to 0.62) | 36 | 0.6 (0.3 to 0.89) | -0.24 (-0.63 to 0.16) | |
| R frontal orbital cortex | | | | | | | |
| Raw score | 80 | 44 | 0.34 (0.16 to 0.51) | 36 | 0.4 (0.2 to 0.59) | -0.06 (-0.32 to 0.2) | 0.650 |
| z-score | 80 | 44 | 0.11 (-0.27 to 0.49) | 36 | 0.24 (-0.18 to 0.66) | -0.13 (-0.7 to 0.44) | |

z-score values indicate how many standard deviations have the post-exercise program values changed with respect to the baseline mean and standard deviation. e.g., a 0.50 z-score means that the mean value at post-exercise program is 0.50 standard deviations higher than the mean value at baseline, indicating a positive change, with negative values indicating the opposite. All data presented were adjusted for baseline values.

**Supplementary eTable 13. Per-protocol effects of the ActiveBrains exercise program on raw (*β* values) and z-scores of post-exercise right posterior hippocampal functional connectivity with prefrontal cortex subregions.**

| | Mean (95% CI) | | | | | | |
|---|---|---|---|---|---|---|---|
| | Nall | N | Exercise group | N | Control group | Difference between groups | *P* |
| L cingulate gyrus, anterior division | | | | | | | |
| Raw score | 80 | 44 | 0.22 (0.16 to 0.29) | 36 | 0.22 (0.15 to 0.3) | 0 (-0.1 to 0.1) | |
| z-score | 80 | 44 | 0.08 (-0.15 to 0.32) | 36 | 0.08 (-0.18 to 0.34) | 0 (-0.35 to 0.35) | 0.992 |
| L inferior frontal gyrus, pars opercularis | | | | | | | |
| Raw score | 80 | 44 | 0.17 (0.09 to 0.25) | 36 | 0.14 (0.05 to 0.23) | 0.03 (-0.1 to 0.15) | |
| z-score | 80 | 44 | 0.22 (-0.06 to 0.51) | 36 | 0.13 (-0.18 to 0.44) | 0.09 (-0.33 to 0.51) | 0.660 |
| L inferior frontal gyrus, pars triangularis | | | | | | | |
| Raw score | 80 | 44 | 0.2 (0.11 to 0.28) | 36 | 0.13 (0.03 to 0.23) | 0.06 (-0.07 to 0.2) | |
| z-score | 80 | 44 | 0.21 (-0.07 to 0.49) | 36 | 0.01 (-0.3 to 0.32) | 0.2 (-0.21 to 0.62) | 0.333 |
| L middle frontal gyrus | | | | | | | |
| Raw score | 80 | 44 | -0.06 (-0.26 to 0.14) | 36 | -0.12 (-0.34 to 0.1) | 0.05 (-0.24 to 0.35) | |
| z-score | 80 | 44 | 0.39 (-0.1 to 0.88) | 36 | 0.26 (-0.28 to 0.8) | 0.13 (-0.59 to 0.86) | 0.717 |
| L superior frontal gyrus | | | | | | | |
| Raw score | 80 | 44 | -0.08 (-0.22 to 0.06) | 36 | -0.1 (-0.26 to 0.06) | 0.02 (-0.19 to 0.23) | |
| z-score | 80 | 44 | 0.32 (-0.02 to 0.66) | 36 | 0.27 (-0.1 to 0.65) | 0.04 (-0.46 to 0.55) | 0.859 |
| L frontal orbital cortex | | | | | | | |
| Raw score | 80 | 44 | 0.16 (-0.05 to 0.37) | 36 | 0.16 (-0.08 to 0.38) | 0 (-0.31 to 0.32) | |
| z-score | 80 | 44 | 0.18 (-0.17 to 0.54) | 36 | 0.18 (-0.21 to 0.56) | 0.01 (-0.52 to 0.53) | 0.976 |
| R cingulate gyrus, anterior division | | | | | | | |
| Raw score | 80 | 44 | 0.23 (0.16 to 0.29) | 36 | 0.21 (0.14 to 0.28) | 0.02 (-0.08 to 0.11) | |
| z-score | 80 | 44 | 0.15 (-0.08 to 0.38) | 36 | 0.08 (-0.17 to 0.34) | 0.07 (-0.28 to 0.41) | 0.693 |
| R inferior frontal gyrus, pars opercularis | | | | | | | |
| Raw score | 80 | 44 | 0.22 (0.12 to 0.33) | 36 | 0.18 (0.06 to 0.3) | 0.04 (-0.11 to 0.2) | |
| z-score | 80 | 44 | 0.18 (-0.15 to 0.5) | 36 | 0.04 (-0.32 to 0.4) | 0.14 (-0.36 to 0.63) | 0.587 |
| R inferior frontal gyrus, pars triangularis | | | | | | | |
| Raw score | 80 | 44 | 0.2 (0.08 to 0.32) | 36 | 0.17 (0.04 to 0.31) | 0.03 (-0.16 to 0.21) | |
| z-score | 80 | 44 | 0.03 (-0.3 to 0.36) | 36 | -0.04 (-0.41 to 0.32) | 0.08 (-0.42 to 0.57) | 0.766 |
| R middle frontal gyrus | | | | | | | |

| | | | | | | | |
|---|---|---|---|---|---|---|---|
| Raw score | 80 | 44 | -0.06 (-0.2 to 0.08) | 36 | -0.09 (-0.24 to 0.06) | 0.03 (-0.18 to 0.23) | |
| z-score | 80 | 44 | 0.32 (-0.04 to 0.69) | 36 | 0.25 (-0.16 to 0.65) | 0.08 (-0.46 to 0.62) | 0.776 |
| R superior frontal gyrus | | | | | | | |
| Raw score | 80 | 44 | -0.1 (-0.24 to 0.04) | 36 | -0.14 (-0.3 to 0.01) | 0.04 (-0.17 to 0.25) | |
| z-score | 80 | 44 | 0.36 (0.04 to 0.67) | 36 | 0.26 (-0.08 to 0.61) | 0.09 (-0.38 to 0.56) | 0.692 |
| R frontal orbital cortex | | | | | | | |
| Raw score | 80 | 44 | 0.15 (-0.1 to 0.4) | 36 | 0.23 (-0.04 to 0.51) | -0.08 (-0.45 to 0.29) | |
| z-score | 80 | 44 | 0.16 (-0.21 to 0.52) | 36 | 0.28 (-0.13 to 0.68) | -0.12 (-0.66 to 0.43) | 0.668 |

z-score values indicate how many standard deviations have the post-exercise program values changed with respect to the baseline mean and standard deviation. e.g., a 0.50 z-score means that the mean value at post-exercise program is 0.50 standard deviations higher than the mean value at baseline, indicating a positive change, with negative values indicating the opposite. All data presented were adjusted for baseline values.

**Supplementary eTable 14. Per-protocol effects of the ActiveBrains exercise program on raw (mm3) and z-scores of post-exercise subcortical brain volumes others than the hippocampus (see Supplementary Table 4 for exercise effects analysis on hippocampus).**

| | | | Mean (95% CI) | | | | |
|---|---|---|---|---|---|---|---|
| | Nall | N | Exercise group | N | Control group | Difference between groups | *P* |
| Right accumbens | | | | | | | |
| Raw score | 82 | 45 | 456.25 (432.53 to 479.96) | 37 | 459.14 (432.99 to 485.30) | -2.90 (–38.22 to 32.43) | 0.871 |
| z-score | 82 | 45 | –0.14 (–0.38 to 0.10) | 37 | –0.11 (–0.37 to 0.16) | –0.03 (–0.39 to 0.33) | |
| Left accumbens | | | | | | | |
| Raw score | 82 | 45 | 552.35 (532.02 to 572.68) | 37 | 542.51 (520.08 to 564.93) | 9.84 (–20.47 to 40.15) | 0.520 |
| z-score | 82 | 45 | 0.00 (–0.18 to 0.18) | 37 | –0.09 (–0.29 to 0.11) | 0.09 (–0.18 to 0.36) | |
| Right amygdala | | | | | | | |
| Raw score | 80 | 44 | 1374.93 (1317.82 to 1432.03) | 36 | 1371.33 (1308.19 to 1434.46) | 3.60 (–81.62 to 88.83) | 0.933 |
| z-score | 80 | 44 | 0.18 (–0.08 to 0.44) | 36 | 0.17 (–0.12 to 0.45) | 0.02 (–0.37 to 0.40) | |
| Left Amygdala | | | | | | | |
| Raw score | 82 | 45 | 1370.04 (1323.15 to 1416.93) | 37 | 1339.44 (1287.72 to 1391.16) | 30.60 (–39.28 to 100.48) | 0.386 |
| z-score | 82 | 45 | 0.26 (0.00 to 0.51) | 37 | 0.09 (–0.19 to 0.37) | 0.16 (–0.21 to 0.54) | |
| Right caudate | | | | | | | |
| Raw score | 82 | 45 | 3826.26 (3749.00 to 3903.53) | 37 | 3857.53 (3772.25 to 3942.80) | -31.26 (–146.77 to 84.25) | 0.592 |
| z-score | 82 | 45 | 0.03 (–0.12 to 0.18) | 37 | 0.09 (–0.08 to 0.25) | –0.06 (–0.29 to 0.16) | |
| Left caudate | | | | | | | |
| Raw score | 81 | 44 | 3811.92 (3747.60 to 3876.23) | 37 | 3827.27 (3757.13 to 3897.42) | -15.36 (–110.56 to 79.85) | 0.749 |
| z-score | 81 | 44 | –0.01 (–0.16 to 0.14) | 37 | 0.03 (–0.14 to 0.19) | –0.04 (–0.26 to 0.19) | |
| Right pallidum | | | | | | | |
| Raw score | 82 | 45 | 1657.46 (1638.84 to 1676.07) | 37 | 1656.65 (1636.12 to 1677.19) | 0.80 (–26.94 to 28.55) | 0.954 |
| z-score | 82 | 45 | 0.03 (–0.09 to 0.14) | 37 | 0.02 (–0.11 to 0.15) | 0.00 (–0.17 to 0.18) | |
| Left pallidum | | | | | | | |
| Raw score | 82 | 45 | 1671.11 (1649.04 to 1693.17) | 37 | 1667.33 (1642.99 to 1691.66) | 3.78 (–29.07 to 36.63) | 0.819 |
| z-score | 82 | 45 | 0.11 (–0.02 to 0.24) | 37 | 0.09 (–0.06 to 0.24) | 0.02 (–0.18 to 0.22) | |
| Right putamen | | | | | | | |

| | | | | | | | |
|---|---|---|---|---|---|---|---|
| Raw score | 82 | 45 | 4989.89 (4937.53 to 5042.24) | 37 | 4979.32 (4921.58 to 5037.06) | 10.57 (–67.39 to 88.52) | 0.788 |
| z-score | 82 | 45 | –0.03 (–0.12 to 0.07) | 37 | –0.05 (–0.15 to 0.06) | 0.02 (–0.12 to 0.16) | |
| Left Putamen | | | | | | | |
| Raw score | 82 | 45 | 4977.08 (4918.12 to 5036.04) | 37 | 4949.99 (4884.97 to 5015.01) | 27.09 (–60.69 to 114.86) | 0.541 |
| z-score | 82 | 45 | 0.00 (–0.11 to 0.12) | 37 | –0.05 (–0.18 to 0.08) | 0.05 (–0.12 to 0.23) | |
| Right thalamus | | | | | | | |
| Raw score | 82 | 45 | 7843.66 (7783.18 to 7904.15) | 37 | 7772.54 (7705.81 to 7839.27) | 71.12 (–19.11 to 161.36) | 0.121 |
| z-score | 82 | 45 | 0.07 (–0.02 to 0.16) | 37 | –0.04 (–0.14 to 0.06) | 0.11 (–0.03 to 0.24) | |
| Left thalamus | | | | | | | |
| Raw score | 82 | 45 | 7920.44 (7868.97 to 7971.92) | 37 | 7928.20 (7871.43 to 7984.97) | –7.75 (–84.42 to 68.91) | 0.841 |
| z-score | 82 | 45 | 0.02 (–0.06 to 0.10) | 37 | 0.03 (–0.06 to 0.12) | –0.01 (–0.13 to 0.11) | |
| Brain Stem | | | | | | | |
| Raw score | 82 | 45 | 18988.27 (18686.42 to 19290.12) | 37 | 18909.90 (18577.01 to 19242.79) | 78.37 (–371.05 to 527.78) | 0.729 |
| z-score | 82 | 45 | 0.09 (–0.05 to 0.24) | 37 | 0.06 (–0.11 to 0.22) | 0.04 (–0.18 to 0.26) | |

z-score values indicate how many standard deviations have the post-exercise program values changed with respect to the baseline mean and standard deviation. e.g., a 0.50 z-score means that the mean value at post-exercise program is 0.50 standard deviations higher than the mean value at baseline, indicating a positive change, with negative values indicating the opposite. All data presented were adjusted for baseline values of the outcome studied.

**Supplementary eTable 15**. **Per-protocol effects of the ActiveBrains exercise program on raw ($cm^3$) and z-scores of post-exercise total brain volumes.**

| | | | Mean (95% CI) | | | | |
|---|---|---|---|---|---|---|---|
| | Nall | N | Exercise group | N | Control group | Difference between groups | *P* |
| Total gray matter | | | | | | | |
| Raw score | 82 | 45 | 795.36 (790.24 to 800.49) | 37 | 799.32 (793.66 to 804.97) | –3.96 (–11.59 to 3.68) | 0.305 |
| z-score | 82 | 45 | –0.10 (–0.18 to –0.02) | 37 | –0.04 (–0.12 to 0.05) | –0.06 (–0.18 to 0.06) | |
| Total white matter | | | | | | | |
| Raw score | 82 | 45 | 415.10 (413.33 to 416.87) | 37 | 415.45 (413.49 to 417.4) | –0.34 (–2.98 to 2.29) | 0.796 |
| z-score | 82 | 45 | 0.08 (0.05 to 0.12) | 37 | 0.09 (0.05 to 0.13) | –0.01 (–0.06 to 0.05) | |
| Total brain volume | | | | | | | |
| Raw score | 82 | 45 | 1210.42 (1204.91 to 1215.93) | 37 | 1214.82 (1208.74 to 1220.89) | –4.39 (–12.6 to 3.81) | 0.290 |
| z-score | 82 | 45 | –0.02 (–0.07 to 0.03) | 37 | 0.02 (–0.04 to 0.08) | –0.04 (–0.12 to 0.04) | |

z-score values indicate how many standard deviations have the post-exercise program values changed with respect to the baseline mean and standard deviation. e.g., a 0.50 z-score means that the mean value at post-exercise program is 0.50 standard deviations higher than the mean value at baseline, indicating a positive change, with negative values indicating the opposite. All data presented were adjusted for baseline values of the outcome studied.

**Supplementary eTable 16**. **Per-protocol effects of the ActiveBrains exercise program on raw (loadings) and z-scores of post-exercise structural covariance network.**

| | | | Mean (95% CI) | | | | |
|---|---|---|---|---|---|---|---|
| Value | Nall | N | Exercise group | N | Control group | Difference between groups | *P* |
| Network 1 | | | | | | | |
| Raw score | 82 | 45 | 95.42 (94.81 to 96.04) | 37 | 95.86 (95.17 to 96.54) | -0.43 (-1.35 to 0.49) | 0.352 |
| z-score | 82 | 45 | -0.05 (-0.13 to 0.03) | 37 | 0.01 (-0.08 to 0.09) | -0.06 (-0.17 to 0.06) | |
| Network 2 | | | | | | | |
| Raw score | 82 | 45 | 92.54 (91.38 to 93.7) | 37 | 93.85 (92.57 to 95.13) | -1.31 (-3.04 to 0.42) | 0.134 |
| z-score | 82 | 45 | -0.15 (-0.27 to -0.03) | 37 | -0.02 (-0.15 to 0.11) | -0.13 (-0.31 to 0.04) | |
| Network 3 | | | | | | | |
| Raw score | 82 | 45 | 91.66 (91.07 to 92.26) | 37 | 92.09 (91.44 to 92.75) | -0.43 (-1.32 to 0.46) | 0.337 |
| z-score | 82 | 45 | -0.05 (-0.13 to 0.03) | 37 | 0.01 (-0.08 to 0.1) | -0.06 (-0.18 to 0.06) | |
| Network 4 | | | | | | | |
| Raw score | 82 | 45 | 87.03 (86.29 to 87.78) | 37 | 87.47 (86.65 to 88.29) | -0.43 (-1.54 to 0.67) | 0.438 |
| z-score | 82 | 45 | -0.16 (-0.25 to -0.07) | 37 | -0.11 (-0.21 to -0.01) | -0.05 (-0.18 to 0.08) | |
| Network 5 | | | | | | | |
| Raw score | 82 | 45 | 81.46 (80.89 to 82.04) | 37 | 81.85 (81.22 to 82.48) | -0.39 (-1.24 to 0.47) | 0.369 |
| z-score | 82 | 45 | -0.08 (-0.16 to -0.01) | 37 | -0.03 (-0.12 to 0.05) | -0.05 (-0.16 to 0.06) | |
| Network 6 | | | | | | | |
| Raw score | 82 | 45 | 80.23 (79.73 to 80.73) | 37 | 80.85 (80.3 to 81.41) | -0.62 (-1.37 to 0.12) | 0.100 |
| z-score | 82 | 45 | -0.1 (-0.16 to -0.03) | 37 | -0.02 (-0.09 to 0.05) | -0.08 (-0.18 to 0.02) | |
| Network 7 | | | | | | | |
| Raw score | 82 | 45 | 76.93 (76.38 to 77.49) | 37 | 77.2 (76.59 to 77.81) | -0.26 (-1.09 to 0.56) | 0.529 |
| z-score | 82 | 45 | -0.13 (-0.2 to -0.06) | 37 | -0.1 (-0.17 to -0.02) | -0.03 (-0.14 to 0.07) | |
| Network 8 | | | | | | | |
| Raw score | 82 | 45 | 76.12 (75.55 to 76.69) | 37 | 76.24 (75.61 to 76.87) | -0.12 (-0.96 to 0.73) | 0.785 |
| z-score | 82 | 45 | -0.11 (-0.19 to -0.03) | 37 | -0.09 (-0.18 to 0) | -0.02 (-0.13 to 0.1) | |
| Network 9 | | | | | | | |
| Raw score | 82 | 45 | 68.64 (68.19 to 69.09) | 37 | 68.92 (68.43 to 69.42) | -0.28 (-0.95 to 0.39) | 0.408 |
| z-score | 82 | 45 | -0.15 (-0.23 to -0.07) | 37 | -0.1 (-0.19 to -0.02) | -0.05 (-0.17 to 0.07) | |
| Network 10 | | | | | | | |
| Raw score | 82 | 45 | 68.52 (68.02 to 69.02) | 37 | 68.73 (68.18 to 69.28) | -0.21 (-0.96 to 0.53) | 0.571 |
| z-score | 82 | 45 | -0.12 (-0.2 to -0.05) | 37 | -0.09 (-0.17 to -0.01) | -0.03 (-0.14 to 0.08) | |
| Network 11 | | | | | | | |
| Raw score | 82 | 45 | 68.41 (67.9 to 68.92) | 37 | 68.67 (68.11 to 69.23) | -0.26 (-1.02 to 0.5) | 0.497 |
| z-score | 82 | 45 | -0.14 (-0.22 to -0.06) | 37 | -0.1 (-0.19 to -0.02) | -0.04 (-0.16 to 0.08) | |
| Network 12 | | | | | | | |
| Raw score | 82 | 45 | 65.56 (65.07 to 66.04) | 37 | 66.14 (65.6 to 66.67) | -0.58 (-1.31 to 0.14) | 0.112 |
| z-score | 82 | 45 | -0.14 (-0.21 to -0.07) | 37 | -0.05 (-0.13 to 0.02) | -0.08 (-0.19 to 0.02) | |
| Network 13 | | | | | | | |
| Raw score | 82 | 45 | 65.87 (65.39 to 66.34) | 37 | 66.13 (65.61 to 66.66) | -0.27 (-0.98 to 0.44) | 0.456 |
| z-score | 82 | 45 | -0.14 (-0.22 to -0.06) | 37 | -0.09 (-0.18 to 0) | -0.04 (-0.17 to 0.08) | |
| Network 14 | | | | | | | |
| Raw score | 82 | 45 | 61.4 (61.02 to 61.78) | 37 | 61.6 (61.18 to 62.02) | -0.2 (-0.77 to 0.36) | 0.476 |
| z-score | 82 | 45 | -0.13 (-0.19 to -0.07) | 37 | -0.1 (-0.16 to -0.03) | -0.03 (-0.12 to 0.06) | |
| Network 15 | | | | | | | |
| Raw score | 82 | 45 | 62.95 (62.54 to 63.37) | 37 | 63.23 (62.77 to 63.68) | -0.28 (-0.89 to 0.34) | 0.379 |
| z-score | 82 | 45 | -0.16 (-0.22 to -0.1) | 37 | -0.12 (-0.19 to -0.05) | -0.04 (-0.13 to 0.05) | |
| Network 16 | | | | | | | |
| Raw score | 82 | 45 | 60.09 (59.73 to 60.44) | 37 | 60.55 (60.16 to 60.94) | -0.46 (-0.98 to 0.07) | 0.087 |
| z-score | 82 | 45 | -0.19 (-0.26 to -0.13) | 37 | -0.11 (-0.18 to -0.03) | -0.09 (-0.19 to 0.01) | |
| Network 17 | | | | | | | |

| | | | | | | | |
|---|---|---|---|---|---|---|---|
| Raw score | 82 | 45 | 64.35 (63.94 to 64.76) | 37 | 64.59 (64.14 to 65.04) | -0.24 (-0.85 to 0.37) | 0.439 |
| z-score | 82 | 45 | -0.14 (-0.22 to -0.06) | 37 | -0.09 (-0.18 to -0.01) | -0.04 (-0.16 to 0.07) | |
| Network 18 | | | | | | | |
| Raw score | 82 | 45 | 62.01 (61.59 to 62.42) | 37 | 62.28 (61.83 to 62.74) | -0.28 (-0.89 to 0.34) | 0.376 |
| z-score | 82 | 45 | -0.11 (-0.18 to -0.04) | 37 | -0.06 (-0.14 to 0.02) | -0.05 (-0.15 to 0.06) | |
| Network 19 | | | | | | | |
| Raw score | 82 | 45 | 57.31 (57.02 to 57.59) | 37 | 57.66 (57.35 to 57.98) | -0.36 (-0.78 to 0.07) | 0.100 |
| z-score | 82 | 45 | -0.14 (-0.22 to -0.07) | 37 | -0.05 (-0.13 to 0.03) | -0.09 (-0.2 to 0.02) | |
| Network 20 | | | | | | | |
| Raw score | 82 | 45 | 53.24 (52.9 to 53.59) | 37 | 53.21 (52.83 to 53.58) | 0.04 (-0.47 to 0.55) | 0.885 |
| z-score | 82 | 45 | -0.16 (-0.25 to -0.08) | 37 | -0.17 (-0.26 to -0.08) | 0.01 (-0.12 to 0.13) | |

z-score values indicate how many standard deviations have the post-exercise program values changed with respect to the baseline mean and standard deviation. e.g., a 0.50 z-score means that the mean value at post-exercise program is 0.50 standard deviations higher than the mean value at baseline, indicating a positive change, with negative values indicating the opposite. All data presented were adjusted for baseline values of the outcome studied. The names of each network based on the brain region covered is provided in **Fig. 5**.

**Supplementary eTable 17. Per-protocol effects of the *ActiveBrains* exercise program on raw and z-scores of post-exercise cardiorespiratory fitness**.

| | Mean (95% CI) | | | | | | |
|---|---|---|---|---|---|---|---|
| | $N_{all}$ | N | Exercise group | N | Control group | Difference between groups | *P* |
| Time-to-exhaustion in the maximal incremental test | 90 | 47 | | 43 | | | |
| Raw score (min) | | | 10.00 (9.24 to 10.77) | | 8.84 (8.05 to 9.64) | 1.15 (0.03 to 2.29) | **0.044** |
| z-score | | | 0.54 (0.27 to 0.82) | | 0.13 (–0.16 to 0.41) | 0.42 (0.01 to 0.82) | |
| Peak Oxygen Consumption, $VO_2$peak | 90 | 47 | | 43 | | | |
| Raw score (mL/kg/min) | | | 39.13 (37.89 to 40.37) | | 37.72 (36.42 to 39.02) | 1.40 (–0.41 to 3.23) | 0.129 |
| z-score | | | 0.39 (0.13 to 0.65) | | 0.10 (–0.18 to 0.37) | 0.29 (–0.08 to 0.67) | |

z-score values indicate how many standard deviations have the post-exercise program values changed with respect to the baseline mean and standard deviation. e.g., a 0.50 z-score means that the mean value at post-exercise program is 0.50 standard deviations higher than the mean value at baseline, indicating a positive change, with negative values indicating the opposite.
All data presented were adjusted for baseline values of the outcome studied.

**Supplementary eTable 18**. **Sex differences in intensity monitored by heart rate during the exercise sessions**.

| | Boys (N = 31) | Girls (N = 16) | Cohen's d | P |
|---|---|---|---|---|
| Session duration (min) | 66.19 ± 2.01 | 65.21 ± 1.92 | 0.49 | 0.112 |
| Mean HR (bpm) | 137.32 ± 8.52 | 138.20 ± 6.83 | 0.11 | 0.725 |
| Mean HR expressed as percentage of MaxHR (%) | 69.86 ± 4.99 | 69.22 ± 2.73 | 0.15 | 0.636 |
| Time above 80% of MaxHR (min) | 25.97 ± 10.23 | 23.89 ± 10.42 | 0.20 | 0.516 |
| Percentage of time above 80% of MaxHR (%) | 39.26 ± 15.91 | 36.64 ± 16.00 | 0.29 | 0.596 |
| Time above anaerobic threshold (min) | 13.18 ± 10.15 | 4.91 ± 3.84 | 0.96 | **0.003** |
| Percentage of time above anaerobic threshold (%) | 19.80 ± 15.34 | 7.48 ± 5.82 | 0.95 | **0.003** |

Values are expressed as means ± standard deviations (SD). Differences between sexes were determined by one-way analysis of variance (ANOVA). Statistically significant values at $P < 0.005$ are shown in bold. Cohen's effect size statistics (d) are also reported and interpreted as small (d = 0.2), medium (d = 0.5), and large (d= 0.8). The intensity of the exercise program was monitored in every child and in every session using a heart rate monitor (POLAR RS300X, Polar Electro Oy Inc., Kempele, Finland). The intensity variables shown in the table rate data represented the whole exercise session (i.e., both the aerobic exercise and resistance training). HR = Heart rate.

**Supplementary eTable 19. Intention-to-treat effects of the *ActiveBrains* exercise program on raw and z-scores of post-exercise intelligence and executive function outcomes.**

| | | | Mean (95% CI) | | | | |
|---|---|---|---|---|---|---|---|
| | $N_{all}$ | N | Exercise group | N | Control group | Difference between groups | *P* |
| Crystallized intelligence | 109 | 57 | | 52 | | | |
| Raw score (typical punctuation) | | | 109.78 (107.73 to 111.83) | | 102.13 (99.96 to 104.3) | 7.65 (4.66 to 10.63) | **0.00002** |
| z-score | | | 0.50 (0.34 to 0.67) | | –0.04 (–0.21 to 0.14) | 0.54 (0.30 to 0.77) | |
| Fluid intelligence | 109 | 57 | | 52 | | | |
| Raw score (typical punctuation) | | | 103.84 (100.96 to 106.71) | | 101.17 (98.17 to 104.18) | 2.66 (–1.51 to 6.83) | 0.209 |
| z-score | | | 0.47 (0.25 to 0.69) | | 0.27 (0.04 to 0.50) | 0.20 (–0.12 to 0.52) | |
| Total intelligence | 109 | 57 | | 52 | | | |
| Raw score (typical punctuation) | | | 105.75 (103.44 to 108.06) | | 99.96 (97.53 to 102.38) | 5.79 (2.44 to 9.14) | **0.001** |
| z-score | | | 0.62 (0.43 to 0.81) | | 0.15 (–0.04 to 0.35) | 0.47 (0.20 to 0.74) | |
| Cognitive flexibility 1 | 109 | 57 | | 52 | | | |
| Raw score (total correct designs) | | | 24.24 (23.02 to 25.46) | | 21.81 (20.53 to 23.09) | 2.43 (0.67 to 4.20) | **0.007** |
| z-score | | | 0.65 (0.47 to 0.84) | | 0.28 (0.08 to 0.48) | 0.38 (0.10 to 0.65) | |
| Cognitive flexibility 2 | 109 | 57 | | 52 | | | |
| Raw score (sec) | | | 80.32 (70.58 to 90.07) | | 88.16 (77.96 to 98.37) | –7.84 (–21.95 to 6.28) | 0.274 |
| z-score | | | 0.27 (0.04 to 0.49) | | 0.08 (–0.15 to 0.32) | 0.18 (–0.15 to 0.51) | |
| Cognitive flexibility composite z-score | 109 | 57 | 0.15 (–0.02 to 0.33) | 52 | –0.17 (–0.36 to 0.02) | 0.32 (0.07 to 0.58) | **0.014** |
| Inhibition | 109 | 57 | | 52 | | | |
| Raw score (sec) | | | 31.24 (28.07 to 34.40) | | 33.53 (30.22 to 36.85) | –2.30 (–6.88 to 2.29) | 0.323 |
| z-score | | | 0.55 (0.37 to 0.74) | | 0.42 (0.22 to 0.61) | 0.14 (–0.13 to 0.41) | |
| Working memory | 109 | 57 | | 52 | | | |
| Raw score (% accuracy) | | | 66.02 (62.79 to 69.25) | | 63.89 (60.51 to 67.28) | 2.13 (–2.57 to 6.82) | 0.371 |
| z-score | | | 0.04 (–0.15 to 0.24) | | –0.09 (–0.29 to 0.12) | 0.13 (–0.16 to 0.41) | |
| Executive function composite z-score | 109 | 57 | 0.14 (–0.03 to 0.31) | 52 | –0.15 (–0.33 to 0.02) | 0.29 (0.05 to 0.53) | **0.019** |

z-score values indicate how many standard deviations have the post-exercise program values changed with respect to the baseline mean and standard deviation. E.g., a 0.50 z-score means that the mean value at post-exercise program is 0.50 standard deviations higher than the mean value at baseline, indicating a positive change, with negative values indicating the opposite.
All data presented were adjusted for baseline values of the outcome studied.
Crystallized, Fluid, and Total Intelligence were measured by the Kaufman Brief Intelligence Test.
Cognitive flexibility 1 was measured by the Design Fluency Test and expressed as number of total correct designs of the three conditions.
Cognitive flexibility 2 was measured by the Trail Making Test and expressed as the total completion time (sec) of Part A subtracted from the total completion time (sec) of Part B. A smaller B – A smaller difference in this score (sec) indicated better cognitive flexibility. Note: In the Figures, the z-score is presented inverted for easier visual interpretation in the same direction than the rest of outcomes, but here in the table are presented the real non-inverted values.
Cognitive flexibility composite z-score was calculated as the re-normalized mean of the z-scores for Cognitive flexibility 1 and Cognitive flexibility 2.
Inhibition was measured by the Stroop Color-Word Test. The inhibition score was obtained by subtracting condition 3 completion time – condition 1 completion time (sec). The smaller the difference between tests' times, the better the performance was considered. Note: In the Figures, the z-score is presented inverted for easier visual interpretation in the same direction than the rest of outcomes, but here in the table are presented the real non-inverted values.
Working memory was measured by the Delayed Non-Match-to sample task. The response accuracy (%) in the high load was used as an indicator of working memory. Higher response accuracy refers to better performance.
Executive function composite z-score was calculated as the re-normalized mean of the z-scores for Cognitive flexibility, Inhibition, and Working memory.

**Supplementary eTable 20. Intention-to-treat effects of the *ActiveBrains* exercise program on raw (standard score) and z-scores of post-exercise academic performance outcomes (Woodcock-Muñoz standardized test).**

| | | | Mean (95% CI) | | | | |
|---|---|---|---|---|---|---|---|
| | $N_{all}$ | N | Exercise group | N | Control group | Difference between groups | *P* |
| Academic skills | 109 | 57 | | 52 | | | |
| Raw score | | | 123.59 (121.14 to 126.04) | | 120.01 (117.44 to 122.58) | 3.58 (0.023 to 7.13) | **0.049** |
| z-score | | | 0.30 (0.15 to 0.46) | | 0.07 (–0.09 to .0.24) | 0.23 (0.001 to 0.46) | |
| Academic fluency | 109 | 57 | | 52 | | | |
| Raw score | | | 105.49 (103.78 to 107.20) | | 105.71 (103.92 to 107.51) | –0.23 (–2.71 to 2.25) | 0.858 |
| z-score | | | 0.12 (–0.02 to 0.26) | | 0.14 (–0.01 to 0.29) | –0.02 (–0.23 to 0.19) | |
| Problem solving | 109 | 57 | | 52 | | | |
| Raw score | | | 102.52 (100.79 to 104.25) | | 100.21 (98.40 to 102.03) | 2.31 (–0.23 to 4.86) | 0.075 |
| z-score | | | 0.31 (0.12 to 0.49) | | 0.06 (–0.14 to 0.25) | 0.25 (–0.03 to 0.53) | |
| Reading | 109 | 57 | | 52 | | | |
| Raw score | | | 110.84 (108.98 to 112.70) | | 109.31 (107.36 to 111.26) | 1.53 (–1.18 to 4.24) | 0.265 |
| z-score | | | 0.19 (0.04 to 0.34) | | 0.07 (–0.08 to 0.22) | 0.12 (–0.09 to 0.34) | |
| Mathematics | 109 | 57 | | 52 | | | |
| Raw score | | | 105.09 (103.02 to 107.16) | | 102.68 (100.51 to 104.85) | 2.41 (–0.63 to 5.44) | 0.119 |
| z-score | | | 0.28 (0.10 to 0.47) | | 0.06 (–0.14 to 0.26) | 0.22 (–0.06 to 0.50) | |
| Writing | 109 | 57 | | 52 | | | |
| Raw score | | | 118.33 (116.07 to 120.58) | | 116.34 (113.98 to 118.70) | 1.99 (–1.27 to 5.25) | 0.230 |
| z-score | | | 0.33 (0.16 to 0.51) | | 0.18 (–0.01 to 0.36) | 0.16 (–0.10 to 0.41) | |
| Total performance | 109 | 57 | | 52 | | | |
| Raw score | | | 112.98 (111.27 to 114.69) | | 110.87 (109.08 to 112.66) | 2.11 (–0.38 to 4.60) | 0.096 |
| z-score | | | 0.29 (0.15 to 0.44) | | 0.11 (–0.04 to 0.27) | 0.18 (–0.03 to 0.39) | |

z-score values indicate how many standard deviations have the post-exercise program values changed with respect to the baseline mean and standard deviation. e.g., a 0.50 z-score means that the mean value at post-exercise program is 0.50 standard deviations higher than the mean value at baseline, indicating a positive change, with negative values indicating the opposite.
All data presented were adjusted for baseline values of the outcome studied.
Academic skills are the sum of components based on basic skills such as reading decoding, mathematics calculation, and spelling.
Academic fluency is the sum of the components based on reading, calculation, and writing fluency.
Problem solving is the sum of the components based on solving academic problems in reading, mathematics, and writing.
Total performance is the overall measure of the academic performance based on reading, mathematics, and writing.

**Supplementary eTable 21. Intention-to-treat effects of the *ActiveBrains* exercise program on raw ($mm^3$) and z-scores of post-exercise hippocampal gray matter volume**.

| | | | Mean (95% CI) | | | | |
|---|---|---|---|---|---|---|---|
| | $N_{all}$ | N | Exercise group | N | Control group | Difference between groups | *P* |
| Hippocampus | 109 | 57 | | 52 | | | |
| Raw score | | | 7096.55 (6996.69 to 7196.42) | | 7190.67 (7086.11 to 7295.23) | –94.11 (–238.83 to 50.60) | 0.200 |
| z-score | | | –0.072 (–0.234 to 0.089) | | 0.079 (–0.089 to 0.248) | –0.152 (–0.385 to 0.082) | |
| Right hippocampus | 109 | 57 | | 52 | | | |
| Raw score | | | 3629.94 (3576.13 to 3683.76) | | 3687.54 (3631.20 to 3743.89) | –57.60 (–135.54 to 20.34) | 0.146 |
| z-score | | | –0.081 (–0.240 to 0.078) | | 0.089 (–0.077 to 0.255) | –0.170 (–0.399 to 0.060) | |
| Right anterior hippocampus | 109 | 57 | | 52 | | | |
| Raw score | | | 2084.18 (2049.70 to 2118.66) | | 2122.24 (2086.14 to 2158.34) | –38.06 (–87.99 to 11.86) | 0.134 |
| z-score | | | –0.088 (–0.254 to 0.079) | | 0.096 (–0.078 to 0.271) | –0.184 (–0.425 to 0.057) | |
| Right posterior hippocampus | 109 | 57 | | 52 | | | |
| Raw score | | | 1544.58 (1521.51 to 1567.65) | | 1560.35 (1536.19 to 1584.50) | –15.77 (–49.20 to 17.66) | 0.352 |
| z-score | | | –0.053 (–0.217 to 0.110) | | 0.059 (–0.113 to 0.230) | –0.112 (–0.349 to 0.125) | |
| Left hippocampus | 109 | 57 | | 52 | | | |
| Raw score | | | 3465.67 (3402.53 to 3528.82) | | 3504.16 (3438.04 to 3570.27) | –38.48 (–130.03 to 53.06) | 0.406 |
| z-score | | | –0.049 (–0.219 to 0.121) | | 0.054 (–0.121 to 0.232) | –0.104 (–0.350 to 0.143) | |
| Left anterior hippocampus | 109 | 57 | | 52 | | | |
| Raw score | | | 1991.96 (1953.98 to 2029.94) | | 2010.78 (1971.01 to 2050.55) | –18.82 (–73.86 to 36.22) | 0.499 |
| z-score | | | –0.041 (–0.213 to 0.132) | | 0.045 (–0.136 to 0.225) | –0.085 (–0.335 to 0.164) | |
| Left posterior hippocampus | 109 | 57 | | 52 | | | |
| Raw score | | | 1474.47 (1444.52 to 1504.41) | | 1489.12 (1457.76 to 1520.48) | –14.66 (–58.09 to 28.78) | 0.505 |
| z-score | | | –0.042 (–0.224 to 0.139) | | 0.046 (–0.144 to 0.237) | –0.089 (–0.352 to 0.175) | |

z-score values indicate how many standard deviations have the post-exercise program values changed with respect to the baseline mean and standard deviation. e.g., a 0.50 z-score means that the mean value at post-exercise program is 0.50 standard deviations higher than the mean value at baseline, indicating a positive change, with negative values indicating the opposite.
All data presented were adjusted for baseline values of the outcome studied.

**Supplementary eTable 22. Descriptive characteristics of the *ActiveBrains* participants that complete the study (i.e., non-dropouts) and those that did not complete the study (i.e., dropouts) at baseline.**

| | All | | Drop-outs | | Non-drop-outs | | P |
|---|---|---|---|---|---|---|---|
| | N | Mean ± SD | N | Mean ± SD | N | Mean ± SD | |
| Age (years) | 109 | 10.04 ± 1.13 | 13 | 10.39 ± 1.27 | 96 | 10.00 ± 1.10 | 0.232 |
| Sex | | | | | | | 0.327 |
| Girls (n %) | 45 | 41% | 7 | 54% | 38 | 40% | |
| Boys (n %) | 64 | 59% | 6 | 46% | 58 | 60% | |
| Weight (kg) | 109 | 56.21 ± 11.23 | 13 | 61.34 ± 11.22 | 96 | 55.52 ± 11.11 | 0.079 |
| Height (cm) | 109 | 144.22 ± 8.41 | 13 | 146.45 ± 8.65 | 96 | 143.92 ± 8.38 | 0.311 |
| Body mass index ($kg/m^2$) | 109 | 26.81 ± 3.62 | 13 | 28.45 ± 3.33 | 96 | 26.59 ± 3.62 | 0.082 |
| Peak height velocity (years) | 109 | –2.26 ± 0.99 | 13 | –1.81 ± 1.21 | 96 | –2.32 ± 0.94 | 0.080 |
| Wave of participation (%) | | | | | | | |
| First (n %) | 19 | 18% | 5 | 39% | 14 | 14% | 0.123 |
| Second (n %) | 45 | 41% | 6 | 46% | 39 | 41% | |
| Third (n %) | 45 | 41% | 2 | 15% | 43 | 45% | |
| Cardiorespiratory fitness | | | | | | | |
| Final time in treadmill test (min) | 108 | 8.55 ± 2.77 | 12 | 7.99 ± 2.86 | 96 | 8.62 ± 2.76 | 0.464 |
| Relative $VO_2max$ (mL/kg/min) | 108 | 37.38 ± 4.75 | 12 | 35.84 ± 4.57 | 96 | 37.58 ± 4.76 | 0.234 |
| Intelligence | | | | | | | |
| Crystallized intelligence (typical punctuation) | 109 | 103.07 ± 12.84 | 13 | 97.92 ± 12.59 | 96 | 103.77 ± 12.78 | 0.124 |
| Fluid intelligence (typical punctuation) | 109 | 97.68 ± 13.13 | 13 | 97.54 ± 12.57 | 96 | 97.7 ± 13.27 | 0.967 |
| Total intelligence (typical punctuation) | 109 | 98.06 ± 12.43 | 13 | 94.92 ± 13.26 | 96 | 98.48 ± 12.33 | 0.335 |
| Executive function | | | | | | | |
| Cognitive flexibility 1 (total correct designs) | 109 | 20.01 ± 6.48 | 13 | 19.15 ± 5.11 | 96 | 20.13 ± 6.66 | 0.614 |
| Cognitive flexibility 2 (sec) | 104 | 90.36 ± 43.12 | 13 | 103.20 ± 41.48 | 91 | 88.53 ± 43.26 | 0.253 |
| Cognitive flexibility composite z-score | 104 | –0.01 ± 1.00 | 13 | –0.06 ± 0.98 | 91 | 0.00 ± 1.01 | 0.831 |
| Inhibition (sec) | 109 | 41.12 ± 19.40 | 13 | 40.04 ± 17.48 | 96 | 41.27 ± 19.73 | 0.832 |
| Working memory (% response accuracy) | 108 | 65.28 ± 17.13 | 13 | 63.47 ± 18.91 | 95 | 65.53 ± 16.97 | 0.686 |
| Executive function composite z-score | 103 | –0.01 ± 1.00 | 13 | –0.10 ± 1.02 | 90 | 0.01 ± 1.00 | 0.725 |
| Academic performance (standard score) | | | | | | | |
| Academic skills | 108 | 118.81 ± 15.69 | 12 | 110.75 ± 20.03 | 96 | 119.82 ± 14.89 | 0.059 |
| Academic fluency | 108 | 104.06 ± 12.06 | 12 | 104.83 ± 15.03 | 96 | 103.97 ± 11.73 | 0.816 |
| Problem solving | 108 | 99.71 ± 9.29 | 12 | 97.92 ± 11.41 | 96 | 99.94 ± 9.03 | 0.480 |
| Reading | 108 | 107.71 ± 13.03 | 12 | 101.17 ± 16.79 | 96 | 108.53 ± 12.35 | 0.213 |
| Mathematics | 108 | 102.04 ± 10.95 | 12 | 99.67 ± 13.21 | 96 | 102.33 ± 10.68 | 0.429 |
| Writing | 108 | 103.79 ± 9.04 | 12 | 104.67 ± 11.46 | 96 | 103.68 ± 8.75 | 0.178 |
| Total academic performance | 108 | 109.52 ± 11.88 | 12 | 105.42 ± 16.19 | 96 | 110.03 ± 11.24 | 0.206 |
| Hippocampal volume ($mm^3$) | 102 | 7036.92 ± 694.08 | 13 | 6629.62 ± 790.46 | 89 | 7096.41 ± 663.04 | **0.023** |

Values are expressed as means ± standard deviations (SD), unless otherwise indicated. Baseline differences between drop-outs and non-drop-outs were determined by one-way analysis of variance (ANOVA) and chi-squared tests for continuous and categorical variables, respectively. Statistically significant values at P < 0.005 are shown in bold.

Intelligence outcomes (i.e., Crystallized, Fluid, and Total Intelligence) were measured by the Kaufman Brief Intelligence Test.

Cognitive flexibility 1 was measured by the Design Fluency Test and expressed as number of total correct designs of the three conditions.

Cognitive flexibility 2 was measured by the Trail Making Test and expressed as the total completion time (sec) of Part A subtracted from the total completion time (sec) of Part B. A smaller B – A difference score (sec) indicated better cognitive flexibility.

Cognitive flexibility composite z-score was calculated as the re-normalized mean of the z-scores for Cognitive flexibility 1 and Cognitive flexibility 2.
Inhibition was measured by the Stroop Color-Word Test. The inhibition score was obtained by subtracting condition 3 completion time – condition 1 completion time (sec). The lower the difference between tests' times, the better the performance was considered.
Working memory was measured by the Delayed Non-Match-to sample task.
Executive function composite z-score was calculated as the re-normalized mean of the z-scores for Cognitive flexibility, Inhibition, and Working memory.
Academic performance was measured by the Spanish version of the Woodcock Johnson III Test of Achievement.
Academic skills are the sum of components based on basic skills such as reading decoding, mathematics calculation, and spelling.
Academic fluency is the sum of the components based on reading, calculation, and writing fluency.
Problem solving is the sum of the components based on solving academic problems in reading, mathematics, and writing.
Total academic performance is the overall measure of the academic performance based on reading, mathematics, and writing.
