## Supplementary material for "Effects of exercise on brain health outcomes in children with overweight/obesity: the ActiveBrains randomized controlled trial": CONSORT checklist for conducting and reporting trials

**This file includes:**

CONSORT checklist for reporting randomized trials.

**Supplement 2. CONSORTchecklist for the Activebrains trial.**

| Section/Topic | Item No | Checklist item | Reported on page No |
|---|---|---|---|
| **Title and abstract** | | | |
| | 1a | Identification as a randomised trial in the title | Page 1 |
| | 1b | Structured summary of trial design, methods, results, and conclusions (for specific guidance see CONSORT for abstracts) | Page 3 |
| **Introduction** | | | |
| Background and objectives | 2a | Scientific background and explanation of rationale | Page 5 |
| | 2b | Specific objectives or hypotheses | Page 5 |
| **Methods** | | | |
| Trial design | 3a | Description of trial design (such as parallel, factorial) including allocation ratio | Page 6 |
| | 3b | Important changes to methods after trial commencement (such as eligibility criteria), with reasons | Supp. file 1, Page |
| Participants | 4a | Eligibility criteria for participants | Supp. file 1, Page 3 |
| | 4b | Settings and locations where the data were collected | Supp. file 1, Page 3 |
| Interventions | 5 | The interventions for each group with sufficient details to allow replication, including how and when they were actually administered | Supp. file 1, Page 3-4 |
| Outcomes | 6a | Completely defined pre-specified primary and secondary outcome measures, including how and when they were assessed | Supp. file 1, Page 5-10 |
| | 6b | Any changes to trial outcomes after the trial commenced, with reasons | NA |
| Sample size | 7a | How sample size was determined | Supp. file 5, Page 10 |
| | 7b | When applicable, explanation of any interim analyses and stopping guidelines | NA |
| Randomisation: | | | |

| | | | |
|---|---|---|---|
| Sequence generation | 8a | Method used to generate the random allocation sequence | Supp. file 1, Page 3 |
| | 8b | Type of randomisation; details of any restriction (such as blocking and block size) | Supp. file 1, Page 3 |
| Allocation concealment mechanism | 9 | Mechanism used to implement the random allocation sequence (such as sequentially numbered containers), describing any steps taken to conceal the sequence until interventions were assigned | Supp. file 1, Page 3 |
| Implementation | 10 | Who generated the random allocation sequence, who enrolled participants, and who assigned participants to interventions | Supp. file 1, Page 3 |
| Blinding | 11a | If done, who was blinded after assignment to interventions (for example, participants, care providers, those assessing outcomes) and how | Supp. file 1, Page 3 |
| | 11b | If relevant, description of the similarity of interventions | NA |
| Statistical methods | 12a | Statistical methods used to compare groups for primary and secondary outcomes | Page 7; Supp. file 1, Page 10-14 |
| | 12b | Methods for additional analyses, such as subgroup analyses and adjusted analyses | Page 7; Supp. file 1, Page 10-14 |
| **Results** | | | |
| Participant flow (a diagram is strongly recommended) | 13a | For each group, the numbers of participants who were randomly assigned, received intended treatment, and were analysed for the primary outcome | Page 8; Fig 1 |
| | 13b | For each group, losses and exclusions after randomisation, together with reasons | Page 20, Fig 1 |
| Recruitment | 14a | Dates defining the periods of recruitment and follow-up | Supp. file 1, Page 40, supp eTable 1 |
| | 14b | Why the trial ended or was stopped | NA |
| Baseline data | 15 | A table showing baseline demographic and clinical characteristics for each group | Supp. file 1, Page 40, supp eTable 1 |

| | | | |
|---|---|---|---|
| Numbers analysed | 16 | For each group, number of participants (denominator) included in each analysis and whether the analysis was by original assigned groups | Page 20, Fig 1 |
| Outcomes and estimation | 17a | For each primary and secondary outcome, results for each group, and the estimated effect size and its precision (such as 95% confidence interval) | Supp. file 1, eTable 2-22 |
| | 17b | For binary outcomes, presentation of both absolute and relative effect sizes is recommended | NA |
| Ancillary analyses | 18 | Results of any other analyses performed, including subgroup analyses and adjusted analyses, distinguishing pre-specified from exploratory | Supp. file 1, supp eTable 2-22 |
| Harms | 19 | All important harms or unintended effects in each group (for specific guidance see CONSORT for harms) | Supp. file 1, Page 5 |
| **Discussion** | | | |
| Limitations | 20 | Trial limitations, addressing sources of potential bias, imprecision, and, if relevant, multiplicity of analyses | Page 12 |
| Generalisability | 21 | Generalisability (external validity, applicability) of the trial findings | Page 11-12; Supp. file 19-22 |
| Interpretation | 22 | Interpretation consistent with results, balancing benefits and harms, and considering other relevant evidence | Page 11-12; Supp. file 19-22 |
| **Other information** | | | |
| Registration | 23 | Registration number and name of trial registry | Page 6 |
| Protocol | 24 | Where the full trial protocol can be accessed, if available | Page 6 |
| Funding | 25 | Sources of funding and other support (such as supply of drugs), role of funders | Page 13-14 |
