## Supplementary material for "Effects of exercise on brain health outcomes in children with overweight/obesity: the ActiveBrains randomized controlled trial": AGReMA checklist for conducting and reporting mediation analyses

### AGReMA[SF]

A Guideline for Reporting Mediation Analyses
Short Form

| Section/Topic | Item Number | Item Description | Reported on page No |
|---|---|---|---|
| **Introduction** | | | |
| Objectives | 1 | State the objectives of the study specific to the mechanisms of interest. The objectives should specify whether the study aims to testor estimate the mechanistic effects | Page 5 (main text) and page 20 (Suppl 1) |
| **Methods** | | | |
| Effects of interest | 2 | Specify the effects of interest | Page 7 (main text) and page 12-13 and page 20-21 (Suppl 1) |
| Causal assumptions | 3 | Specify assumptions about the causal model | Page 12-13 (Suppl 1) |
| Measurement | 4 | Clearly describe the interventions or exposures, mediators, outcomes, confounders, and moderators that were used in the analyses. Specify how and when they were measured, the measurement properties, andwhether blinded assessment was used | Page 5-13 (Suppl 1) |
| Statistical methods | 5 | Describe the statistical methods used to estimate the causal relationships of interest. This description should specify analytical strategies used to reduce confounding, model building procedures, justification for the inclusion or exclusion of possible interaction terms, modelling assumptions, and methods used to handle missing data. Provide a reference to the statistical software and package used | Page 12-13 (Suppl 1) |
| **Results** | | | |
| Participants | 6 | Describe baseline characteristics of participants included in mediation analyses. Report the total sample size and number of participants lostduring follow-up or with missing data | Page 8 (main text) |
| Outcomes and estimates | 7 | Report point estimates and uncertainty estimates for the exposure-mediator and mediator-outcome relationships. If inference concerning the causal relationship of interest is considered feasible given the causal assumptions, report the point estimate and uncertainty estimate | Page 9 (main text) |
| **Discussion** | | | |
| Limitations | 8 | Discuss the limitations of the study including potential sources of bias | Page 12 (main text) |
| Interpretation | 9 | Interpret the estimated effects considering the study's magnitude and uncertainty, plausibility of the causal assumptions, limitations, generalizability of the findings, and results from relevant studies | Page 11 (main text) Page 20 and 21 (Suppl 1) |

*From:* Lee H, Cashin AG, Lamb SE, Hopewell S, Vansteelandt S, VanderWeele TJ, et al. A Guideline for Reporting Mediation Analyses of Randomized Trials and Observational Studies. The AGReMA Statement. JAMA. 2021;326(11):1045–1056. doi:10.1001/jama.2021.14075

AGReMA-SF is designed for articles that report mediation analyses of randomized trials or observational studies as a secondary focus of a paper. AGReMA-SF should be used in conjunction with CONSORT or STROBE for complete reporting.

For more information, visit: agrema-statement.org



(cc) BY-NC-ND
