## Supplementary material for "Effects of exercise on brain health outcomes in children with overweight/obesity: the ActiveBrains randomized controlled trial": Statistical analysis plan

### STATISTICAL ANALYSIS PLAN (SAP)

**Official title: ActiveBrains: Effects of an Exercise-based Randomized Controlled Trial on Cognition, Brain Structure and Brain Function in Overweight/Obese Preadolescent Children**

Registered in https://clinicaltrials.gov/

**NCT Number:** 02295072

**Date of the document:** June 19, 2021

#### Table of contents

1. PURPOSE OF THE SAP 3

2. STUDY DESIGN 3

3. STUDY AIMS 3

3.1. PRIMARY AIMS 3
3.2. SECONDARY AIMS 3

4. OUTCOMES 4

4.1. PRIMARY OUTCOMES 4
4.2. SECONDARY OUTCOMES 5

5. STATISTICAL ANALYSIS 7

5.1. USE OF PER-PROTOCOL AND INTENTION-TO-TREAT PRINCIPLES 7
5.2. ANALYSIS SOFTWARE AND DESCRIPTIVE ANALYSIS 7
5.3. ANALYSIS OF THE EFFECTS OF THE ACTIVEBRAINS EXERCISE PROGRAM 8
5.4. MEDIATION ANALYSES 8
5.5. MODERATION ANALYSES 9
5.6. INTENTION-TO-TREAT AND DROPOUT ANALYSES 9
5.7. TESTING POTENTIAL COMPENSATORY AND CONTAMINATION EFFECTS OF THE INTERVENTION ON OVERALL ACTIVITY LEVELS, AND ANALYSES ON THE INTENSITY OF THE TRAINING SESSIONS 10

6. REFERENCES 10

### Statistical Analysis Plan (SAP)

#### 1. PURPOSE OF THE SAP

The present document contains a wide description of the statistical analysis plan (SAP) aimed to detail the different outcome measures of study and computations performed as well as all sets of analyses of the ActiveBrains randomized controlled trial (RCT).

The present SAP focused on the step 1 analysis performed to determine whether a 20-week exercise program have a significant benefit over intelligence, executive function, academic performance, and brain outcomes in comparison with a control group. This SAP also informs about all secondary analyses to test potential mediators and moderators of the main exercise effects observed in this intervention. All these analyses will be performed by the principle investigator and trial coordinator (Francisco B. Ortega) and its research team from the PROFITH (PROmoting FITness and Health Through Physical Activity) Research Group at the Sport and Health University Research Institute (iMUDS), Department of Physical and Sports Education, Faculty of Sport Sciences, University of Granada, Granada, Spain. In next steps, we will investigate exercise effects on secondary outcomes (i.e., physical and mental health, white matter integrity, neuroelectric measurements, bone health outcomes, sleep-related outcomes, molecular outcomes, and functional magnetic resonance imaging [fMRI]) of the Activebrains RCT study.

#### 2. STUDY DESIGN

The ActiveBrains project is an individual randomized controlled trial (1:1) with an experimental group that participates in a 20-weeks physical exercise programme and a waiting-list control group that keeps usual lifestyle. We will obtain two different time points to test the effects of exercise, a baseline outcome and a post-intervention outcome. Details of the ActiveBrains project have been described elsewhere (1).

#### 3. STUDY AIMS

##### 3.1. Primary aims

Our primary aim is to investigate the effects of a 20-week exercise program on behavioral outcomes, including intelligence, executive functions (i.e., cognitive flexibility, inhibition and working memory) and academic performance, as well as on brain outcomes (i.e., hippocampal volume as a primary region of interest) in children with overweight/obesity.

##### 3.2. Secondary aims

The secondary aims are to explore potential mediators and moderators of the main exercise effects observed in this intervention: 1) we will perform exploratory analyses on specific brain regions of interest (e.g., subregions of the hippocampus and prefrontal cortex) as well as broader-brain hypothesis-free exploratory analyses with the purpose of identifying structural and functional brain changes that could mediate the effects of the exercise intervention on behavioral outcomes;

2) we will investigate cardiorespiratory fitness (CRF) as the main candidate mediator; 3) we will test potential moderators of the intervention effects, such as sex, age, biological maturation, socioeconomic status and baseline levels of specific outcomes studied; 4) we will test potential compensatory and contamination effects of the intervention and control groups, respectively, on overall activity levels objectively-assessed by 24-hour worn accelerometers.

### 4. OUTCOMES

All outcomes explained below will be collected and defined in two different time points for analyses: baseline (pre-intervention) and post-intervention. Some of the outcomes will be defined only for the baseline time points for moderation analyses.

#### 4.1. Primary outcomes

##### *Intelligence*

Intelligence is assessed by the Spanish version of the Kaufman Brief Intelligence Test (K-BIT) (2). The vocabulary subtest provides an estimated crystallized intelligence score and the matrices subtest provides an estimated fluid intelligence score. We will use the age-specific percentiles for both crystallized and fluid scores, and total intelligence will be calculated from their sum.

##### *Executive function*

The three core-dimensions of executive function will be evaluated in this study: cognitive flexibility, inhibition, and working memory (3). A full description of cognitive flexibility, inhibition, and working memory tests can be found elsewhere (4,5).

- Cognitive flexibility will be assessed using the Design Fluency Test and the Trail Making Test (6). The total number of correctly drawn designs from all three conditions of the Design Fluency Test will be used as indicator of cognitive flexibility 1 in main analysis. Higher number of correctly drawn designs refers to better cognitive flexibility performance. In the Trail Making Test, the total completion time of Part B will be subtracted from the total completion time of Part A and will be used as an indicator of cognitive flexibility 2 in main analysis (7). A smaller B – A difference (sec) indicated better cognitive flexibility.
- Inhibition will be measured via a modified version of the Stroop test (paper-pencil version) (6). An inhibition score will be calculated for main analysis as: condition 3 completion time minus condition 1 completion time (sec) (8). The lower the difference between test times (sec), the better the performance.
- Working memory will be measured by a modified version of the Delayed Non-Match-to-Sample (DNMS) computerized task (9). Response accuracy (%) in the high load will be used as an indicator of working memory. Higher response accuracy refers to better working memory performance.

##### *Academic performance*

Academic performance will be assessed by the Spanish version of the Woodcock-Johnson III Tests of Achievement (10). Standard scores of reading, mathematics, writing, academic skills (i.e., the sum of tests based on basic skills such as reading decoding, mathematics calculation and spelling), academic fluency (i.e., the sum of tests based on reading, calculation and writing fluency), problem solving (i.e., the sum of tests based on solving academic problems in reading, mathematics and writing) and total performance (i.e., the overall measure of academic performance based on reading, mathematics and writing) will be used for analyses.

##### *Brain MRI outcomes*

All images will be obtained using a 3.0 Tesla Siemens Magnetom Tim Trio scanner (Siemens Medical Solutions, Erlangen, Germany) with a 32-channel head coil. Volume and shape of the hippocampus will be extract using the FMRIB's Integrated Registration and Segmentation Tool (FIRST) (11), a semi-automated model-based segmentation tool in FMRIB's Software Library (FSL) version 5.0.7. We will extract the volume in $mm^3$ of the hippocampus to be included in the main analysis. In addition, the hippocampus segmentation will be split into anterior and posterior sub-regions for each hemisphere separately and its volume ($mm^3$) will be obtained.

#### 4.2. Secondary outcomes

##### *Brain outcomes*

For secondary analyses, volume ($mm^3$) of other subcortical regions, different from hippocampus, are segmented for both hemispheres separately (i.e., 6 per hemisphere: nucleus accumbens, amygdala, caudate, globus pallidum, putamen, and thalamus; and brain stem) will be extracted.

Using FreeSurfer software version 5.3.0 we will extract cortical thickness (mm), surface area ($mm^2$) and volume ($mm^3$) of prefrontal cortex sub-regions (i.e., 6 per hemisphere: cingulate gyrus, anterior division; inferior frontal gyrus, pars opercularis; inferior frontal gyrus, pars triangularis; middle frontal gyrus; superior frontal gyrus and frontal orbital cortex) and we will use the extracted thickness, area, and volume of each sub-region as outcome in the group-level analysis (12–14).

Another secondary outcome will be the functional connectivity between hippocampus and prefrontal cortex (15). The residualized parameter estimate maps will be converted to z scores (via Fishers r to z transformation) to achieve normality and will be entered into higher level analyses. For the hippocampal connectivity, we will use the anterior and posterior sub-regions for each hemisphere separately, as seeds.

We will use the SPM software (SPM 12; Wellcome Department of Cognitive Neurology, London, UK) for the whole-brain volumetric analyses (16). Total gray and white matter volumes will be derived from segmented images, and total brain volume will be calculated by adding the volumes of gray and white matter. In addition, we will use the voxel-wise functional connectivity network maps of blood-oxygen-level-dependent (BOLD) signals for functional connectivity analysis between hippocampus seed and prefrontal cortex.

Finally, Non-negative Matrix Factorization (NNMF) analysis will be used to identify structural networks. NNMF is a method for extracting structural networks where volume covaries across all participants (17). Smoothed structural gray matter images (all processing information can be found in detail elsewhere (16)) for each subject were reshaped into a matrix including all available pre- and post- images for a high-quality accuracy of the structural networks.

##### *Cardiorespiratory fitness*

For analyses, we will use peak oxygen consumption ($VO_2$peak, mL/kg/min) and final completion time (min) of a maximal incremental treadmill test, namely time-to-exhaustion, as indicators of CRF. Particularly for mediation analyses, we will use a delta (Δ) of change between post-intervention CRF and pre-intervention CRF.

##### *Biological maturation*

A baseline measurement of years from peak height velocity (PHV) will be calculated by subtracting the age of PHV from chronological age, so that it is interpreted as how many years from the PHV offset a person is, with a value ranging from negative values (before the PHV; less mature) to positive values (after the PHV; more mature) (18,19). This baseline outcome will be used in moderation analyses

##### *Socioeconomic status*

We will compute a dichotomized parental combined variable for the educational level at baseline as low (neither parent had university education) and high (at least one of the parents had university education), to be used in the moderation analyses (20,21). In addition, parents' answers on their occupation at baseline will be categorized as high (1 to 3), medium (4 to 8), and low (9 to 12). Afterwards, we will compute a final dichotomized parental combined variable for the occupational level at baseline as low (neither parent had a high occupational level) and high (at least one of the parents had a high occupational level), to be used in the moderation analyses.

##### *Other secondary outcomes: physical and mental health*

In addition to the outcomes related to the primary objectives described above, we will apply the same statistical plan and approaches to investigate the effects of this intervention on a set of physical health outcomes (cardio metabolic risk factors and bone health) and mental health outcomes (depression, anxiety, optimism, happiness, among others).

##### *Overall activity assessment before and during the intervention*

We will determine the changes in overall activity levels in children from both groups from baseline to during the intervention (i.e., in the middle of the intervention, week 10) (22,23). For the secondary analyses proposed herein, the ENMO with negative values rounded to zero of the raw accelerations of accelerometers worn on the right hip will be used as an indicator of overall activity (24–27). Same procedures will be performed over the accelerations from the non-dominant wrist and used in sensitivity analyses.

### 5. STATISTICAL ANALYSIS

#### 5.1. Use of per-protocol and intention-to-treat principles

Two analysis will be defined: the per-protocol analysis and the intention-to-treat (ITT). We will use the per-protocol principle to report the main findings for all behavioral and MRI outcomes in all children with overweight/obesity that followed these criteria: (1) completed the study and the pre- and post-intervention assessments, and (2) attended at least 70% of the recommended 3 sessions/week (i.e., exercise group). Main analyses will be performed using the per-protocol criteria for two reasons: 1) we are interested in knowing the efficacy rather than effectiveness of our intervention, i.e., we want to know the effects on brain health outcomes when a child actually does the planned exercise program (operationally defined as attending a minimum of 70% of the sessions); and 2) in the field of neuroimaging, with analyses conducted directly on images, it is rarely done and technically difficult, to apply imputation methods on images missing at post-exercise evaluations. Therefore, participants who complete both pre- and post-intervention evaluations are usually included in analyses.

Moreover, we will additionally analyze the data using the ITT for the primary *a priori*-planned analyses only. Under the ITT principle, we will use multiple imputation for observations lost at post-intervention (28) (additional information on multiple testing is available in Section 5.6). We divide our analyses and findings into those *a priori*-planned as primary outcomes when the study was designed, and those explanatory analyses *a posteriori*-planned to further understand and interpret our main findings.

#### 5.2. Analysis software and descriptive analysis

The statistical procedures will be performed using the SPSS software (version 20.0, IBM Corporation) and R software (v. 3.1.2, https://www.cran.r-project.org/). A significant difference level of $P < 0.05$ will be set. Additionally, we investigated which of the significant findings persisted after adjustment for multiple testing on the primary outcomes (29).

Characteristics of the study sample will be given as mean and SD, or frequency and percentage, as appropriate. A CONSORT flow diagram will be created to display the progress of all participants through the trial. The number of participants in the per-protocol and ITT analyses will be given and reported reasons for exclusion from the per-protocol analysis will be summarized.

##### 5.3. Analysis of the effects of the ActiveBrains exercise program

Originally, as a general approach, the main effects of the exercise programme versus control on the study outcomes were expected to be examined by means of one-way analysis of variance (ANOVA), using the pre-post differences as outcome and study group as fixed factor so that we could test whether the changes observed significantly differed between exercise and control groups. This method is equivalent to the one used in a previous major RCT that also tested the effect on cognitive outcomes (30) and used ANalysis of COVariance (ANCOVA) including post-intervention outcomes as dependent variables, group (i.e., exercise vs. control) as a fixed factor, and baseline data of the study outcome as covariate. The inclusion of the study outcome baseline value as covariate and the post-intervention outcome as dependent is equivalent to study the change in the outcome, and therefore this model indicates the time x group interaction intended to know the effects of the intervention.

Raw scores from each outcome were first winsorized (when needed) to limit the influence of extreme values; this method consists of replacing extreme high/low values for the closest (highest/lowest) valid value (30). The z-scores for each outcome at post-exercise program were also formed by dividing the difference of the raw score of each participant from the baseline mean by the baseline standard deviation (i.e., (post-exercise individual value – baseline mean) / baseline SD). This way of reporting the effects has been used in previous major RCT focused on cognitive outcomes (30) and has two main advantages: 1) provides standardized estimates that allow comparisons among outcomes with different original units of measure, which are often abstract and not-intuitive in cognitive testing; and 2) these z-scores of change can be interpreted as effect size indicators, e.g. 0.5 z-score means that the mean value at post-exercise program is 0.5 SDs higher than the mean value at baseline, indicating a positive change, with negative values indicating the opposite. As effect size indicators, they can be interpreted according to the standard benchmarks, i.e. a value around 0.2 is considered a small effect size, 0.5 is considered a medium effect size and 0.8 is considered a large effect size (31).

MRI data need to be handled using methods specifically developed for this field. Our group has expertise in MRI data analysis and will choose the best approach to every single research question addressed.

##### 5.4. Mediation analyses

We will test whether the effects of the intervention on the main study outcomes will be mediated by changes in CRF following the bootstrapping method (32). Mediation analyses will be performed using the PROCESS macro for SPSS (SPSS Inc., Chicago, Illinois) with a resample procedure of 5,000 bootstrap samples. These mediation analyses will be performed for the outcomes for which significant differences are observed between exercise and control groups in main effect analyses. The unstandardized (B) and standardized (β) regression coefficients will be presented for four equations: Equation 1 regressed the mediator (e.g., change in CRF) on the independent variable (group). Equation 2 regressed the dependent variables (i.e., executive function or academic performance outcomes) on the independent variable. Equation 3 regressed the dependent variables on both the mediator (equation 3) and the independent variable (equation 3'). We will also include the outcome of interest at baseline as a confounder. The indirect effects along with its confidence intervals (CIs) will be given and the significance will be considered if the indirect effect significantly differ from zero (i.e., zero is not contained within the CIs). Finally, the percentage of the total effect will be computed to know how much of the total effect is explained by the mediation, as follows: (indirect effect / total effect) × 100. This mediation analysis will be performed using the CRF outcomes (time-to-exhaustion and $VO_2$peak) as mediator variables. The same modeling will be applied to test whether the effects observed on academic performance outcomes will be mediated by the exercise-induced changes in executive function or intelligence outcomes.

##### 5.5. Moderation analyses

In order to explore whether the effects of the intervention were modified by potential moderators, we will run the same models as for the main effects' analyses but stratifying the analyses by sub-groups of populations according to sex (boys vs. girls), age (8-9 vs. 10-11 years of age), biological maturation (below and above the median of PHV), parental educational level (low vs. high), parental occupational level (low vs. high), and baseline outcome levels (below and above the median). In a first step, visual inspection of the effects sizes by subgroups shown in plots will be used to observe the consistency of the effects across potential moderators. In a second step, we will run ANCOVA models to test whether the interaction term (e.g. sex × group, age × group, etc.) in those cases is significant.

##### 5.6. Intention-to-treat and dropout analyses

For the ITT analysis, we will perform multiple imputation to predict missing values at post-exercise outcomes using the predictive mean matching approach. We will therefore perform 10 iterations to create 5 databases, which will be then averaged to obtain the imputed values for the ITT analyses (28). Once we have a dataset with imputed data when missing for the whole sample of study initially allocated into the study groups, we will run the same models to test the effects of the intervention as described above.

In addition, we will use a one-way ANOVA to test whether the participants that complete the baseline evaluations and randomization, but leave the study during the intervention period or do not complete the post-exercise evaluations (i.e., namely the dropouts), differ in the main study outcomes at baseline from the participants who complete the study and post-exercise evaluations (i.e., namely the non-dropouts).

##### 5.7. Testing potential compensatory and contamination effects of the intervention on overall activity levels, and analyses on the intensity of the training sessions

We will perform a 1-dimension curve analysis using SPM1D package available for MATLAB (http://www.spm1d.org) (33) to study whether acceleration values (i.e., expressed as ENMO [mg]) identify a significant increase in physical activity during the exercise program in comparison with the physical activity pattern at baseline for the control and exercise groups. SPM1D is a statistical parameter mapping tool using random field theory and can perform conventional statistics on 1-dimensional data, as is the case of the waveform acceleration data. Weekly average acceleration curves will be presented separately for exercise and control groups from midnight (i.e., 00:00 AM) to next midnight, i.e., 24 h curves centered at noon (12:00 PM). Paired t-tests over the curves will be used to identify significant differences between baseline and exercise patterns for each group throughout the day. SPM involves 4 steps to compute the t-test analysis (34): 1) computing the value of a test statistic at each point in the normalized time series; 2) estimating temporal smoothness on the basis of the average temporal gradient; 3) computing the value of test statistic above which only $\alpha = 5\%$ of the data would be expected to reach had the test statistic trajectory from an equally smooth random process; 4) computing the probability that specific supra threshold regions could have resulted from an equivalently smooth random process. Finally, we will test sex differences in the intensity achieved during exercise sessions using one-way ANOVA with different heart rate outcomes in separate models and sex as fixed factor.
